# Spinal Cord Stimulation for Persistent Spinal Pain Syndrome Type II: A Systematic Review and Subgroup Meta-analysis of Randomized Controlled Trials

**DOI:** 10.64898/2026.02.20.26346691

**Authors:** Pouria Delbari, Ramtin Pourahmad, Amir hessam Zare, Muhammad Hussain Ahmadvand, Saba Sabet, Kimia Rasouli, Martin Jakobs

## Abstract

**Background:** Persistent Spinal Pain Syndrome (PSPS) type II represents a challenging clinical entity with limited therapeutic options. Various spinal cord stimulation (SCS) modalities have emerged as potential treatments, but their comparative effectiveness remains unclear.

**Objective:** Our goal in this paper is to systematically evaluate and compare the efficacy of different SCS modalities in patients with PSPS type II through meta-analysis of available randomized controlled trials.

**Evidence Review:** We conducted a systematic review following PRISMA guidelines, searching major databases for randomized controlled trials evaluating SCS modalities in PSPS type II patients until the end of May 2025(search updated on October 3^rd^). Primary outcomes included pain intensity (VAS) and functional disability (ODI) at 6 and 12 months. Subgroup analyses compared tonic versus burst stimulation and high-frequency versus low-frequency SCS.

**Findings:** Nine randomized controlled trials were included, encompassing 565 patients across different SCS modalities. For the primary outcome of clinically meaningful pain relief (≥50% reduction), pooled analysis demonstrated that 45% (95% CI: 18-75%, I² = 92.2%) of patients achieved this threshold for back pain and 55% (95% CI: 45-65%, I² = 0%) for leg pain. Subgroup analysis revealed significant differences in back pain responder rates by stimulation modality: High-frequency SCS demonstrated responder rates of 92% (95% CI: 79-98%) versus 28% (95% CI: 13-49%) for conventional frequencies (p < 0.001). For leg pain, no significant difference was observed between tonic (51%, 95% CI: 37-65%) and burst stimulation (60%, 95% CI: 45-74%, p = 0.36) and mean VAS scores demonstrated significantly lower pain with high-frequency SCS (13.30, 95% CI: 8.82-17.78) compared to conventional frequency (28.42, 95% CI: 24.02–32.88, p<0.0001). For back pain, mean VAS scores decreased from a baseline of 73.03 to 41.67 (95% CI: 36.12-47.22, I²=22.8%) at 6 months and remained stable at 35.66 (95% CI: 25.39-45.93, I²=75.0%) at 12 months. Leg pain showed more pronounced improvement, with VAS scores declining from a baseline of 61.81 to 23.75 (95% CI: 17.69-29.81, I²=78.8%) at 6 months and 29.16 (95% CI: 24.81-33.52, I²=0%) at 12 months). Meta-regression identified longer pain duration and older age as positive predictors of response, while higher baseline leg pain predicted lower responder rates. Serious adverse events occurred in 10%, with a 16% revision surgery rate. Only two studies demonstrated a low risk of bias across all domains.

**Conclusions:** Current evidence demonstrates that various SCS modalities provide clinically meaningful pain relief in PSPS type II patients, with approximately half achieving ≥50% pain reduction. High-frequency SCS shows significantly superior responder rates for back pain compared to conventional tonic stimulation, while burst stimulation yields significantly superior reductions in continuous pain intensity metrics. However, the limited number of studies, substantial heterogeneity, and lack of head-to-head comparisons prevent definitive recommendations regarding optimal stimulation parameters. Future large-scale randomized trials with standardized protocols and responder-based outcomes are needed to establish evidence-based treatment algorithms for PSPS type II patients.

## Introduction

After lumbar spine surgery, 10–40% of patients experience Persistent Spinal Pain Syndrome Type II (PSPS-T2), formerly called Failed Back Surgery Syndrome (FBSS), which poses a significant healthcare burden ^1,2^. These patients experience persistent or recurrent pain despite anatomically successful procedures, leading to significant functional disability and diminished quality of life. Clinicians lack clear evidence-based guidelines for therapeutic escalation because of the diverse pathophysiology involving epidural fibrosis, arachnoiditis, recurrent disc herniation, and central sensitization, which has hindered the development of standardized treatment algorithms ^1,2^.

For more than thirty years, the main neuromodulation strategy for PSPS-T2 has been spinal cord stimulation. For radicular leg pain neuromodulation, traditional low-frequency tonic spinal cord stimulation (SCS) (40–60 Hz) shows responder rates of roughly 50–60%, with established efficacy validated in randomized controlled trials ^3^. However, response rates are significantly lower, and its efficacy for axial low back pain, which is frequently the main cause of disability remains inconsistent ^4^. This therapeutic gap has driven development of alternative waveforms, including paresthesia-free burst stimulation and high-frequency (10 kHz) SCS ^5^, both showing promise in early trials for treating refractory back pain. SCS paradigms have evolved from traditional tonic waveforms to include various rhythmic stimulation patterns. While the original BurstDR™ waveform (Abbott Neuromodulation, Austin, TX, USA) utilizes a specific, patented five-spike pulse train ^6^, other manufacturers have introduced ‘burst-like’ rhythmic stimulation paradigms. While the clinical superiority of one rhythmic pattern over another remains a subject of debate ^7^, for the purposes of this systematic review, these are collectively analyzed as sub-threshold rhythmic stimulation to evaluate their broad efficacy compared to tonic paradigms.

Despite accumulating randomized controlled trial data, comparative effectiveness between SCS modalities remains unclear. Previous systematic reviews have synthesized this evidence using mean pain score changes as the primary outcome ^8^. While demonstrating statistical differences, this approach provides limited clinical utility for individual patient selection. Mean changes are susceptible to skewed distributions and may mask bimodal response patterns where some patients achieve substantial relief while others experience no benefit ^9,10^, a characteristic feature of neuromodulation therapies. Furthermore, existing meta-analyses report extreme statistical heterogeneity (I² >75%) ^3^, indicating fundamental differences in study populations, protocols, or outcome measurement that challenge the validity of pooled estimates.

Two critical questions remain unanswered for clinical practice. First, what proportion of PSPS-T2 patients can expect clinically meaningful improvement with each SCS modality? The Initiative on Methods, Measurement, and Pain Assessment in Clinical Trials (IMMPACT) recommends that ≥50% pain reduction represents substantial improvement ^9^, a threshold that correlates with patient-reported satisfaction and functional recovery. Responder-based analysis using this threshold provides actionable information, not whether average scores change statistically, but whether individual patients achieve meaningful benefit. Second, do the relative merits of different modalities vary by pain phenotype, such that tonic stimulation might remain optimal for leg-dominant pain while burst or high-frequency approaches provide superior axial pain relief? These questions directly inform treatment selection, particularly as the modalities differ substantially in cost, programming complexity, and the presence or absence of paresthesia^11,12^.

Our systematic review addresses these gaps through responder-based analysis of all available randomized controlled trials comparing SCS modalities in PSPS-T2. By focusing on the proportion of patients achieving ≥50% pain reduction rather than mean score changes, we provide clinically interpretable estimates of treatment success rates. Our prespecified subgroup analyses stratified by both stimulation modality (tonic vs. burst; high-frequency vs. low-frequency) and pain location (axial vs. radicular) systematically investigate whether modality choice explains observed heterogeneity and whether treatment effects differ for distinct pain phenotypes. This approach clarifies the comparative benefits of available SCS technologies and identifies critical evidence gaps requiring future head-to-head trials.

## Methods

### Protocol Registration and Reporting

This systematic review was prospectively registered in the International Prospective Register of Systematic Reviews (PROSPERO: CRD420251138521) and conducted in accordance with the Preferred Reporting Items for Systematic Reviews and Meta-Analyses (PRISMA) 2020 guidelines.

### Search Strategy

A comprehensive systematic literature search was performed across three major electronic databases: PubMed/MEDLINE, Embase, and Scopus from inception through May 28, 2025 then we updated our search on 3 October 2025. The search strategy employed a combination of medical subject headings (MeSH) and free-text terms related to persistent spinal pain syndrome, failed back surgery syndrome, and spinal cord stimulation. Key search terms included: “Persistent spinal pain syndrome,” “PSPS,” “FBSS,” “failed back surgery syndrome,” and “spinal cord stimulation” in various combinations using Boolean operators. The complete search strings for each database are provided in the supplementary materials. The search was restricted to English-language publications.

### Eligibility Criteria

#### Study Design

Only randomized controlled trials (RCTs) were eligible for inclusion. Conference abstracts, case series, observational studies, and non-randomized comparative studies were excluded.

#### Population

Studies enrolling adult patients diagnosed with Persistent Spinal Pain Syndrome Type II (PSPS-T2), also referred to as Failed Back Surgery Syndrome (FBSS), were included. PSPS-T2 was defined as persistent or recurrent back and/or leg pain following at least one anatomically successful spinal surgery. No restrictions were applied regarding the number of previous surgical interventions, patient age, baseline pain scores, or pain duration.

#### Intervention and Comparators

Trials evaluating any modality of spinal cord stimulation were eligible, including conventional tonic stimulation, burst stimulation, high-frequency stimulation, and other novel waveforms. Studies comparing different SCS modalities against each other, conventional medical management, sham stimulation, or placebo were included.

#### Outcomes

Eligible studies were required to report at least one of the following outcomes at 6-month or 12-month follow-up: pain intensity measured by visual analog scale (VAS) or for back pain and/or leg pain, functional disability assessed by the Oswestry Disability Index (ODI), proportion of responders achieving ≥50% pain reduction, or adverse events. Studies reporting outcomes only at earlier time points were excluded.

#### Exclusions

Studies were excluded if they: (1) enrolled patients with mixed pain etiologies where PSPS-T2 patients could not be analyzed separately, (2) included only trial stimulation periods without permanent implantation data, or (3) lacked sufficient data for meta-analysis extraction. Additionally, case reports, congress papers, abstracts, and case series with fewer than 5 patients were excluded from the study.

### Study Selection Process

Two independent reviewers (R.P and S.S) screened all titles and abstracts identified through the database searches. Full-text articles of potentially eligible studies were then independently reviewed by both reviewers against the predetermined eligibility criteria. Disagreements at any stage were resolved through discussion and consensus. If consensus could not be reached, a third senior reviewer was consulted.

### Data Extraction

Two reviewers independently extracted data from each included study using a standardized data extraction form. The following information was systematically extracted: (1) study characteristics (first author, publication year, country, study design, sample size); (2) patient demographics (age, sex, number of previous surgeries); (3) intervention details (SCS modality, stimulation parameters including frequency, pulse width, amplitude); (4) comparator details; (5) outcome measures at 6 and 12 months including mean VAS scores for back pain, leg pain, and overall pain when reported separately, mean ODI scores, number of responders achieving ≥50% pain reduction, and adverse events; and (6) loss to follow-up. For responder analysis, we extracted the raw number of patients achieving ≥50% pain reduction and total number of patients in each group when available. When responder data were reported only as percentages, we calculated the number of responders based on the reported sample size. Discrepancies in data extraction were resolved through discussion and re-examination of the original publications.

### Risk of Bias Assessment

The methodological quality of included RCTs was independently assessed by two reviewers using the revised Cochrane Risk of Bias tool (RoB 2.0). The following domains were evaluated:

1. bias arising from the randomization process, (2) bias due to deviations from intended interventions, (3) bias due to missing outcome data, (4) bias in measurement of the outcome, and (5) bias in selection of the reported result. Each domain was rated as “low risk,” “some concerns,” or “high risk” of bias. An overall risk of bias judgment was generated for each study based on domain-specific assessments. Disagreements were resolved through discussion.

### Outcome Measures

#### Primary Outcome

The primary outcome was the proportion of patients achieving clinically meaningful pain relief, defined as ≥50% reduction in pain intensity from baseline on the VAS (0-100 mm scale) or <u>Oswestry Disability Index</u> (ODI) at 6-month and 12-month follow-up. This threshold was selected based on Initiative on Methods, Measurement, and Pain Assessment in Clinical Trials (IMMPACT) recommendations for substantial pain improvement in chronic pain trials. Responder rates were analyzed separately for back pain, leg pain, and overall pain when studies reported these outcomes distinctly.

#### Secondary Outcomes

Secondary outcomes included: (1) mean VAS/ODI scores for back pain at 6 and 12 months, (2) mean VAS/ODI scores for leg pain at 6 and 12 months, (3) mean VAS/ODI scores for overall pain at 6 and 12 months, (4) mean Oswestry Disability Index scores at 6 and 12 months, and (5) adverse events and device-related complications.

### Data Synthesis and Statistical Analysis

All statistical analyses were performed using R statistical software (version 4.x, R Foundation for Statistical Computing, Vienna, Austria) with the *meta* and *metafor* packages ^13^. All forest plots and funnel plots are available in the supplementary file.

#### Meta-analysis of Responder Rates

For dichotomous outcomes (proportion of responders achieving ≥50% pain reduction), we calculated pooled proportions with 95% confidence intervals using the DerSimonian-Laird random-effects model. The logit transformation was applied to stabilize variances before pooling. Results were back-transformed for presentation as proportions with 95% confidence intervals.

#### Meta-analysis of Continuous Outcomes

For continuous outcomes (VAS and ODI scores), we calculated weighted mean differences with 95% confidence intervals using random-effects models. When studies reported outcomes on different scales (e.g., VAS 0-100), scores were converted to a common scale (VAS 0-100) for pooling.

#### Subgroup Analyses

Pre-specified subgroup analyses were conducted to compare: (1) conventional tonic stimulation versus burst(The ‘Burst’ subgroup includes both the patented BurstDR™ paradigm (Al-Kaisy et al.^14^) and other rhythmic stimulation patterns, such as the burst-like waveforms utilized in other papers)stimulation, and (2) high-frequency SCS (defined as ≥1 kHz based on the original study classifications) versus low-frequency SCS (<1 kHz).

Subgroup comparisons were performed using interaction tests, with statistical significance set at p < 0.05. Meta-regression was employed to explore potential sources of heterogeneity, examining covariates including mean patient age, proportion of males, baseline pain scores, and follow-up duration.

#### Meta-Regression Analysis

To explore potential sources of heterogeneity, univariable meta-regression analyses were conducted using the metafor package in R. Continuous moderators examined included mean patient age, mean body mass index (BMI), proportion of female patients, baseline pain scores (back pain VAS, leg pain VAS, and ODI), pain duration (years), trial stimulation duration (days), trial success rate (%), and publication year. Meta-regression was performed for primary and secondary outcomes when at least three studies provided sufficient data for the covariate of interest. The significance of moderators was assessed using the Knapp-Hartung method with p-values <0.05 considered statistically significant. The proportion of between-study variance explained by each moderator was quantified using R² analog statistics. Only statistically significant moderators with clinically meaningful effect sizes are reported in the results section, the complete result of meta-regression is available as meta-regression table in the supplementary file.

#### Assessment of Heterogeneity

Statistical heterogeneity across studies was assessed using the I² statistic, Cochran’s Q test, and tau² (between-study variance). I² values of 25%, 50%, and 75% were considered to indicate low, moderate, and substantial heterogeneity, respectively. When substantial heterogeneity (I² >75%) was detected, we explored potential sources through subgroup analysis and meta-regression.

#### Publication Bias

Publication bias was assessed visually using funnel plots and statistically using the trim-and-fill method. Asymmetry in funnel plots was interpreted as potential evidence of publication bias or small-study effects.

#### Sensitivity Analyses

Sensitivity analyses were planned to assess the robustness of findings by: (1) excluding studies at high risk of bias, (2) excluding studies with high loss to follow-up (>20%), and (3) using fixed-effect models instead of random-effects models.

## Findings

### Study Selection Process

The systematic literature search identified 1,132 records across three databases: PubMed (n=609), Embase (n=878), and Scopus (n=968). After removing 1132 duplicate records, 1323 unique citations underwent title and abstract screening. At this stage, 951 records were excluded as they did not meet the predefined inclusion criteria. The remaining 372 records proceeded to full-text assessment. Of the 372 reports assessed for eligibility, 227 were excluded for the following reasons: No separate result for SCS (n = 33) and no separate result PSPS-T2(194). 122 were not randomized controlled trials, and 7 studies had data overlap with other included publications and 6 studies were crossover studies, which could not be analyzed with our included papers ^15–20^. Ultimately, 9 studies met all inclusion criteria and were included in this systematic review and meta-analysis. The study selection process is presented in Figure S1 according to the PRISMA 2020 guidelines ^21^.

### Study Characteristics

The 9 included randomized controlled trials enrolled a total of 565 patients with PSPS-T2 The studies were conducted across multiple countries, including the Netherlands, United States, France, and the United Kingdom, with one multinational trial. Most studies were published between 2018 and 2021. Patient demographics varied across studies. Among studies reporting age data, the mean age of participants ranged from 45.1 to 59.4 years. The proportion of female participants ranged from 42% to 85.7% across studies. Several studies did not report complete demographic data for individual treatment arms.

The included trials evaluated various SCS modalities and stimulation parameters. Seven study arms evaluated SCS alone as a single intervention, while six arms assessed combination therapies involving SCS with other treatments. Regarding waveform types, the majority of studies utilized tonic stimulation, while two study arms investigated burst waveform SCS. Stimulation frequency varied, with most studies employing low-frequency protocols, though some investigated high-frequency stimulation or compared both low and high-frequency approaches. All studies compared different SCS modalities or SCS versus control interventions in patients with PSPS Type II following failed back surgery. The primary outcomes assessed included pain intensity measured by VAS score or ODI score, and the proportion of patients achieving clinically meaningful pain relief, typically defined as ≥50% pain reduction from baseline. Additional outcomes included functional disability, quality of life measures, patient satisfaction, and adverse events, though the specific outcomes and follow-up durations varied across trials.

## Meta-analysis results

### Pain Reduction

Our primary outcomes were the proportion of patients achieving ≥50% reduction in overall pain, back pain and leg pain intensity at the end of follow-up. Seven RCTs (n=334 patients) reported back pain responder rates, demonstrating that 45% (95% CI: 18-75%) achieved clinically meaningful relief, though with substantial heterogeneity (I² = 92.2%, τ² = 2.80, p < 0.0001). In contrast, leg pain showed more consistent results across three RCTs (n=94), with 55% achieving ≥50% reduction (95% CI: 45-65%, I² = 0%, p = 0.60). Overall pain responder rates across all trials were 56% (95% CI: 30-78%, I² = 92.9%, τ² = 2.35, p < 0.0001). Meta-regression analysis identified significant moderators of treatment response. Longer pain duration was significantly associated with higher responder rates for both overall pain reduction (β = 0.43, p < 0.001, R² = 64.2%) and back pain reduction (β = 0.44, p = 0.038, R² = 47.5%). Similarly, older patient age was associated with higher overall responder rates (β = 0.23, p = 0.003, R² = 55.8%), indicating that older patients and those with a longer history of pain were more likely to achieve ≥50% pain relief. Conversely, higher baseline leg pain scores were a negative predictor, associated with lower responder rates for both overall pain (β = −0.12, p < 0.001, R² = 89.5%) and back pain (β = −0.14, p = 0.007, R² = 69.4%).

Subgroup analyses revealed significant differences in treatment response based on stimulation parameters for back pain and overall pain outcomes. For back pain, high-frequency SCS (≥1 kHz) achieved markedly higher response rates than conventional low-frequency stimulation (92% [95% CI: 79-98%] vs. 28% [95% CI: 13-49%], p < 0.001) (Figure 1). When analyzing overall pain reduction across all studies, similar patterns emerged high-frequency SCS showed superior efficacy compared to conventional frequencies (92% [95% CI: 84-96%] vs. 31% [95% CI: 18-47%], p < 0.0001). Our subgroup analysis of leg pain responder rates showed no significant differences across stimulation modalities, with consistent response rates of approximately 55% regardless of waveform type (burst 60% vs. tonic 51%, p = 0.36) (Figure 2).

**Figure 1.**
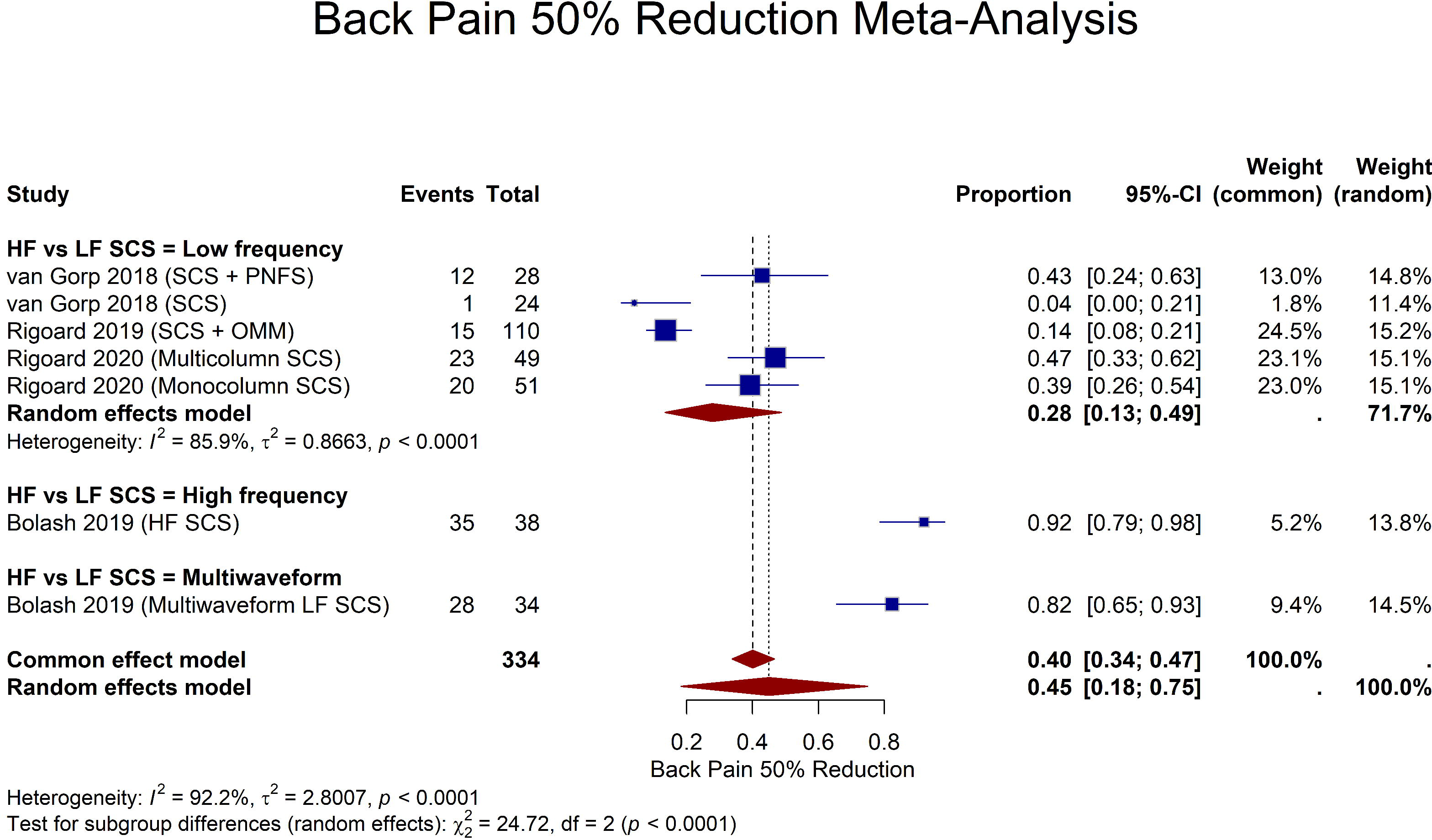
Forest plot of back pain responder rates (≥50% reduction) stratified by stimulation frequency.

**Figure 2.**
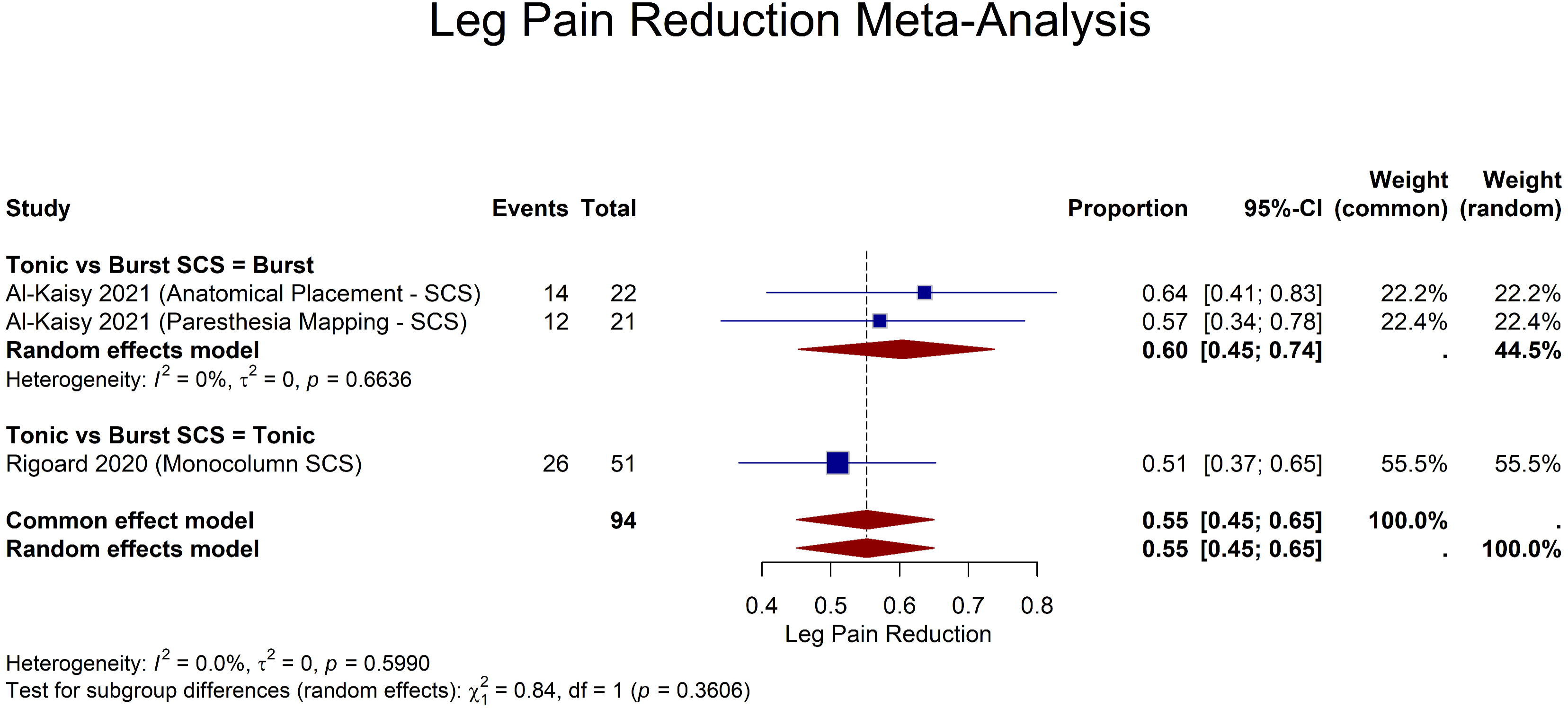
Forest plot of leg pain responder rates (≥50% reduction) stratified by stimulation modality.

**Figure 3.**
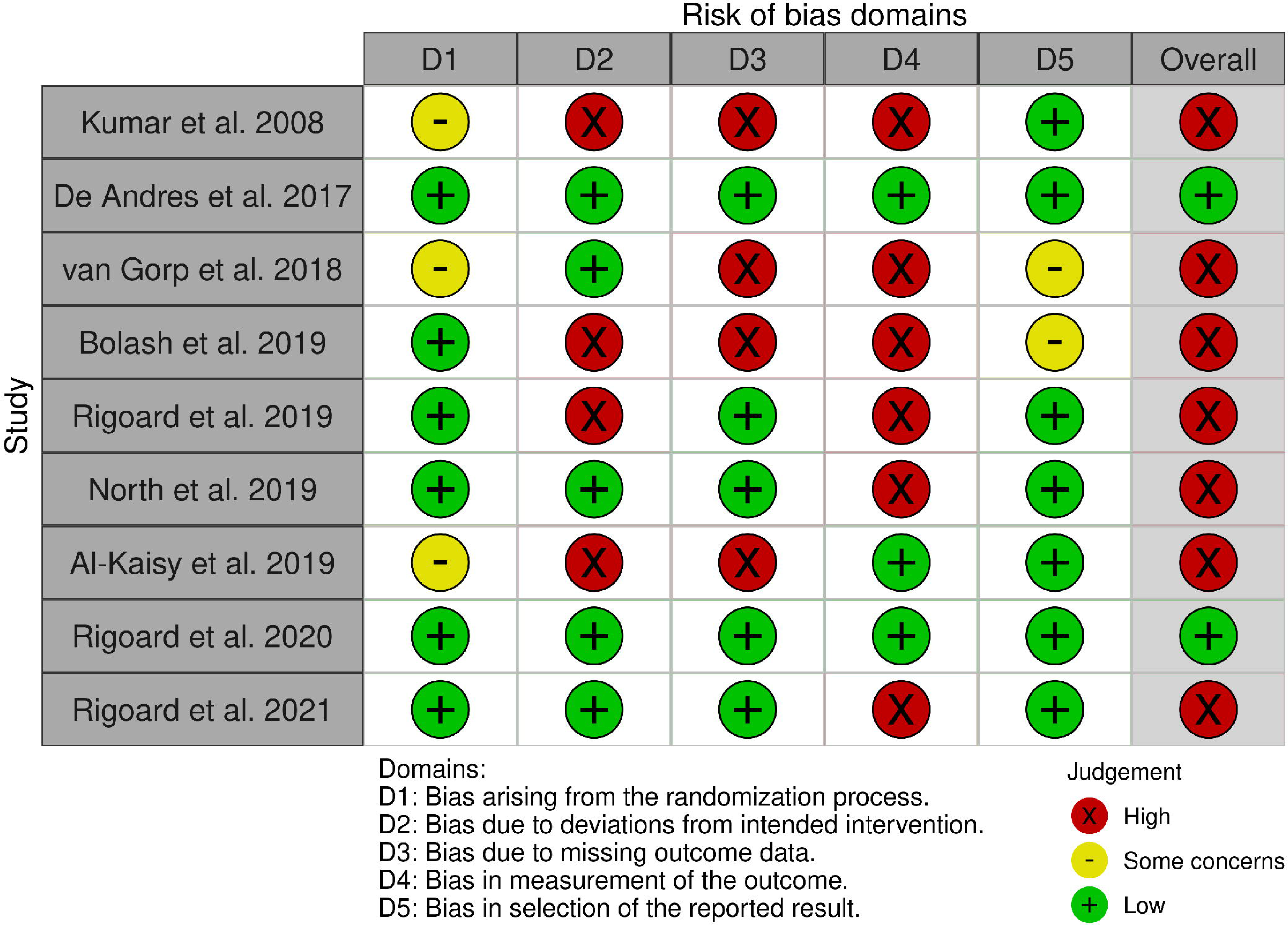
Traffic lights plot for Risk of bias assessment for included randomized controlled trials using the Cochrane Risk of Bias 2 tool.

### Pain Intensity and Functional Outcomes

Pain intensity demonstrated substantial reductions from baseline across all measured domains. For back pain, mean VAS scores decreased from a baseline of 73.03 to 41.67 (95% CI: 36.12-47.22, I²=22.8%) at 6 months and remained stable at 35.66 (95% CI: 25.39-45.93, I²=75.0%) at 12 months. Leg pain showed more pronounced improvement, with VAS scores declining from a baseline of 61.81 to 23.75 (95% CI: 17.69-29.81, I²=78.8%) at 6 months and 29.16 (95% CI: 24.81-33.52, I²=0%) at 12 months. Functional disability, measured by the Oswestry Disability Index, improved from a baseline of 51.75 to 34.13 (95% CI: 26.75-41.50, I²=89.9%) at 6 months and 30.07 (95% CI: 22.98-37.17, I²=88.7%) at 12 months.

Subgroup analyses revealed varying effects of stimulation parameters on pain and functional outcomes across different timepoints. For back pain, burst stimulation demonstrated significantly lower VAS scores compared to tonic stimulation at the 3rd month (25.86 [95% CI: 19.02–32.71] vs. 55.18 [95% CI: 29.01–81.34], p=0.034) and 12th month (27.52 [95% CI: 19.91–35.13] vs. 43.47 [95% CI: 33.69–53.26], p=0.011). However, at the 6th month, while burst stimulation scores remained lower, the difference did not reach statistical significance (35.06 vs. 45.22, p=0.053). Consistent with previous findings, SCS with PNFS was associated with higher 12th month Back Pain VAS score compared to SCS alone interventions (46.90 vs. 28.52, p<0.001), reflecting baseline group differences in the included study.

For leg pain at 6 months, high-frequency stimulation yielded superior outcomes compared to low-frequency stimulation, with significantly lower mean VAS scores (13.30 [95% CI: 8.82–17.78] vs. 28.42 [95% CI: 24.02–32.88], p<0.001).

Analysis of functional disability at 6 months identified significant differences based on frequency and intervention type. High-frequency stimulation was associated with significantly lower ODI scores compared to low-frequency stimulation (22.92 [95% CI: 20.39–25.45] vs. 36.15 [95% CI: 28.60–43.70], p=0.001). Complete subgroup analyses for all timepoints are provided in Table 2.

**Table 1.**
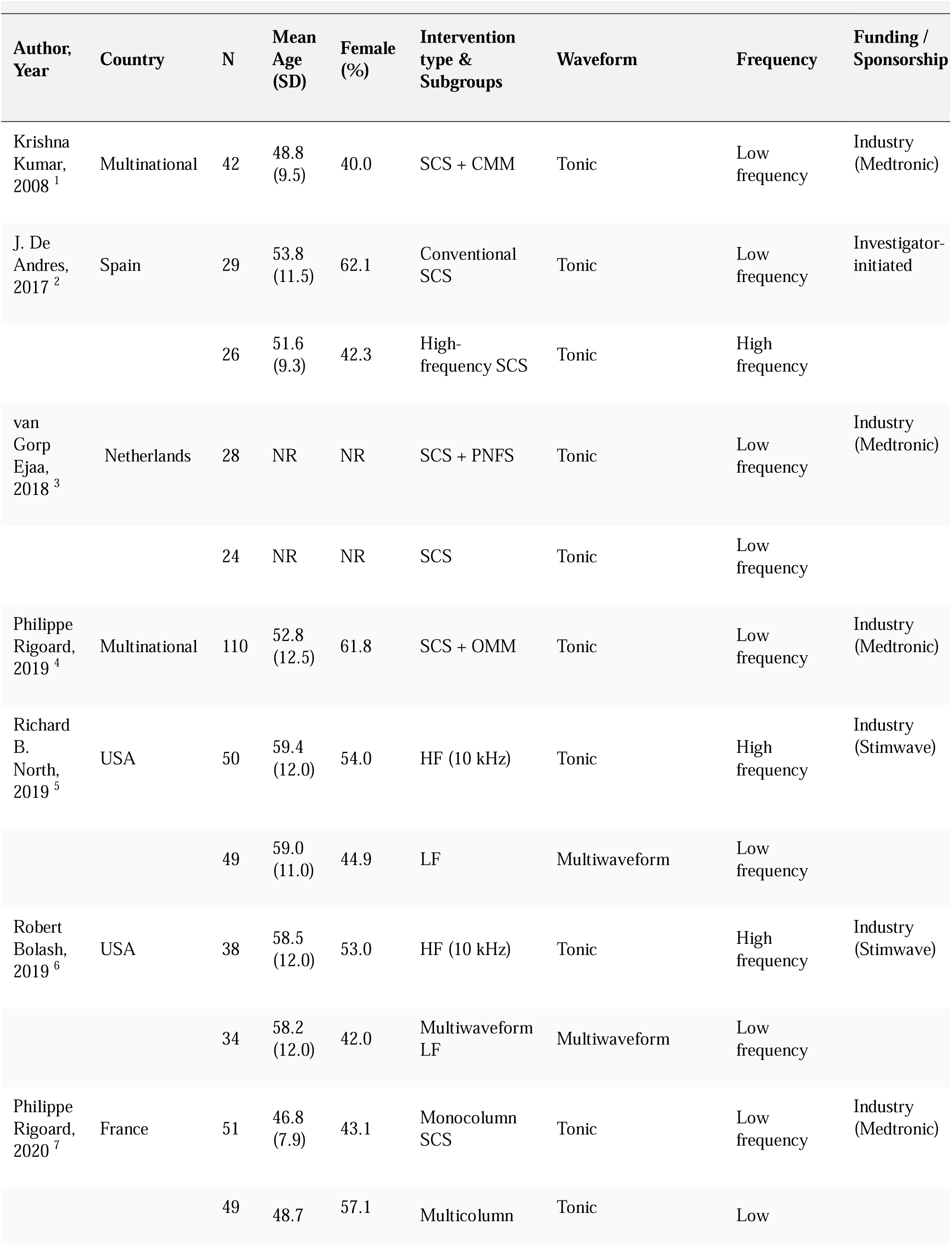
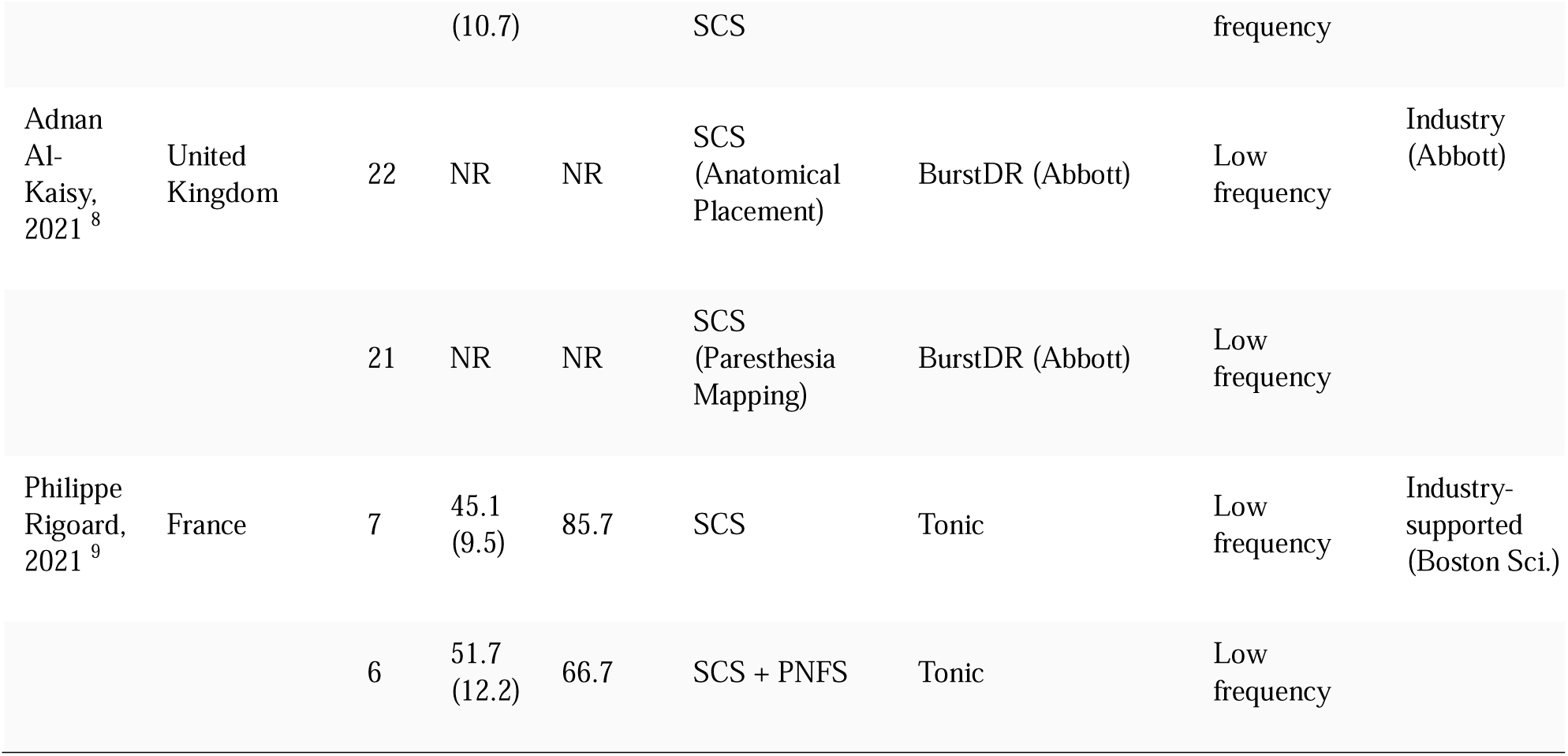
Study Characteristics.

| Author, Year | Country | N | Mean Age (SD) | Female (%) | Intervention type & Subgroups | Waveform | Frequency | Funding / Sponsorship |
| --- | --- | --- | --- | --- | --- | --- | --- | --- |

**Table 1. Study Characteristics**
| Author, Year | Country | N | Mean Age (SD) | Female (%) | Intervention type & Subgroups | Waveform | Frequency | Funding / Sponsorship |
| --- | --- | --- | --- | --- | --- | --- | --- | --- |
| Krishna Kumar, 2008 <sup>1</sup> | Multinational | 42 | 48.8 (9.5) | 40.0 | SCS + CMM | Tonic | Low frequency | Industry (Medtronic) |
| J. De Andres, 2017 <sup>2</sup> | Spain | 29 | 53.8 (11.5) | 62.1 | Conventional SCS | Tonic | Low frequency | Investigator-initiated |
|  |  | 26 | 51.6 (9.3) | 42.3 | High-frequency SCS | Tonic | High frequency |  |
| van Gorp Ejaa, 2018 <sup>3</sup> | Netherlands | 28 | NR | NR | SCS + PNFS | Tonic | Low frequency | Industry (Medtronic) |
|  |  | 24 | NR | NR | SCS | Tonic | Low frequency |  |
| Philippe Rigoard, 2019 <sup>4</sup> | Multinational | 110 | 52.8 (12.5) | 61.8 | SCS + OMM | Tonic | Low frequency | Industry (Medtronic) |
| Richard B. North, 2019 <sup>5</sup> | USA | 50 | 59.4 (12.0) | 54.0 | HF (10 kHz) | Tonic | High frequency | Industry (Stimwave) |
|  |  | 49 | 59.0 (11.0) | 44.9 | LF | Multiwaveform | Low frequency |  |
| Robert Bolash, 2019 <sup>6</sup> | USA | 38 | 58.5 (12.0) | 53.0 | HF (10 kHz) | Tonic | High frequency | Industry (Stimwave) |
|  |  | 34 | 58.2 (12.0) | 42.0 | Multiwaveform LF | Multiwaveform | Low frequency |  |
| Philippe Rigoard, 2020 <sup>7</sup> | France | 51 | 46.8 (7.9) | 43.1 | Monocolumn SCS | Tonic | Low frequency | Industry (Medtronic) |
|  |  | 49 | 48.7 | 57.1 | Multicolumn | Tonic | Low |  |

**Table 1. Study Characteristics**
| Author, Year | Country | N | Mean Age (SD) | Female (%) | Intervention type & Subgroups | Waveform | Frequency | Funding / Sponsorship |
| --- | --- | --- | --- | --- | --- | --- | --- | --- |
|  |  |  | (10.7) |  | SCS |  | frequency |  |
| Adnan Al-Kaisy, 2021<sup>8</sup> | United Kingdom | 22 | NR | NR | SCS (Anatomical Placement) | BurstDR (Abbott) | Low frequency | Industry (Abbott) |
|  |  | 21 | NR | NR | SCS (Paresthesia Mapping) | BurstDR (Abbott) | Low frequency |  |
| Philippe Rigoard, 2021<sup>9</sup> | France | 7 | 45.1 (9.5) | 85.7 | SCS | Tonic | Low frequency | Industry-supported (Boston Sci.) |
|  |  | 6 | 51.7 (12.2) | 66.7 | SCS + PNFS | Tonic | Low frequency |  |

**Table 2.**
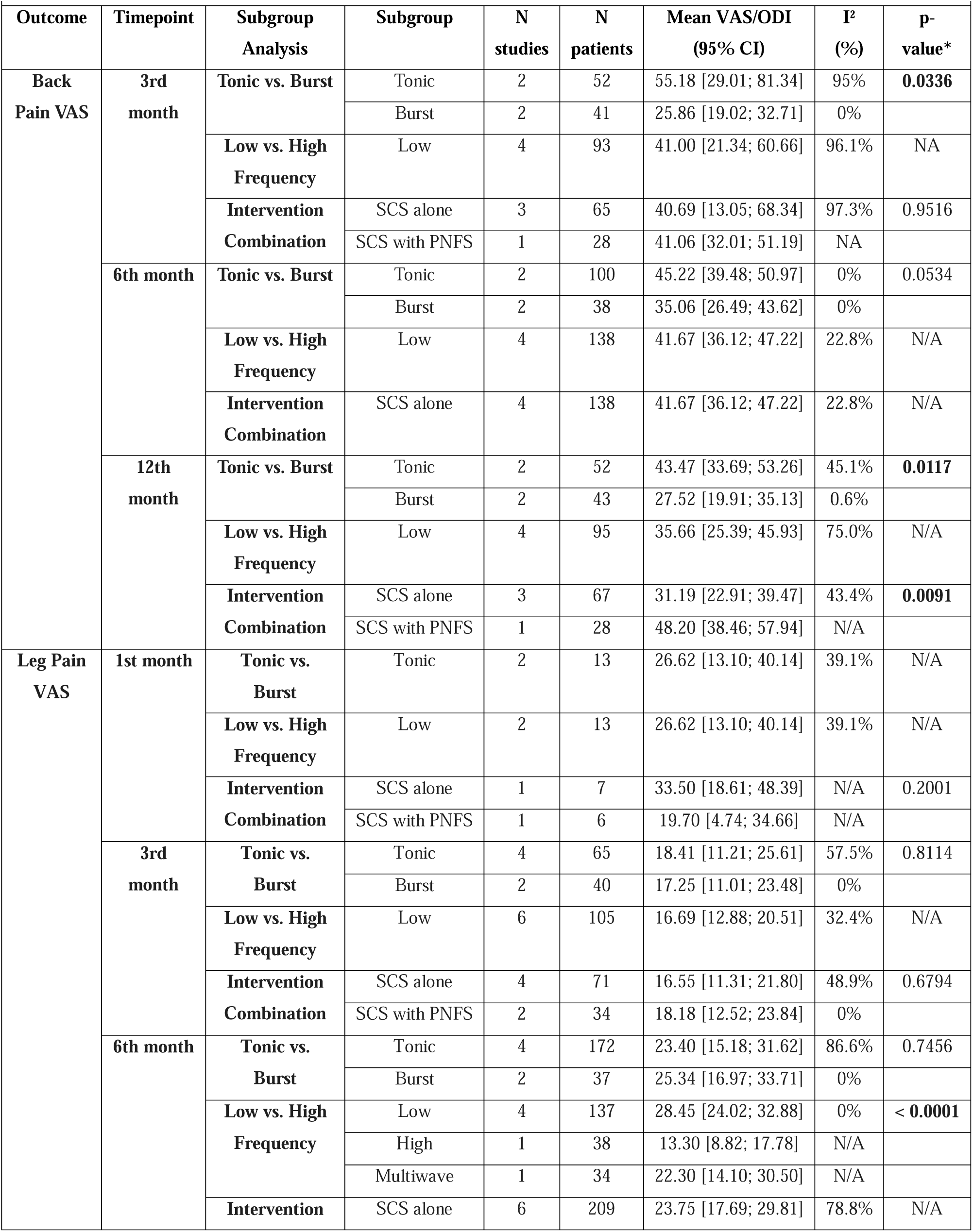
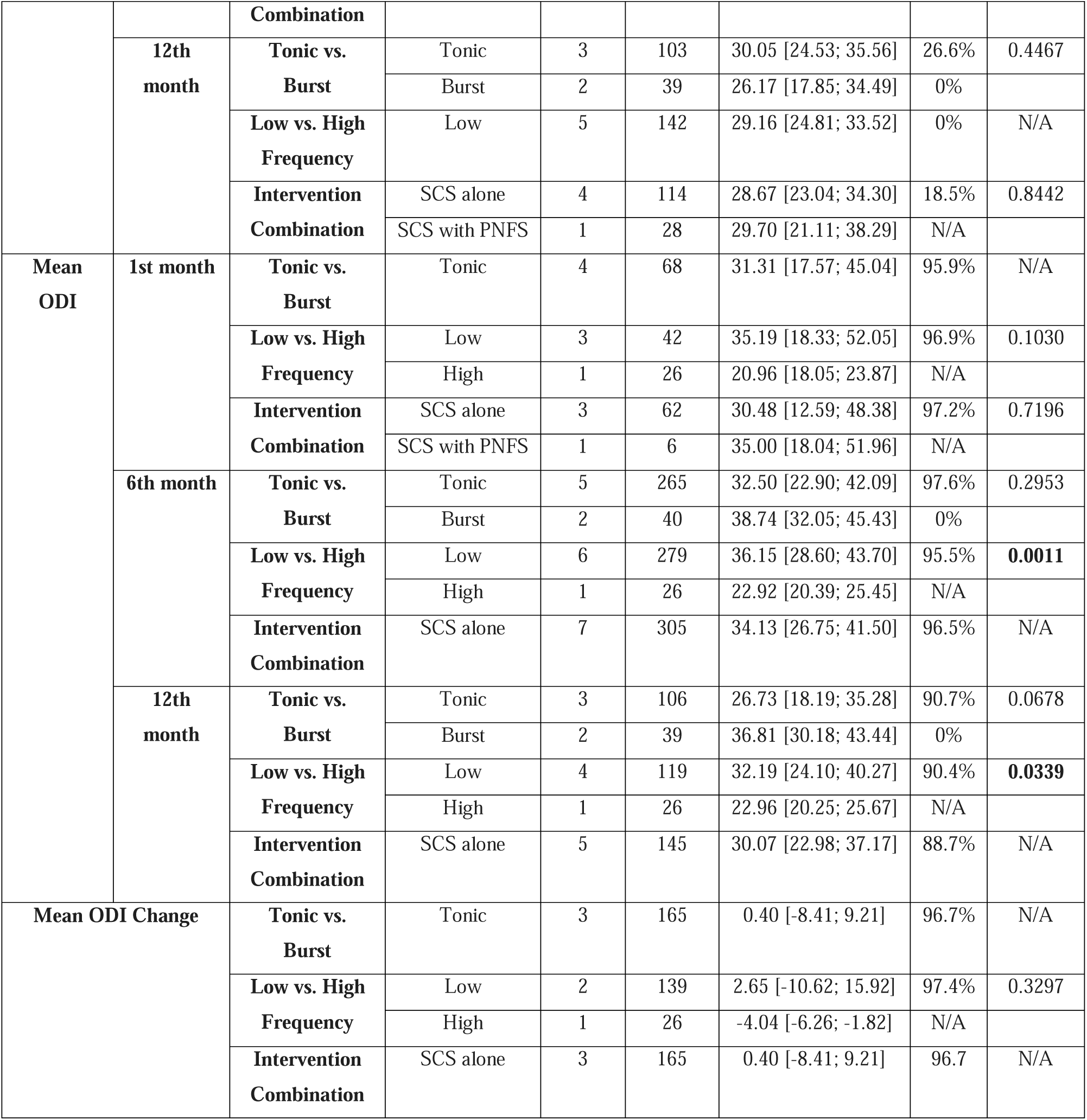
Subgroup for mean VAS/ODI in different timepoints.

### Safety and Tolerability Outcomes

Serious adverse events occurred in 10% of patients (95% CI: 5-18%, 5 studies, n=237, I²=40.5%). Hardware failure in 5% (95% CI: 2-11%, 5 studies, n=169, I²=0%), and lead fracture in 5% (95% CI: 2-11%, 5 studies, n=169, I²=0%). Procedural complications were infrequent, with infection occurring in 6% (95% CI: 4-9%, 8 studies, n=379, I²=0%) and hematoma in 2% (95% CI: 0-6%, 4 studies, n=155, I²=0%). Device explanation was required in 4% of patients (95% CI: 2-9%, 4 studies, n=155, I²=0%). Revision surgery was necessary in 16% of cases (95% CI: 8-29%, 4 studies, n=207, I²=71.0%), with substantial heterogeneity across studies. Patient-reported tolerability outcomes demonstrated high treatment satisfaction, with 91% of patients reporting satisfaction with their therapy (95% CI: 82-96%, I²=80.8%). Pocket pain was reported by 4% of patients (95% CI: 1-12%, 5 studies, n=237, I²=57.9%), and uncomfortable stimulation affected 4% (95% CI: 1-12%, 4 studies, n=127, I²=4.1%). Complete safety plots are in the Supplementary file.

### Sensitivity Analyses

Leave-one-out sensitivity analyses demonstrated that pooled effect estimates remained largely stable across most primary and secondary outcomes. For the primary outcome of back pain responder rates, omitting individual studies resulted in pooled proportions ranging from 36% to 54% (overall: 45%), with persistent high heterogeneity (I²: 88–94%). Leg pain responder rates showed consistent estimates (range: 53–60%, overall: 55%) with homogeneous results (I²: 0%) regardless of study exclusion. Pain intensity outcomes at 6 months remained robust. Back pain VAS estimates ranged from 40 to 44 mm (overall: 42 mm) with generally low to moderate heterogeneity (I²: 0–44%). In contrast, leg pain VAS estimates showed greater variability, ranging from 19 to 27 mm (overall: 21 mm), where omitting Bolash et al. (2019) notably reduced heterogeneity from 83% to 0%.

Functional disability outcomes demonstrated stability at 6 months (ODI range: 27–35, overall: 31) and 12 months (ODI range: 24–27, overall: 25), though heterogeneity remained high (I² > 80%) across all analyses. For safety outcomes, exclusion of individual studies did not materially alter pooled estimates for infection (range: 4.6–6.2%; I²: 0%), hardware failure (range: 3.3–6.5%; I²: 0–20%), or hematoma rates (range: 1.4–1.9%; I²: 0%). However, revision surgery rates were sensitive to the exclusion of Kumar et al. (2008), which reduced the pooled rate from 16% to 12% and significantly dropped heterogeneity from 80% to 3.5%. Similarly, satisfaction rates remained high (range: 90–94%), with heterogeneity dropping to 0% when excluding Al-Kaisy et al. (2021). These sensitivity analyses confirm the robustness of our findings, with most outcomes showing stable estimates, though specific studies were identified as primary drivers of heterogeneity for leg pain intensity and revision surgery rates.

### Publication Bias

Assessment of publication bias using trim-and-fill analysis and Egger’s regression test revealed evidence of small-study effects in several outcomes. Egger’s test demonstrated significant asymmetry for multiple safety outcomes, including explantation (p=0.0219), hardware failure (p=0.0007), infection (p=0.0015), lead fracture (p=0.0007), and uncomfortable stimulation (p=0.0061). Among efficacy outcomes, significant asymmetry was detected for leg pain VAS at 3 months (p=0.0162) and ODI at 12 months (p=0.0492), while primary responder rates and most other VAS timepoints showed no significant asymmetry.

Trim-and-fill analysis corroborated these findings, requiring study imputations for outcomes showing Egger’s test significance. For safety outcomes, infection rates required the imputation of 3 studies, increasing the adjusted pooled estimate to 6.5% (from 5.8%). Similarly, pocket pain rates required 3 imputations (adjusted: 10.3%), while explantation, hardware failure, lead fracture, and uncomfortable stimulation each required 2 imputations to correct for asymmetry.

For efficacy outcomes, leg pain VAS at 3 months required 3 imputations, resulting in an adjusted estimate of 14.9 mm (vs. original 15.6 mm). ODI at 12 months required 2 imputations, slightly adjusting the pooled mean to 24.3 (from 25.1). Other outcomes, including back pain responder rates and long-term pain intensity scores, remained robust without requiring significant adjustment.

Beyond the statistical evidence of small-study effects, the funding landscape of the included trials suggests an additional layer of potential bias. Of the 9 RCTs analyzed, 8 of them were directly sponsored, funded, or initiated by medical device manufacturers. This is a critical observation, as industry-sponsored studies in neuromodulation tend to report a greater overall treatment effect and higher responder rates compared to investigator-initiated studies. This ‘sponsorship effect’ is exemplified by the fact that the only purely independent trial in our review (De Andres et al. 2017^22^) yielded more conservative results than its industry-funded counterparts. The predominance of manufacturer-initiated research, combined with the findings of the trim-and-fill analysis, suggests that the pooled efficacy estimates—particularly for newer waveforms—may be subject to an overestimation of clinical benefit Risk of bias assessment Risk of bias assessment using the Cochrane Risk of Bias 2 tool revealed that only two studies (Rigoard et al. 2020 and De Andres et al. 2017) demonstrated low risk of bias across all domains, with the remaining nine studies rated as having overall high risk of bias (Figure 4). Domain-specific analysis identified systematic concerns across multiple domains. For randomization (D1), six studies showed low risk while three had some concerns. Deviations from intended interventions (D2) represented the most problematic domain, with four studies demonstrating high risk of bias, primarily due to the inherent difficulty of blinding participants and personnel in neuromodulation trials where paresthesia presence may unmask treatment allocation. Missing outcome data (D3) posed concerns in four studies rated as high risk, reflecting substantial loss to follow-up or incomplete outcome reporting. Measurement of outcomes (D4) was compromised in six studies due to the unblinded nature of pain assessment in most trials, with three studies achieving low risk through adequate blinding of outcome assessors. Selection of reported results (D5) showed better performance, with seven studies at low risk and two with some concerns. The predominance of high overall risk of bias, particularly in domains D2 and D4, reflects the methodological challenges inherent to SCS trials where effective blinding is difficult to achieve and maintain.

## Discussion

Our systematic review and meta-analysis demonstrates that spinal cord stimulation provides clinically meaningful pain relief in PSPS-T2, with efficacy dependent on both stimulation modality and pain location. Our main finding is that, in comparison to traditional tonic stimulation, high-frequency SCS achieves a significantly higher responder rate for back pain (92% versus 28%, p<0.001). Furthermore, while pooled dichotomous responder rates for burst stimulation were constrained by data availability following rigorous trial exclusion, burst waveforms demonstrated significantly superior continuous back pain relief at both three and twelve months compared to conventional tonic paradigms. On the other hand, leg pain responses were similar across modalities; regardless of waveform type, approximately 55% of patients experienced a ≥50% reduction in pain.

### The Axial Gap: Why Newer Waveforms Win for Back Pain

The superiority of burst and high-frequency SCS for axial back pain addresses a longstanding therapeutic limitation. Conventional tonic stimulation, while effective for radicular leg pain with responder rates of 50-60% ^23,24^, has historically struggled to provide adequate coverage of axial low back pain without uncomfortable paresthesia ^4^. This discrepancy is supported by our meta-analysis, which demonstrates that conventional tonic stimulation achieved only a 28% response rate for back pain, whereas high-frequency modalities achieved a profound 92% response rate. The clinical superiority of sub-perception waveforms extends to continuous metrics, with burst stimulation yielding significantly lower back pain VAS scores than tonic stimulation at one-year follow-up. This disparity in efficacy most likely results from fundamental variations in the activation, spatial targeting, and modulation of the dorsal column and deeper dorsal horn circuitry. whereas high-frequency modalities achieved a profound 92% response rate. This exceptionally high response profile aligns tightly with recent real-world pragmatic trials demonstrating profound and sustained axial coverage utilizing 10 kHz paradigms without the requirement for overlapping paresthesia ^25^, an efficacy further validated by contemporary independent meta-analyses confirming superior long-term functional recovery ^26^.

Rigoard et al. demonstrated in the PROMISE study that multicolumn lead arrays combined with optimized field shaping enable superior dorsal horn coverage, potentially explaining improved back pain outcomes ^27^. The technical advances enabling paresthesia-free stimulation appear particularly critical for axial pain, as conventional tonic stimulation at intensities sufficient to cover low back dermatomes frequently produces uncomfortable sensations that limit therapeutic efficacy ^22^. Al-Kaisy et al. demonstrated in their frequency-optimization trial that 5882 Hz achieved significantly lower back pain scores (3.22 on VAS) compared to lower frequencies and sham stimulation (4.83 for sham, p=0.002), supporting the frequency-dependent nature of axial pain relief ^28^.

### Leg Pain: The Durability of Tonic Stimulation

Our analysis revealed no significant differences in leg pain responder rates between stimulation modalities; tonic achieved 51% versus 60% for burst (p=0.36). The established efficacy of traditional tonic SCS for radicular pain syndromes is reflected in this equivalency across waveforms, which stands in stark contrast to back pain outcomes. With 48% of responders at 24 months, the PROCESS trial demonstrated the effectiveness of tonic SCS for predominant leg pain ^24^. With consistent ∼55% responder rates across all modalities, our pooled analysis validates these historical benchmarks.

While back pain predominance requires consideration of burst or high-frequency alternatives, tonic SCS is still a validated and cost-effective option for patients with leg-dominant pain ^29^. Both clinical results and the use of healthcare resources are optimized by this phenotype-dependent approach to modality selection.

### Mechanisms of Action: Paresthesia versus Paresthesia-Free

The divergent outcomes between back and leg pain across stimulation modalities likely reflect distinct neurophysiological mechanisms. Conventional tonic SCS is thought to operate via the Gate Control Theory, producing paresthesia through dorsal column activation that masks nociceptive signals ^30,31^, while burst and high-frequency stimulation is thought to function through sub-threshold, producing analgesia without conscious sensory perception ^32,33^. Al-Kaisy et al. demonstrated in the CRISP study that anatomical lead placement at T9-T10 produced equivalent pain relief to paresthesia-guided placement for burst SCS, with 81% of subjects achieving at least 50% back pain relief regardless of placement method ^14^. This finding suggests that paresthesia-free modalities may target deeper pain processing networks that do not require precise paresthesia overlap with painful areas.

The specific frequency parameters within paresthesia-free stimulation appear critical to clinical efficacy. Al-Kaisy et al. demonstrated in a sham-controlled crossover study that 5882 Hz stimulation produced significantly greater axial low back pain relief compared to lower frequencies (1200 Hz, 3030 Hz) and sham stimulation, with sham producing similar effects to lower frequencies revealing substantial placebo effects that may confound interpretation of earlier open-label trials ^28^. Vesper et al. similarly found burst stimulation significantly outperformed both 500 Hz tonic stimulation and placebo at 2-year follow-up ^29^. These mechanistic distinctions carry practical implications: Van Roosendaal et al. quantified that higher frequency and burst patterns require greater energy consumption than conventional tonic stimulation, imposing greater recharging burden that influences real-world treatment adherence^34^.

### Addressing Heterogeneity and Conflicting Evidence

The substantial heterogeneity observed in our back pain responder analysis demands critical examination. Multiple factors contribute to this variability: heterogeneous definitions of treatment response, varying follow-up durations (6 versus 12 months), differences in lead configurations, and inconsistent programming protocols across trials. More fundamentally, the distinction between SCS-naïve patients and those receiving salvage therapy after failed conventional stimulation introduces selection bias that substantially influences reported outcomes ^35^. To address these sources of heterogeneity, we conducted meta-regression analysis, which identified significant moderators of treatment response. Longer pain duration was associated with higher responder rates, while younger patient age predicted lower pain intensity at 6-month follow-up.

The placebo effect in neuromodulation trials presents another critical consideration. Hara et al. reported no significant difference between burst stimulation and placebo in their rigorously-controlled trial ^18^, contradicting earlier open-label studies by Schu et al. that demonstrated robust burst superiority ^36^. It’s important to talk about the bigger question of how well SCS works. Two recent Cochrane systematic reviews have made some people worry about the evidence foundation for neuromodulation. O’Connell et al. ^37^ have done a review of 15 randomized trials on spinal neuromodulation for various chronic pain conditions, their conclusion was that the evidence for the efficacy of spinal cord stimulation (SCS) compared to placebo was not certain, with effect sizes falling below clinically significant thresholds. Traeger et al. ^38^ reviewed 13 trials using SCS for low back pain and concluded that it didn’t assist significantly, if at all, compared to a placebo. These reviews highlight a notable inconsistency in the literature, with open-label studies comparing SCS with conventional management consistently reporting substantial treatment effects, while placebo-controlled studies demonstrate considerably smaller, and at times non-significant, benefits. This trend suggests that placebo responses, patient expectations, and investigator bias may cause unblinded trial designs to exaggerate actual treatment effects. Note that blinded and unblinded studies were included in our meta-analysis. The efficacy estimates we provide might represent an upper limit of effect, and the true magnitude of modality-specific benefits might be smaller than our combined estimates indicate.

Our meta-regression identified older age and longer pain duration as unexpected positive predictors of therapeutic success. This may reflect age-related psychological adaptation, where older patients often report higher satisfaction with partial functional recovery compared to younger cohorts ^39^. Physiologically, prolonged chronicity might signal a stabilized central sensitization state amenable to neuromodulation, contrasting with the volatile inflammatory milieu of early PSPS-T2 ^40^. Conversely, severe baseline leg pain negatively predicted outcomes, suggesting that profound radiculopathy involves irreversible axonal damage or Wallerian degeneration that electrical modulation cannot override, necessitating early intervention ^41^.

### Safety Profile

Our pooled safety analysis revealed 10% serious adverse events, 6% infection, and 16% revision surgery rates. These complication frequencies align closely with established literature benchmarks. North et al. documented comparable revision surgery requirements at long-term follow-up ^23^. Importantly, high-frequency and burst SCS often require higher energy consumption than tonic stimulation, imposing greater recharging burden on patients—a “usability safety factor” that influences real-world treatment satisfaction ^27^. Van Roosendaal et al. quantified this difference, demonstrating that combination therapies increase total charge per second significantly, with implications for battery longevity and patient compliance ^34^.

### Future Directions: Combination and Personalization

The lack of leg pain superiority for newer waveforms, coupled with substantial heterogeneity in back pain outcomes strongly suggests that “one size does not fit all” in SCS therapy. Rigoard et al. in the MULTIWAVE study demonstrated that hybrid SCS systems allowing patients to toggle between tonic, burst, and high-frequency waveforms based on activity and pain patterns may represent the next paradigm in neuromodulation ^42^. This personalized approach acknowledges that optimal stimulation parameters likely vary not only between patients but within individual patients across time and context.

For specific pain locations such as groin or foot pain, dorsal root ganglion (DRG) stimulation may prove superior to conventional SCS ^43^. Future treatment algorithms should incorporate DRG stimulation as a complementary option rather than viewing SCS modalities as mutually exclusive. The integration of machine learning approaches to predict individual patient responses to specific waveforms based on pain phenotype, psychological factors, and neurophysiological testing represents an important frontier for precision neuromodulation ^44^.

### Limitations

Several limitations warrant consideration. First, the small sample size limits statistical power for subgroup comparisons and rare adverse events. Second, substantial heterogeneity reflects unmeasured differences in lead configurations, implantation techniques, and patient selection bias that our analyses could not fully explain. Third, the predominance of industry-sponsored trials and our trim-and-fill analysis suggests potential publication bias with overestimation of treatment benefits. Fourth, most trials compared SCS modalities against conventional medical management rather than direct head-to-head comparisons, limiting definitive ranking of modern waveforms. Fifth, follow-up duration remained ≤12 months in most trials, precluding assessment of long-term durability, despite evidence that SCS benefits may diminish over time. Finally, inconsistent responder definitions across trials (≥50%, ≥30%, or patient satisfaction) complicated pooled analyses and may have introduced classification bias.

### Conclusion

While high-frequency SCS represents a paradigm shift for back pain management in PSPS-T2, achieving an exceptional 92% responder rate compared to 28% for conventional tonic stimulation, and burst stimulation provides significantly superior continuous back pain relief, tonic SCS remains a robust standard for leg-dominant pain with 51-60% responder rates across modalities. Treatment recommendations should evolve beyond debates over “which waveform is best” toward patient-specific selection algorithms incorporating pain phenotype (axial versus radicular dominance), prior treatment history, cost considerations, and individual patient preferences regarding paresthesia. The future of SCS therapy lies not in universal protocols but in personalized, adaptive approaches that optimize outcomes for each patient’s unique pain presentation.

## Data Availability

The data that support the findings of this study are available from the corresponding author, M. Jakobs, upon reasonable request.

