## Supplementary for "Spinal Cord Stimulation for Persistent Spinal Pain Syndrome Type II: A Systematic Review and Subgroup Meta-analysis of Randomized Controlled Trials": Supplementary file.docx

| **Table S1.** The search strategy of electronic databases | | |
| --- | --- | --- |
| PubMed | ("failed back surgery syndrome"[MeSH Terms] OR "failed back surgery syndrome"[Title/Abstract] OR "FBSS"[Title/Abstract] OR "post-laminectomy syndrome"[Title/Abstract] OR "PSPS"[Title/Abstract] OR "Persistent spinal pain syndrome"[Title/Abstract])  AND  (("SCS"[Title/Abstract] OR "spinal cord stimulation"[Title/Abstract])  OR  ("PNS"[Title/Abstract] OR "peripheral nerve stimulation"[Title/Abstract])) | 609 |
| Embase | ('failed back surgery syndrome'/exp OR 'failed back surgery syndrome':ti,ab,kw OR 'fbss':ti,ab,kw OR 'post-laminectomy syndrome':ti,ab,kw OR 'psps':ti,ab,kw OR 'persistent spinal pain syndrome':ti,ab,kw) AND ('scs':ti,ab,kw OR 'spinal cord stimulation':ti,ab,kw OR 'pns':ti,ab,kw OR 'peripheral nerve stimulation':ti,ab,kw) | 878 |
| Scopus | ("failed back surgery syndrome" OR "failed back surgery syndrome" OR FBSS OR "post-laminectomy syndrome" OR PSPS OR "Persistent spinal pain syndrome") AND ((SCS OR "spinal cord stimulation") OR (PNS OR "peripheral nerve stimulation")) | 968 |

**Identification of studies via databases and registers**

Records removed before the screening:

Duplicate records removed (n =1132)

Records identified from*:

PubMed (n =609)

Embase (n =878)

Scopus (n =968)

**Identification**

Records excluded by title & abstract screening (n =951)

Records screened

(n =1323)

Records gone through full text screening (n =372)

No separate result for SCS (n = 33)

No separate result PSPS II (194)

**Screening**

Reports excluded:

Not RCT (123)

Data overlap (6)

Crossover (7)

Reports assessed for eligibility

(n = 145)

Studies included in the Meta analysis

(n =9)

**Included**

Figure S1. PRISMA

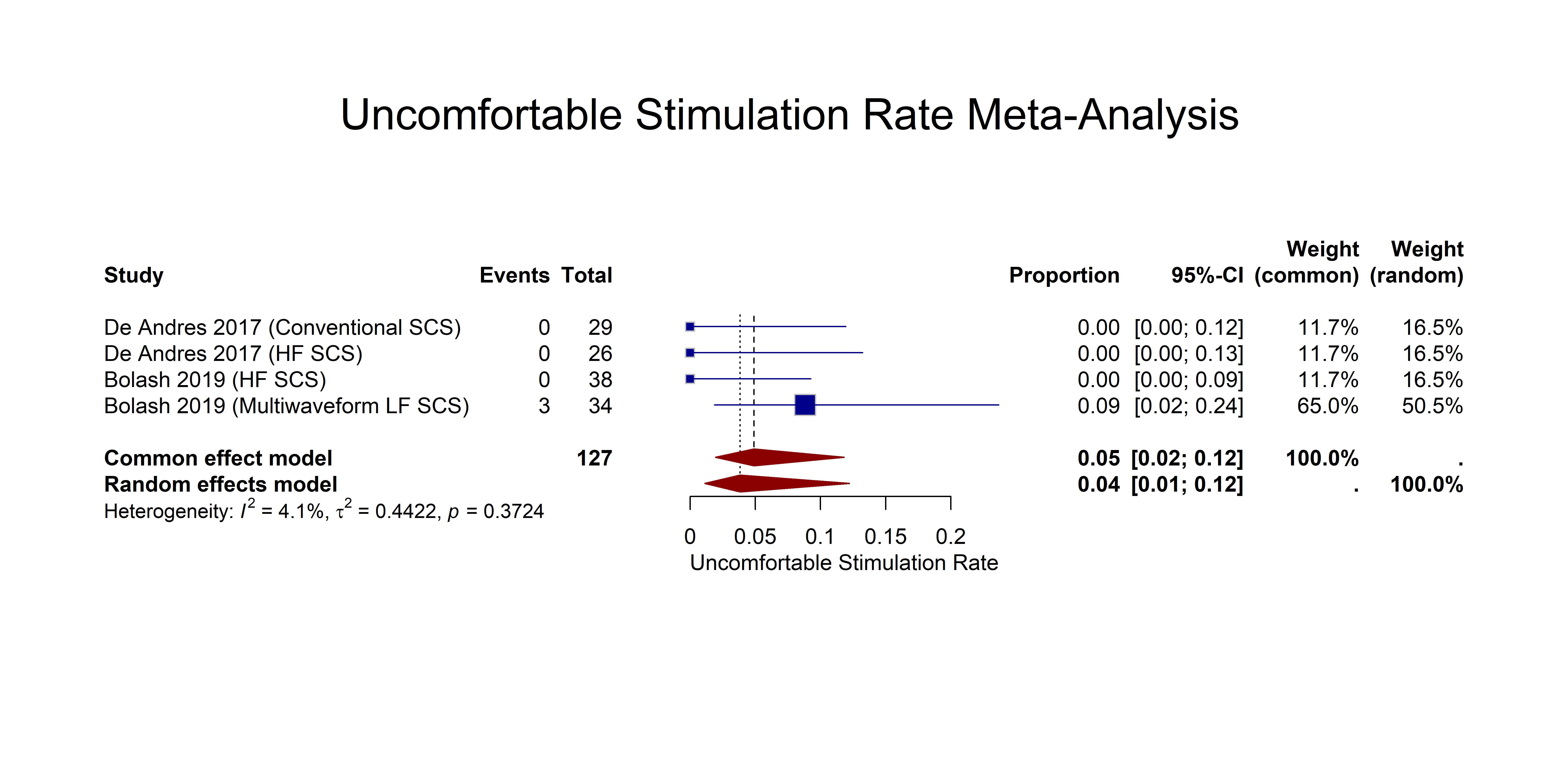

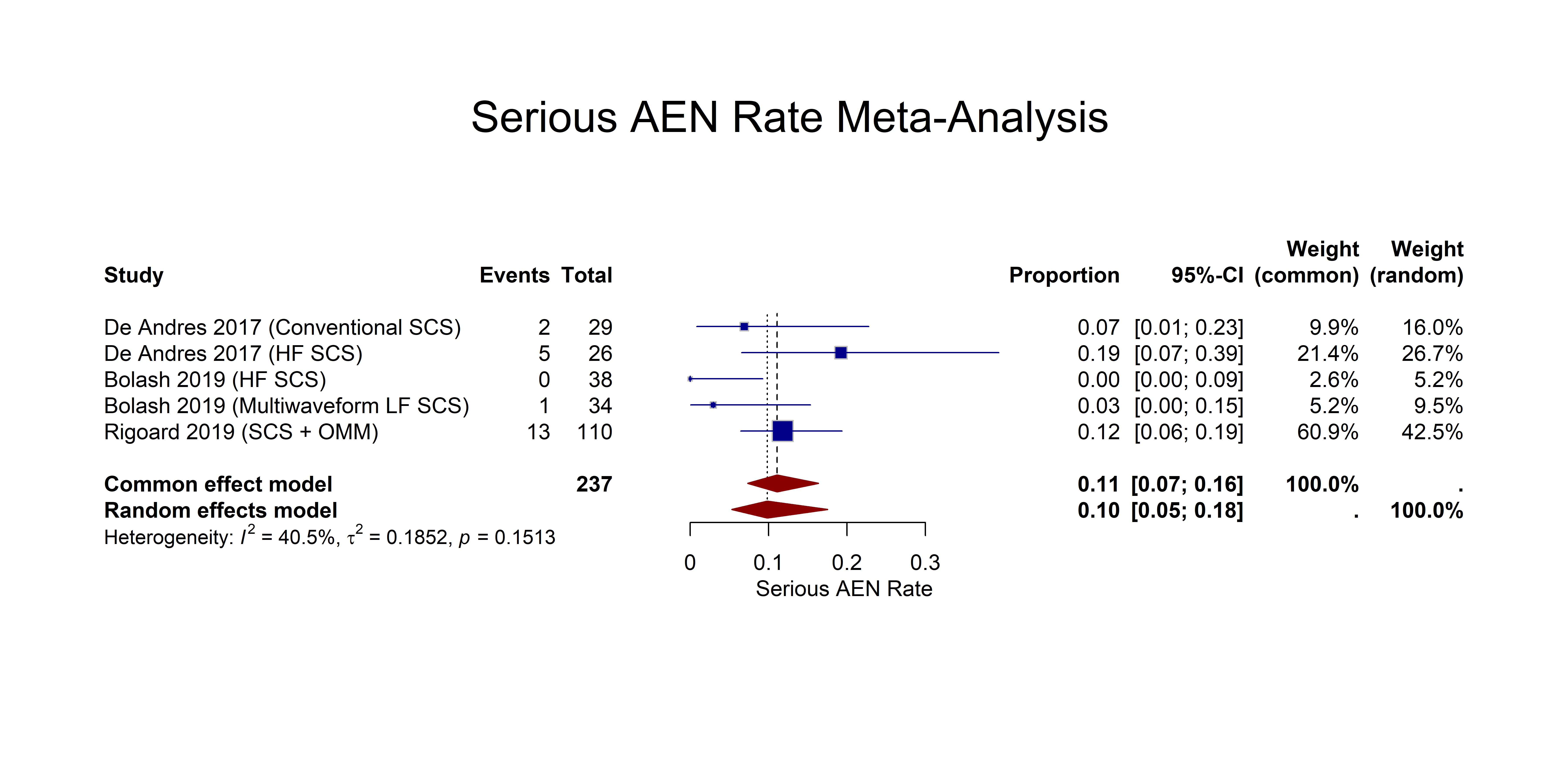

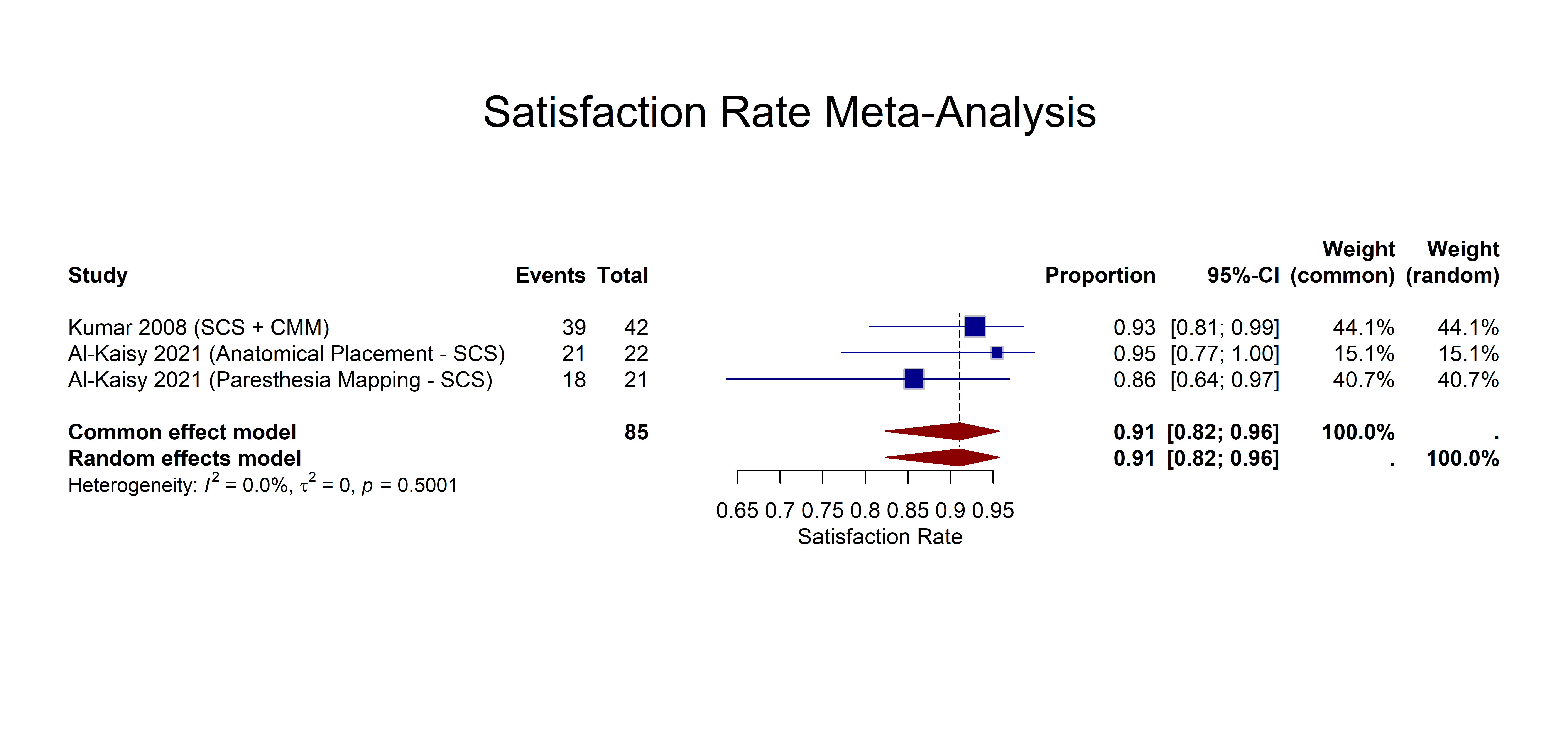

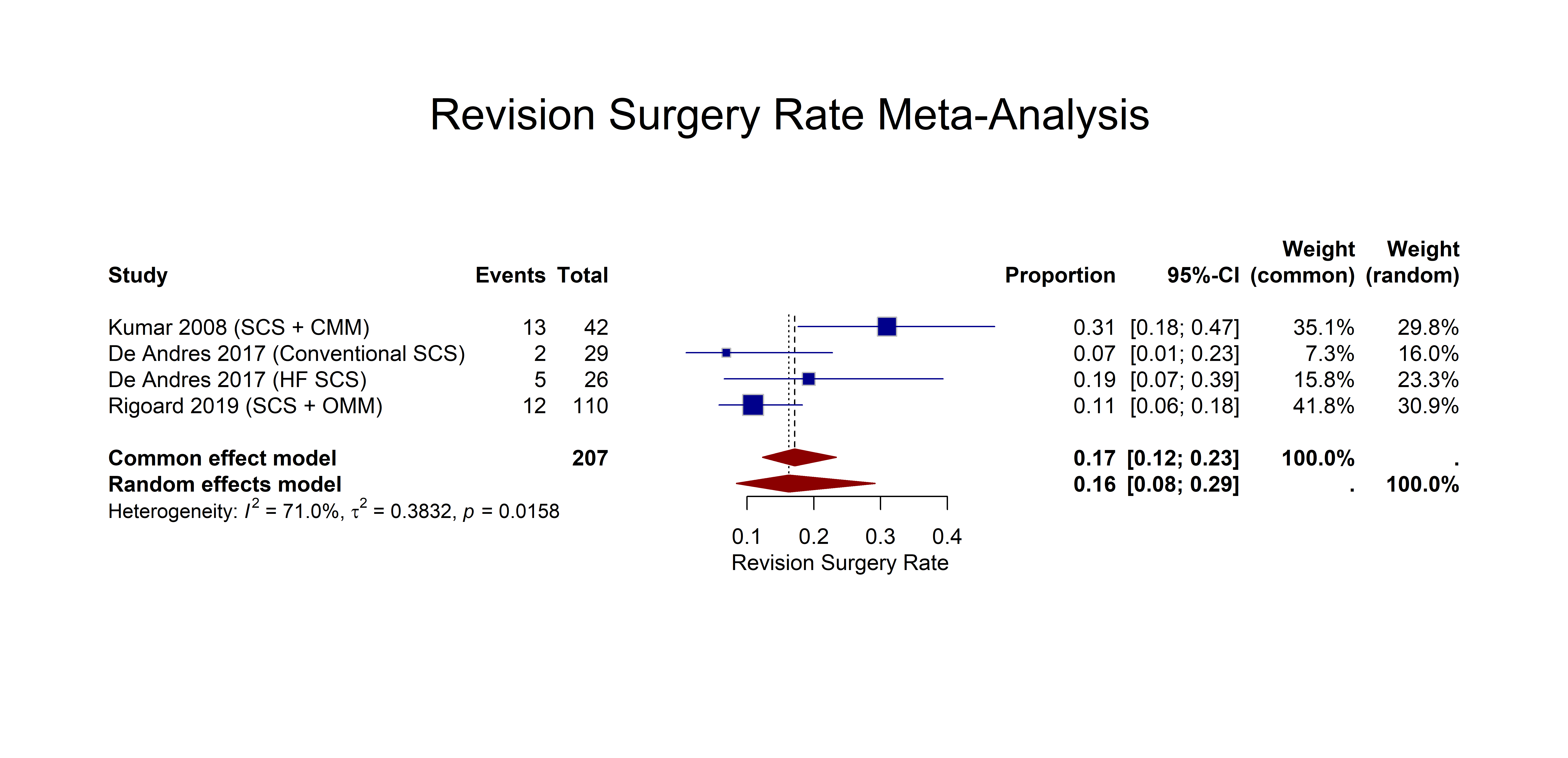

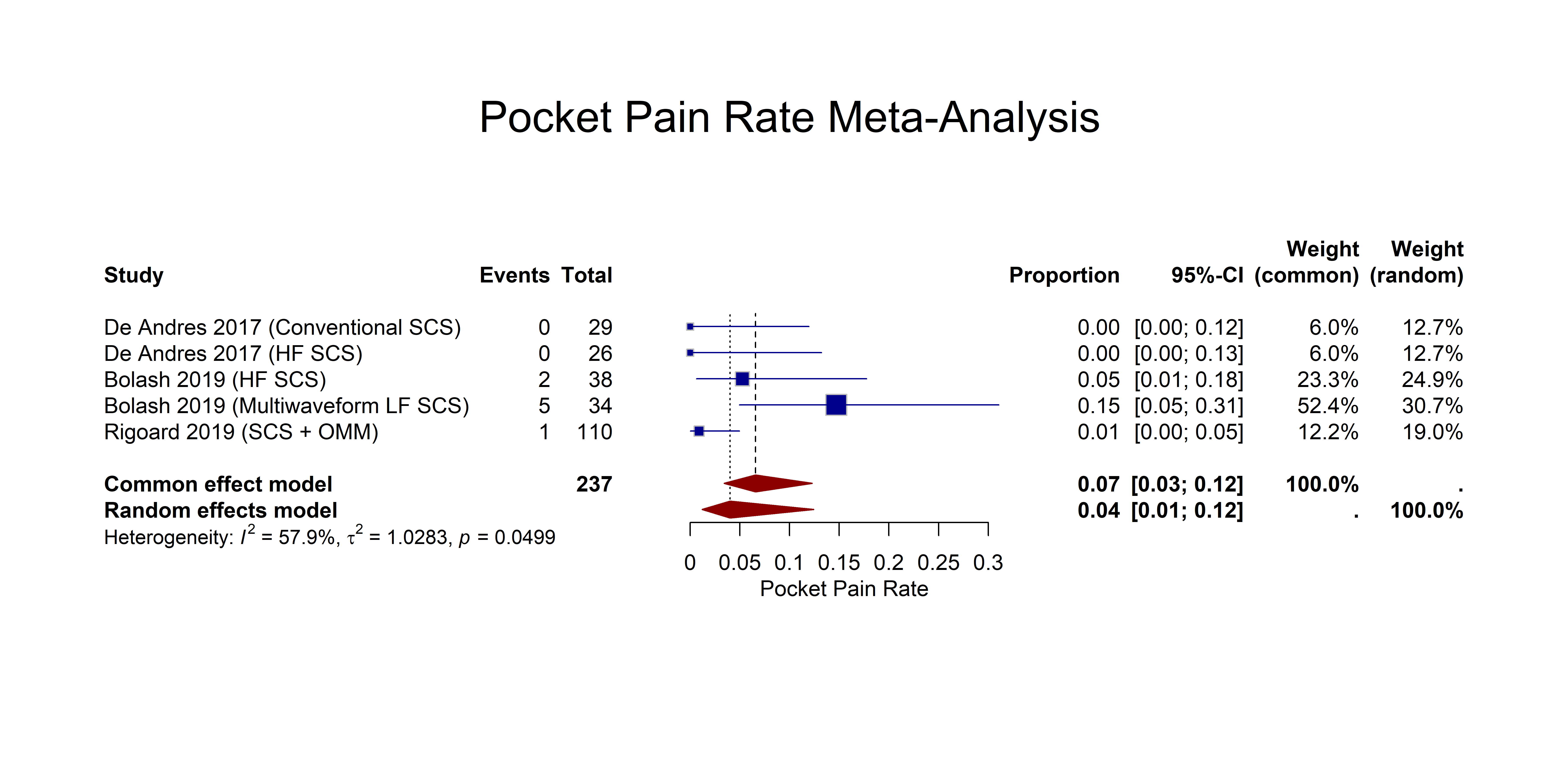

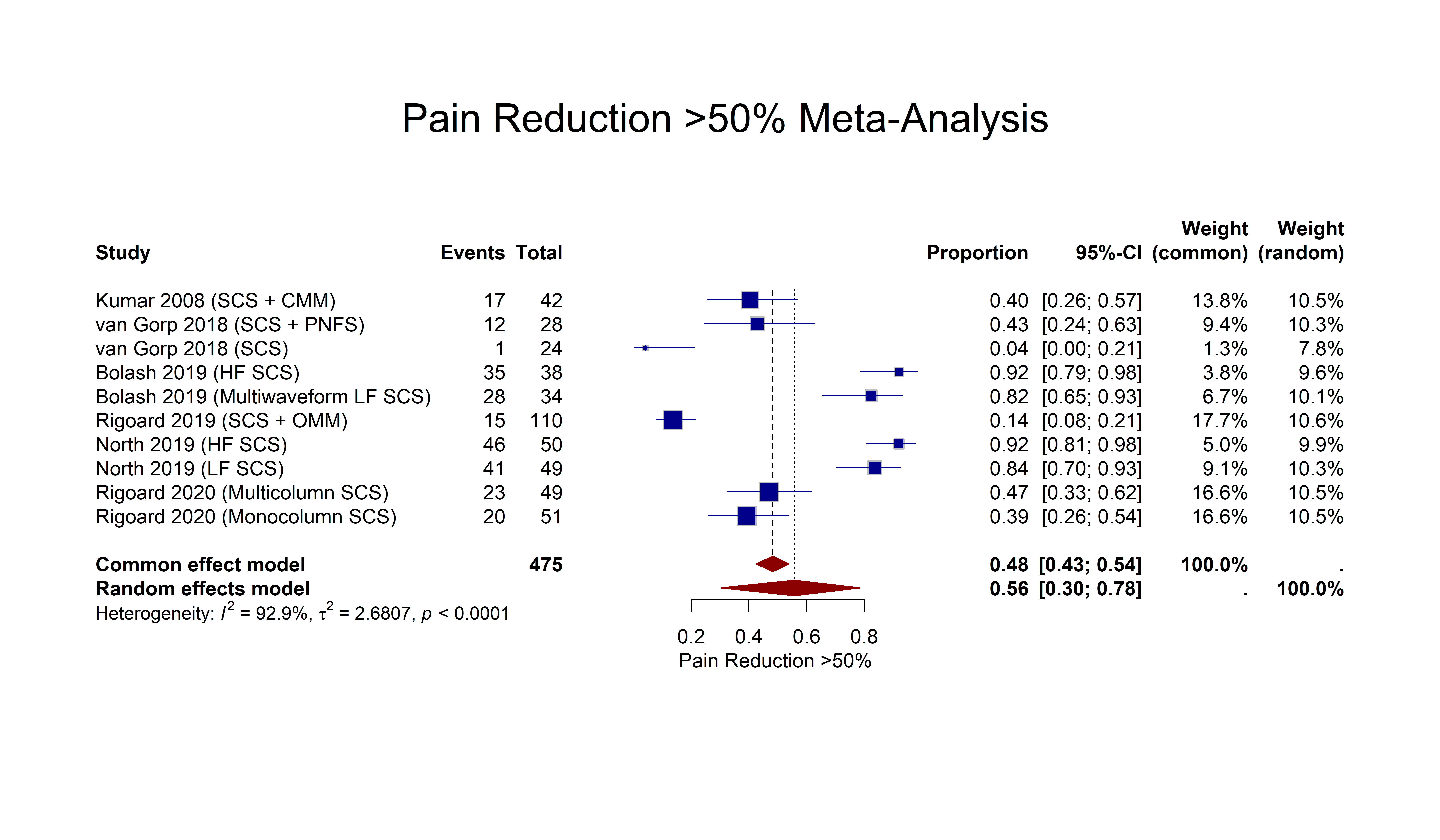

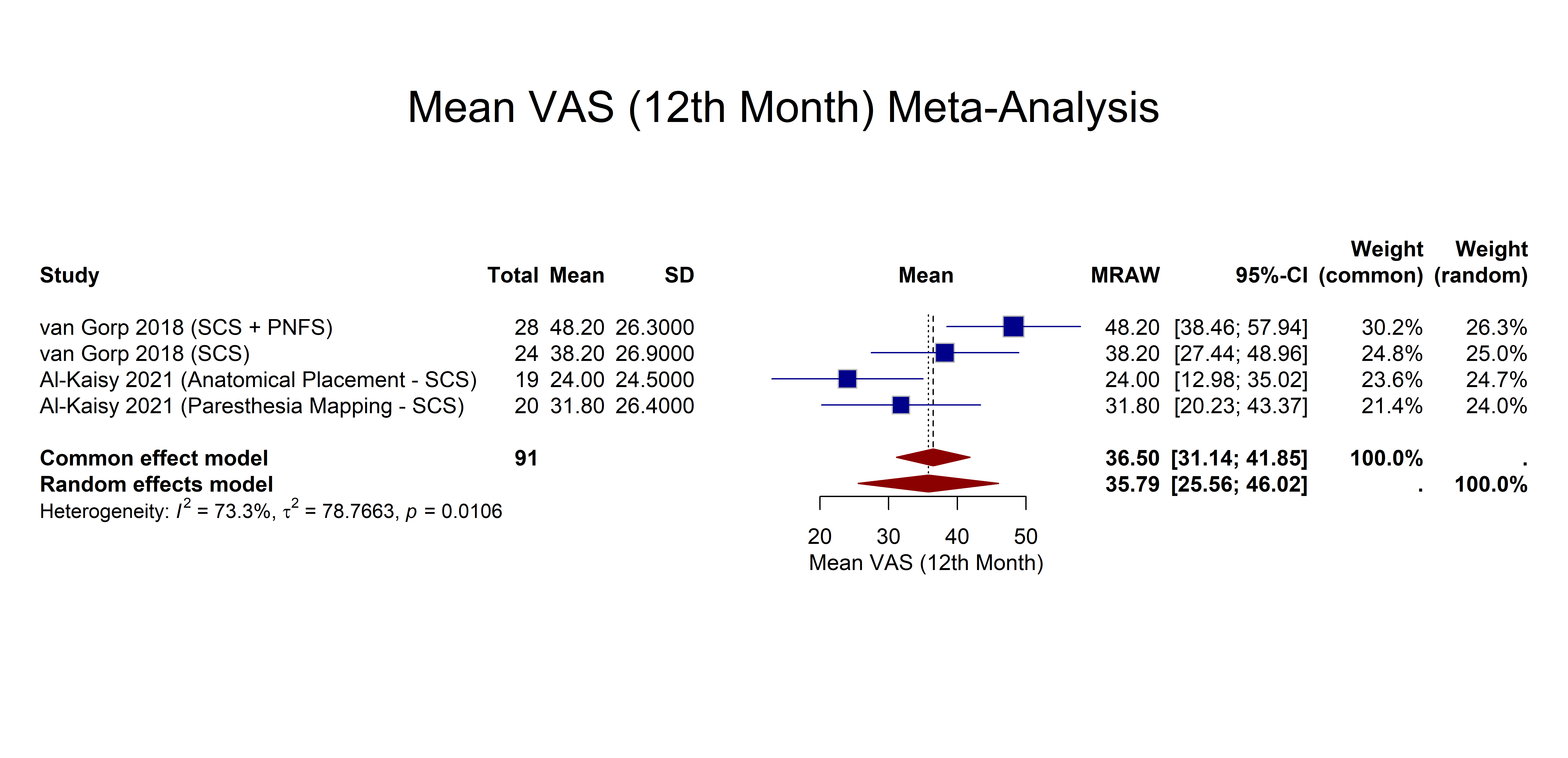

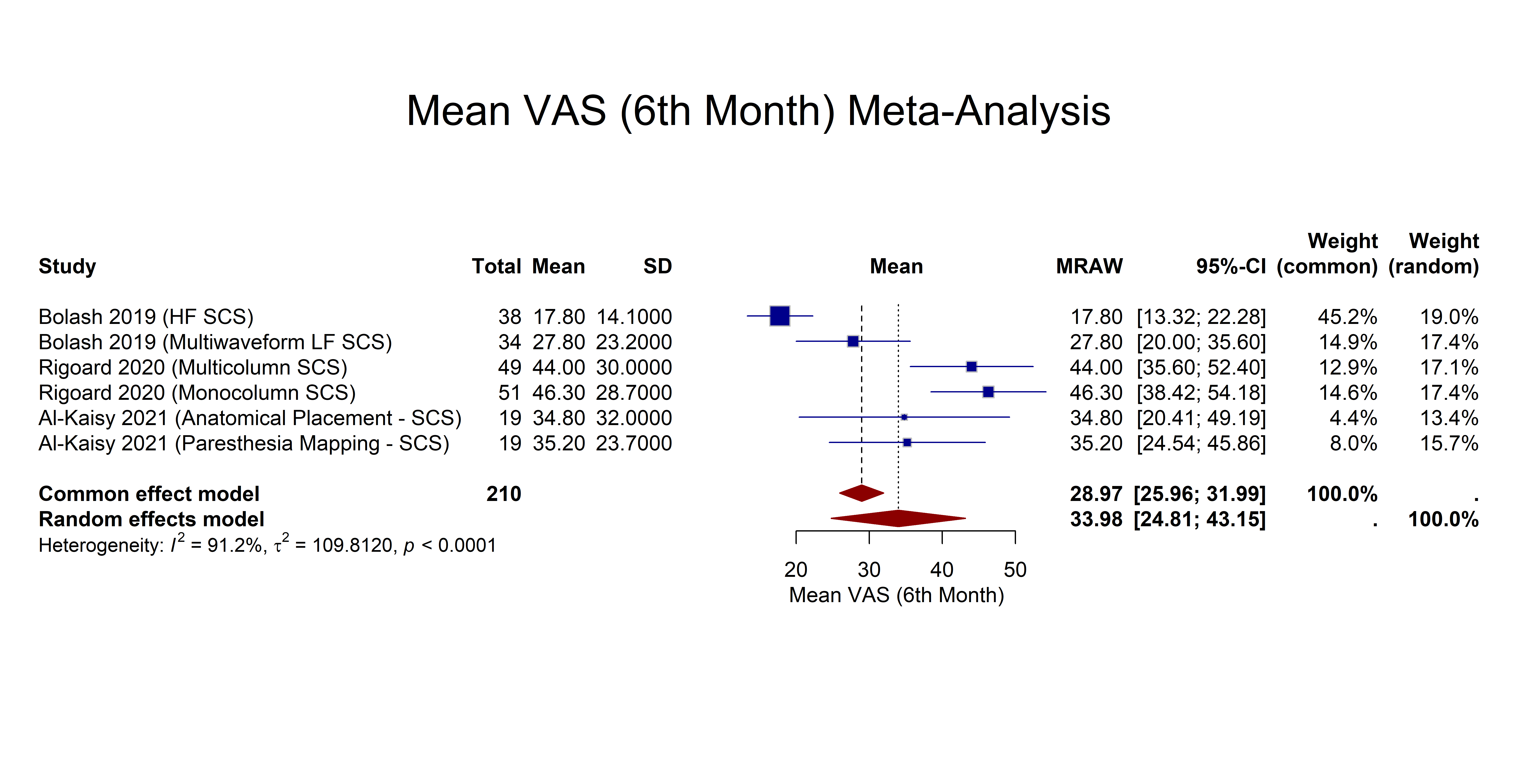

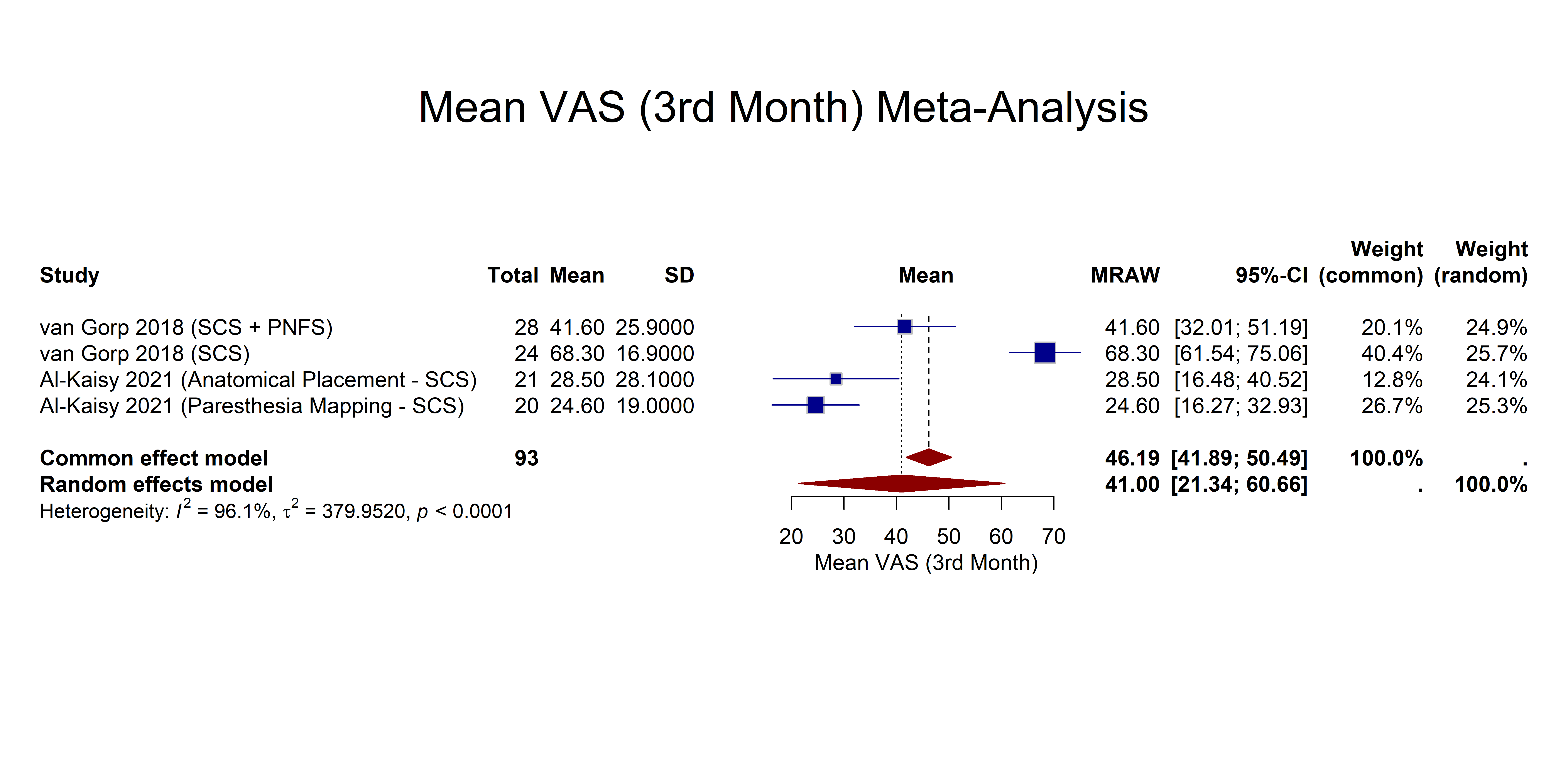

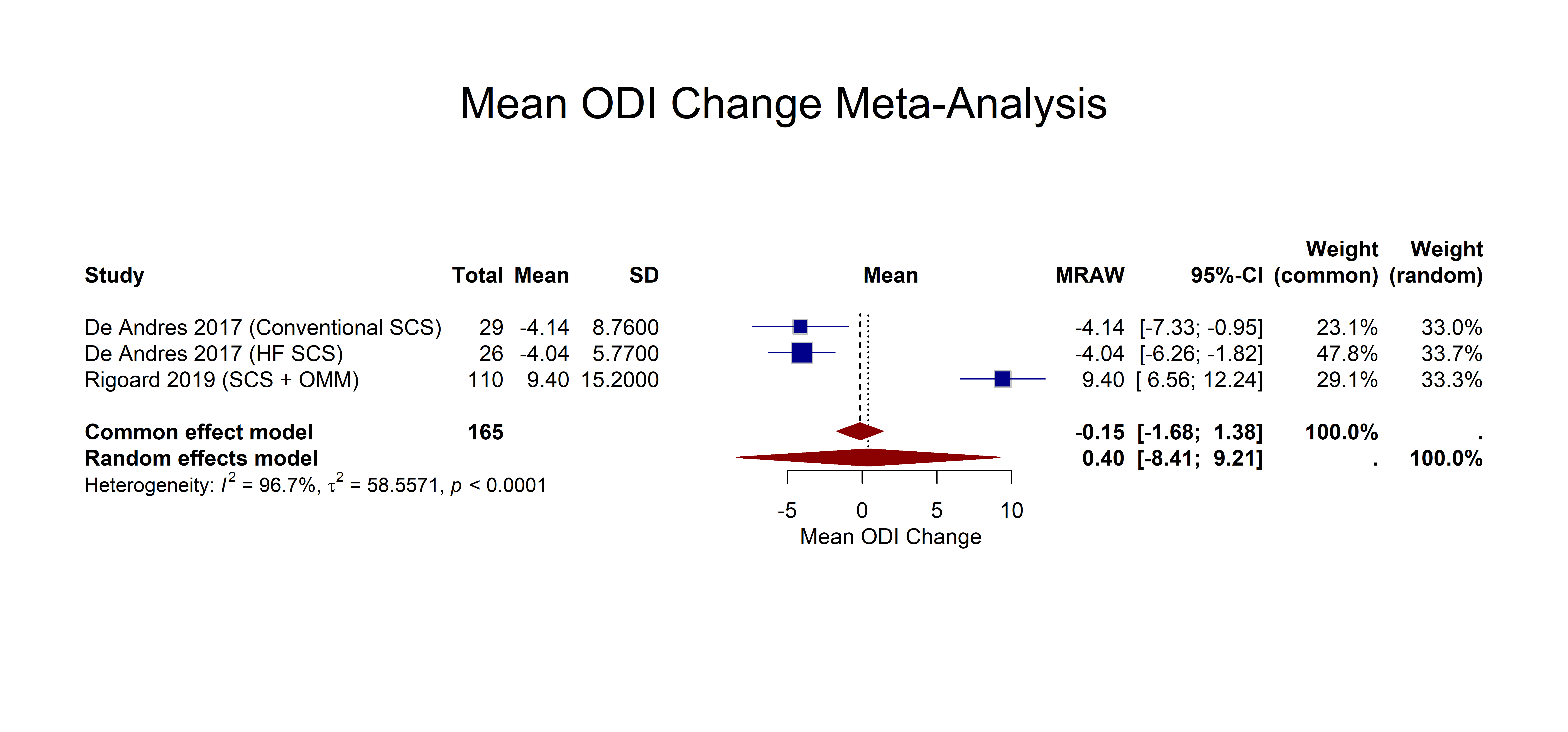

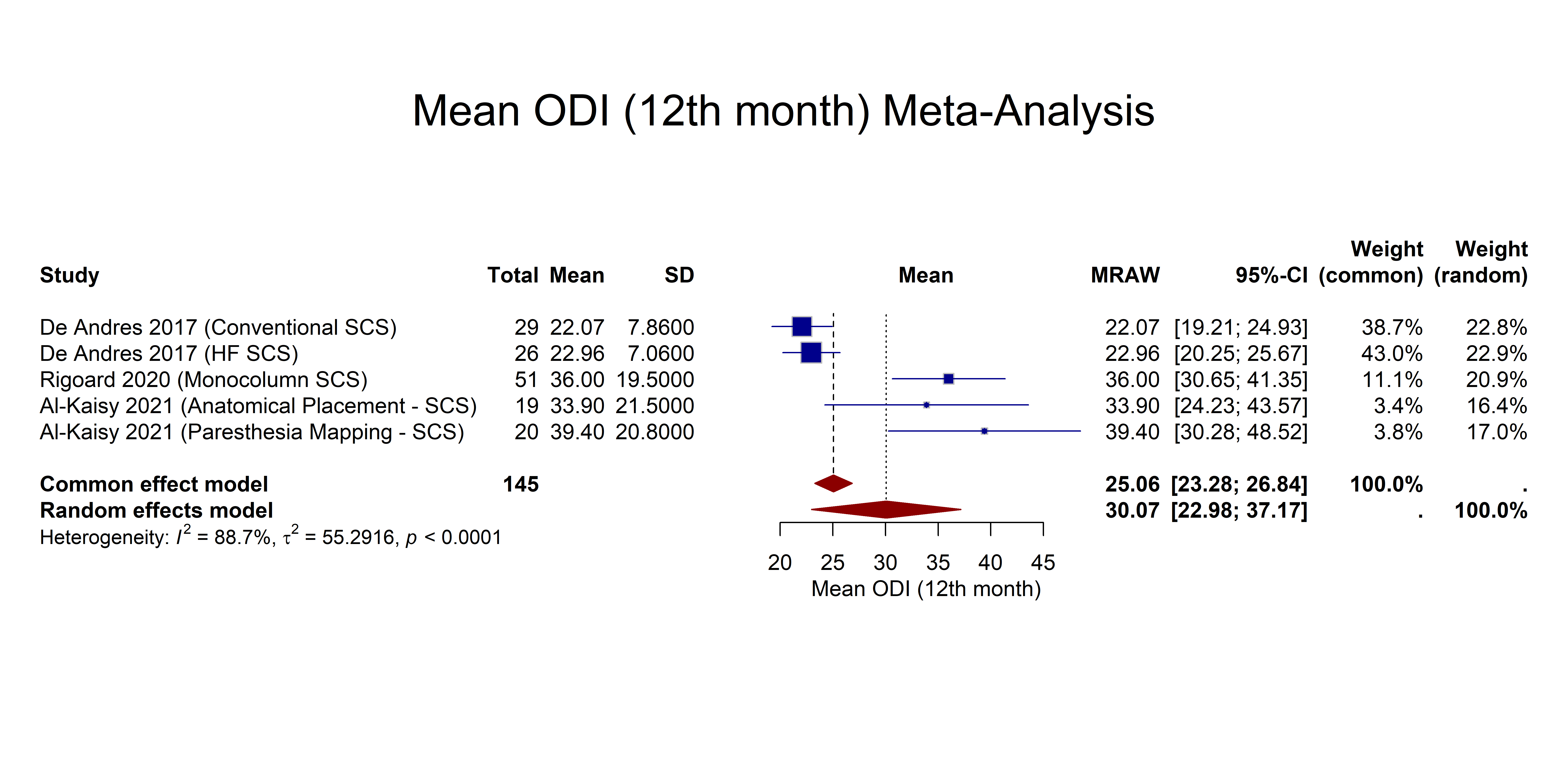

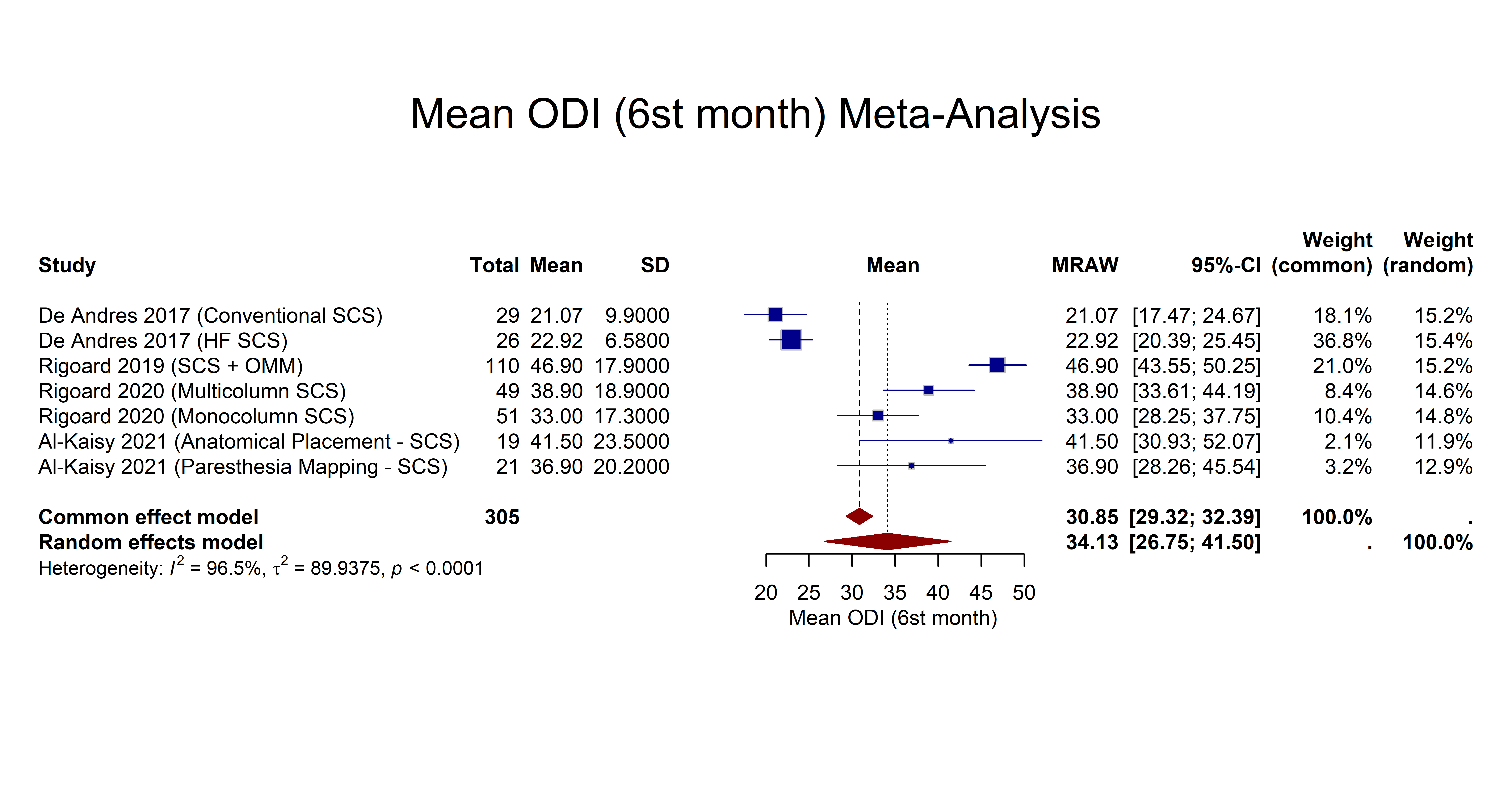

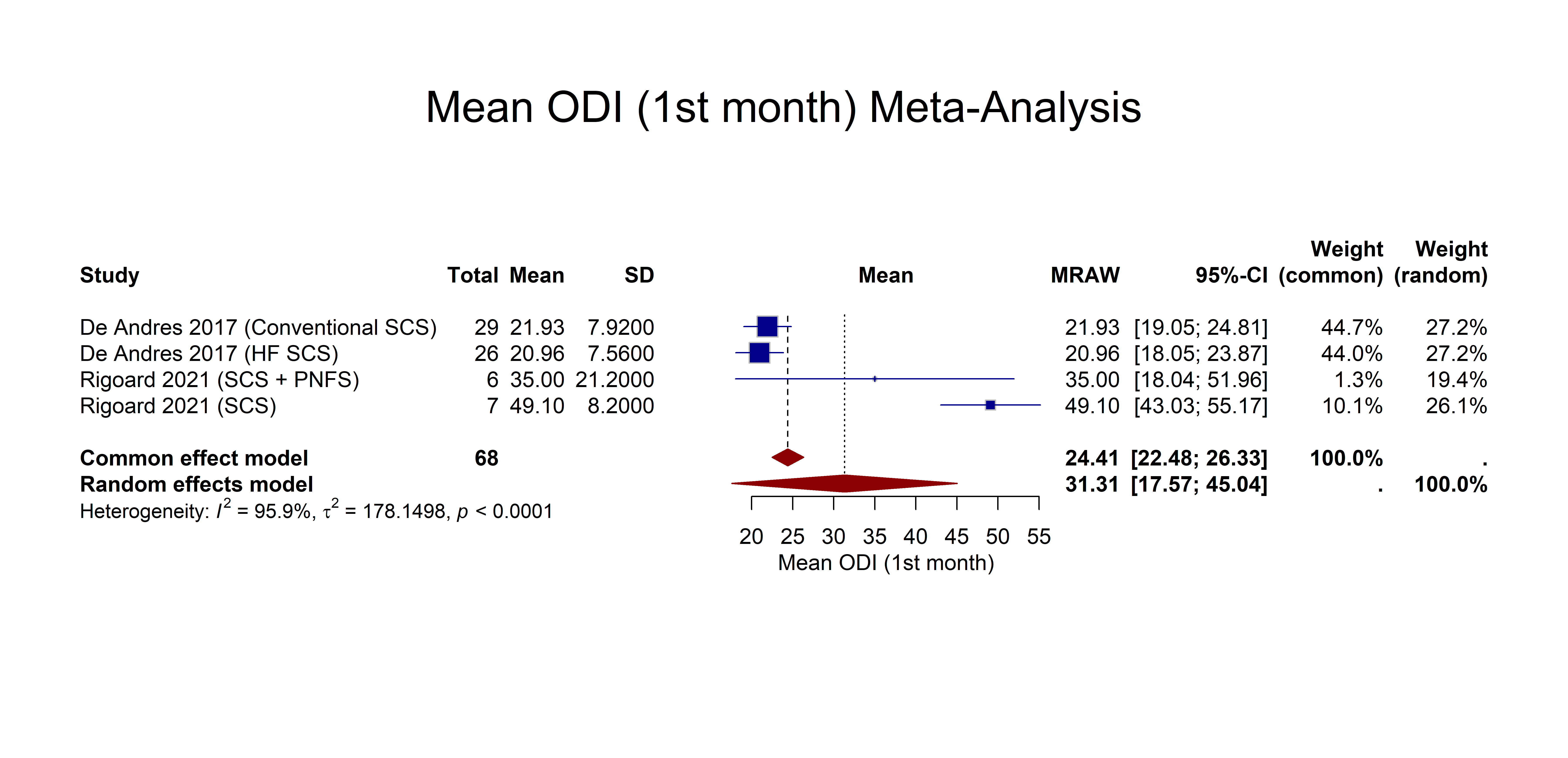

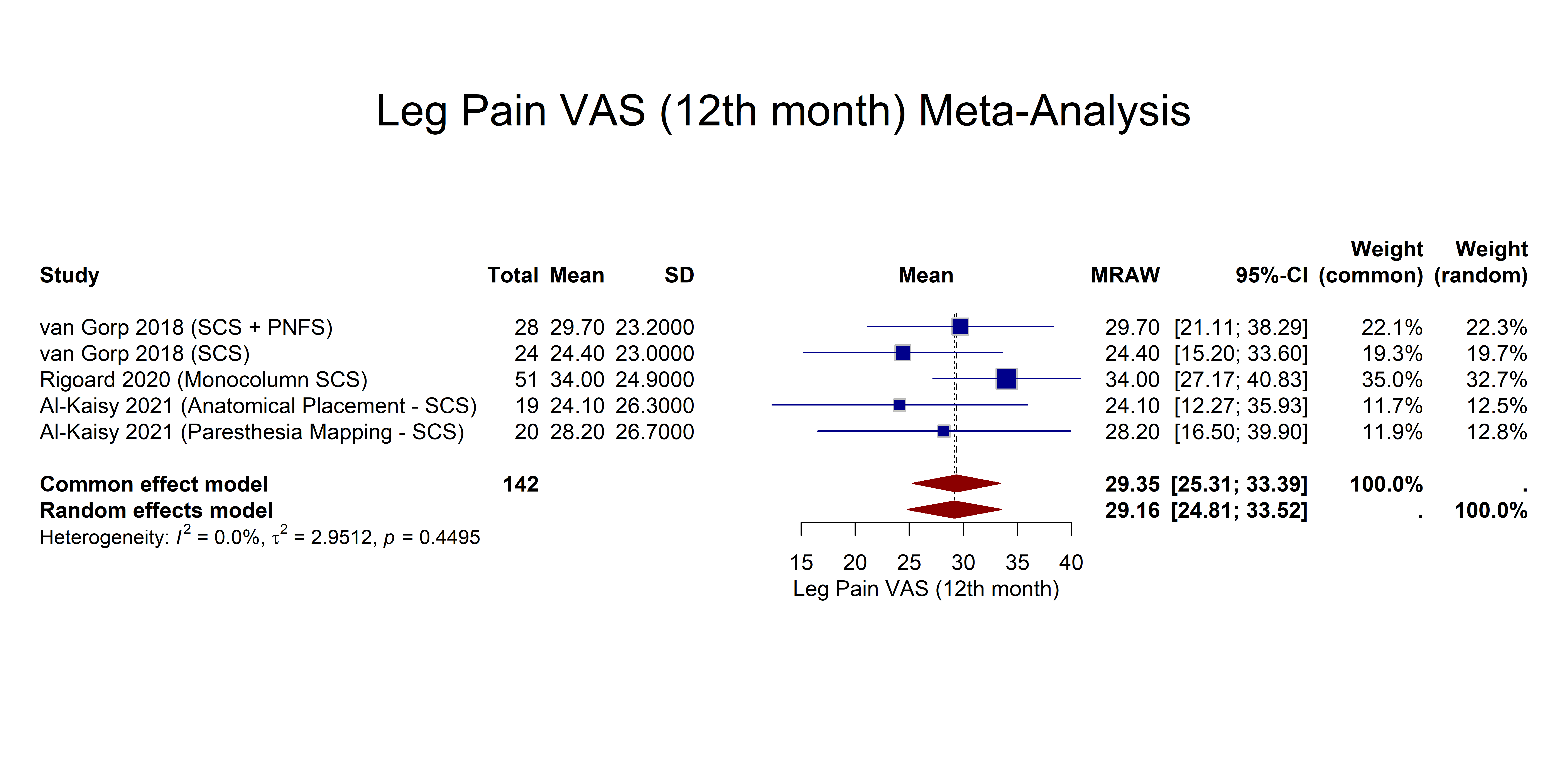

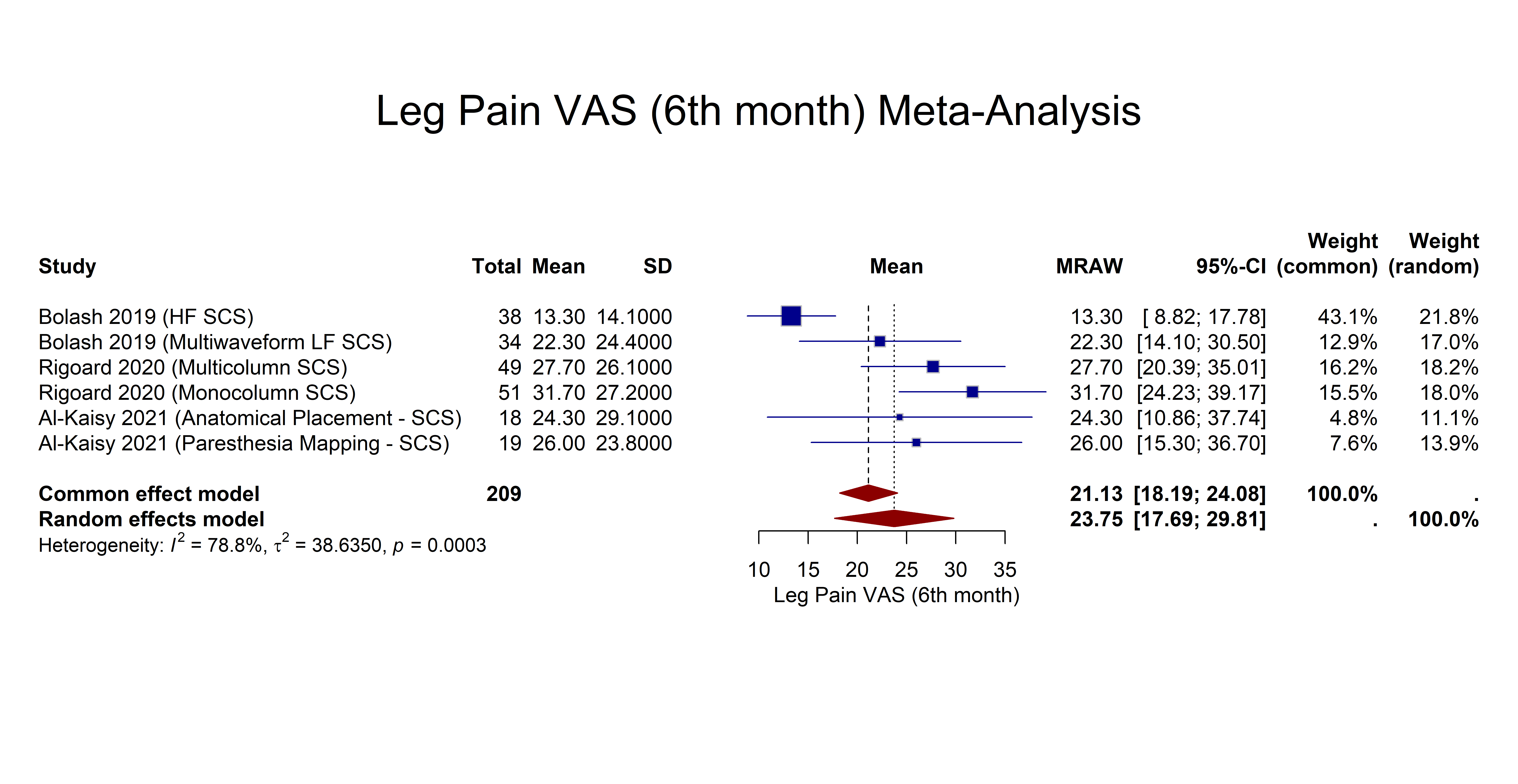

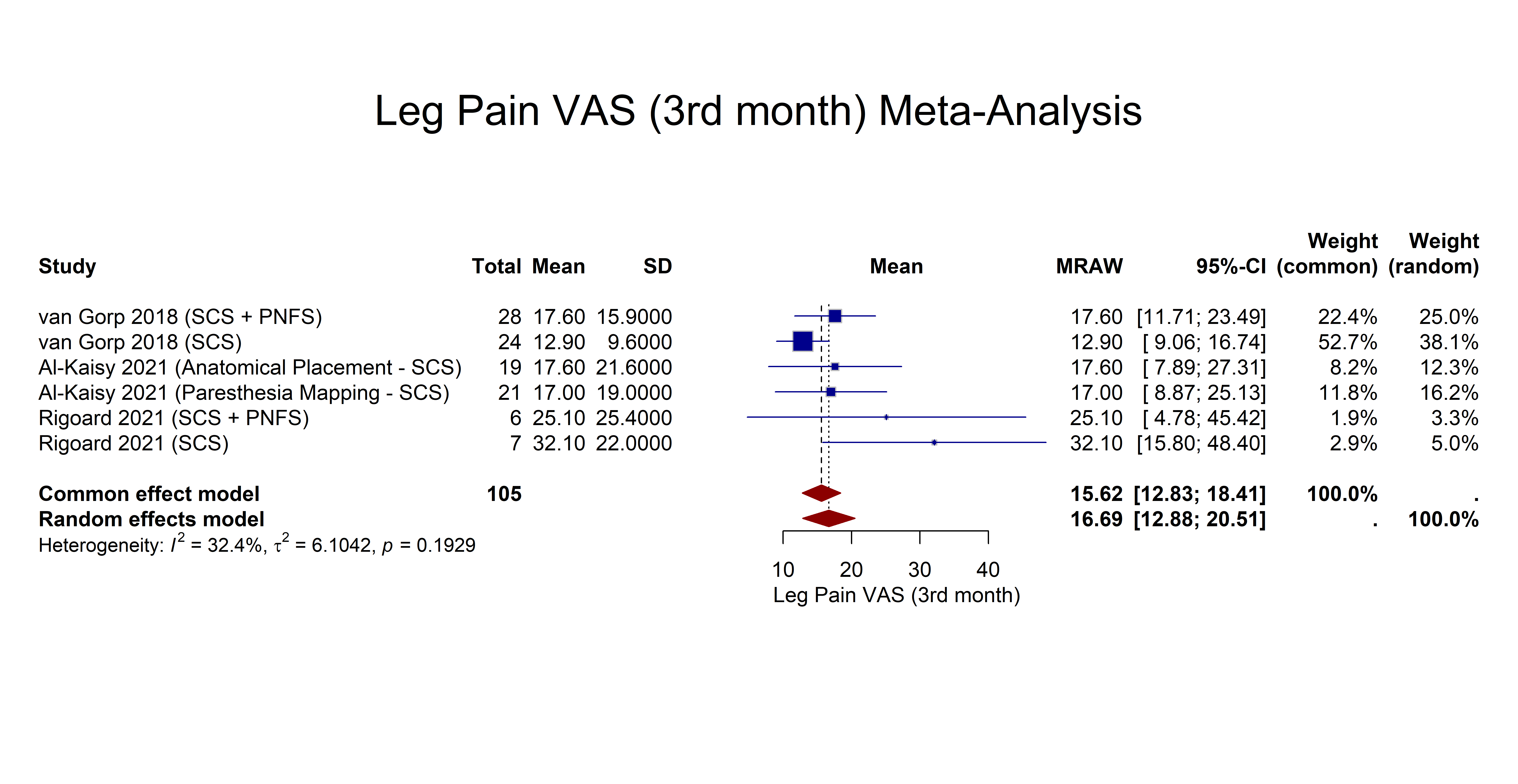

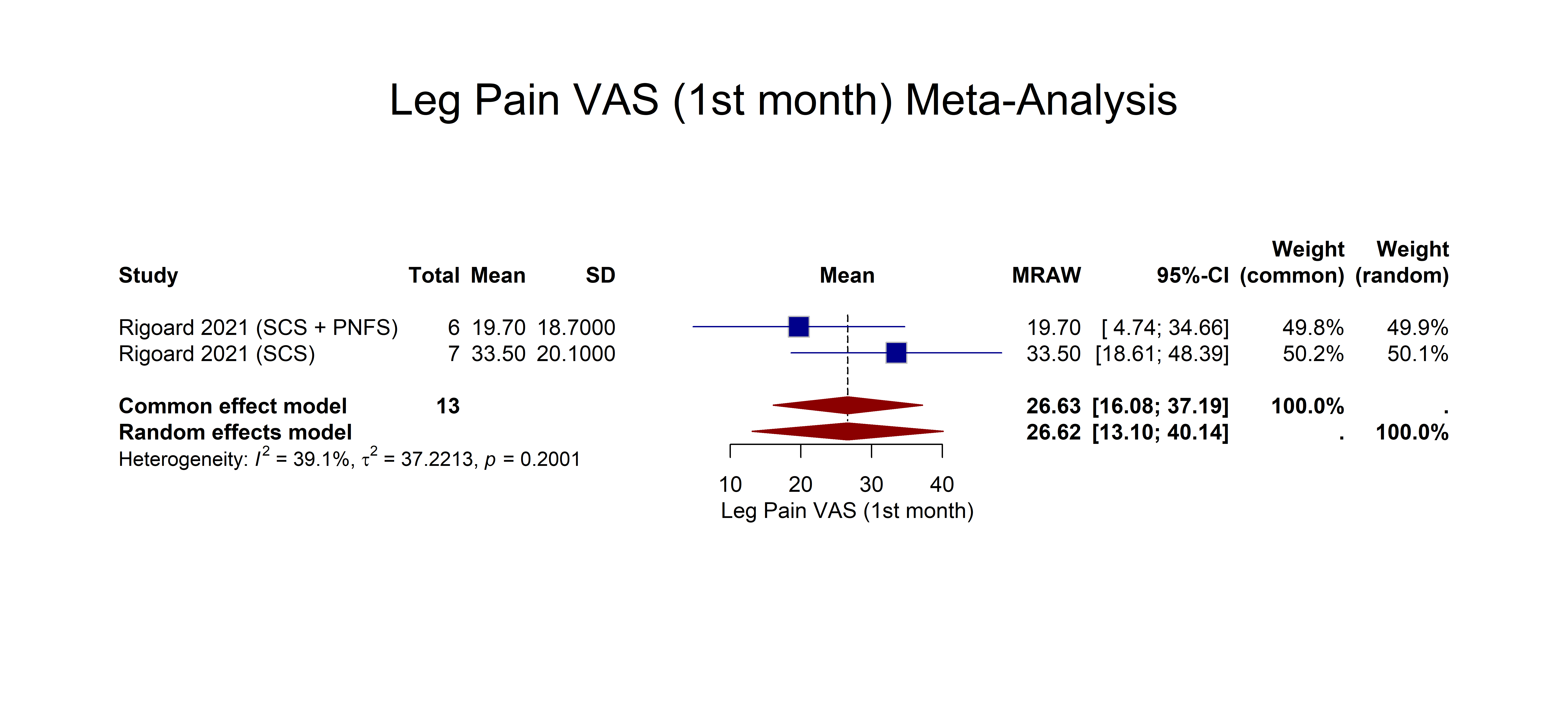

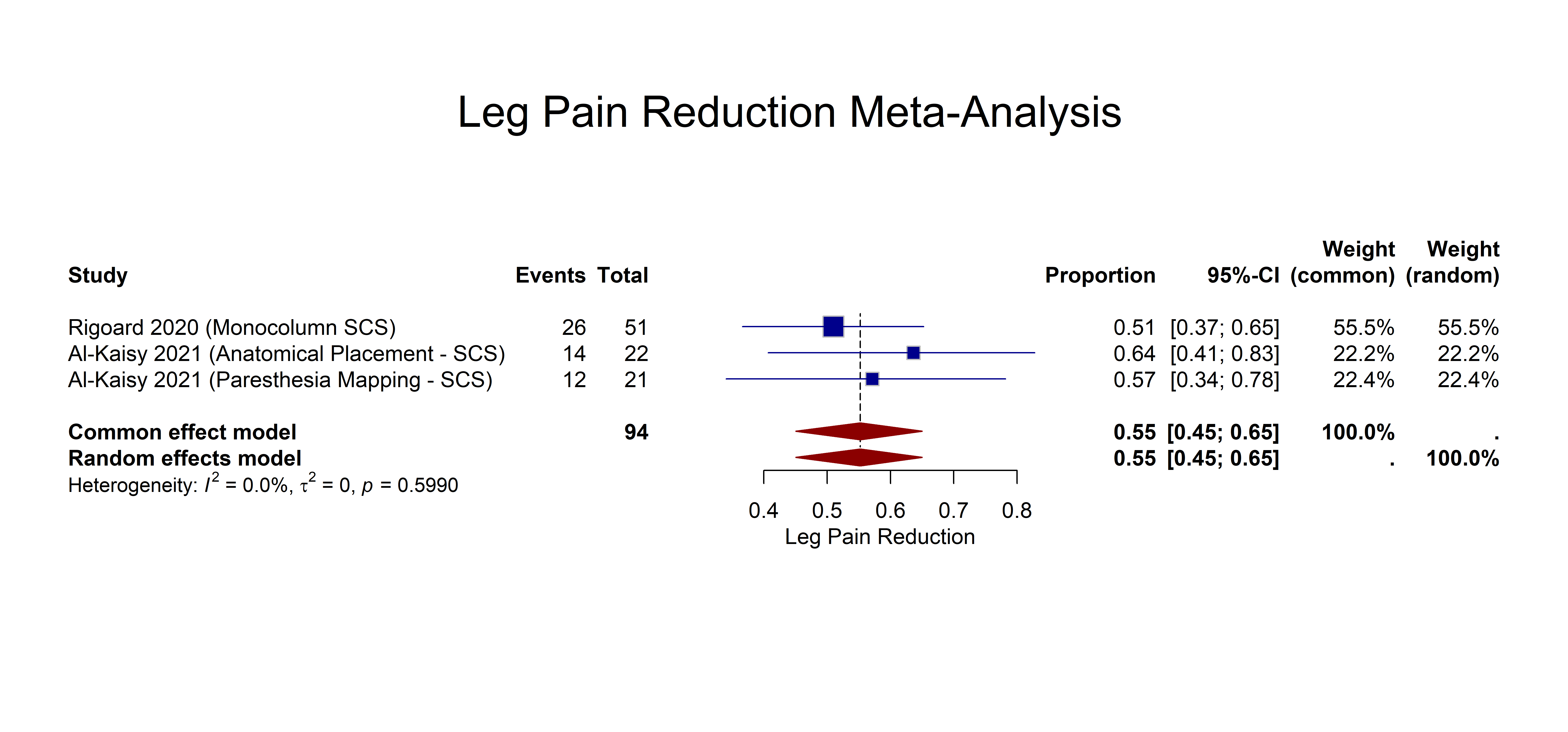

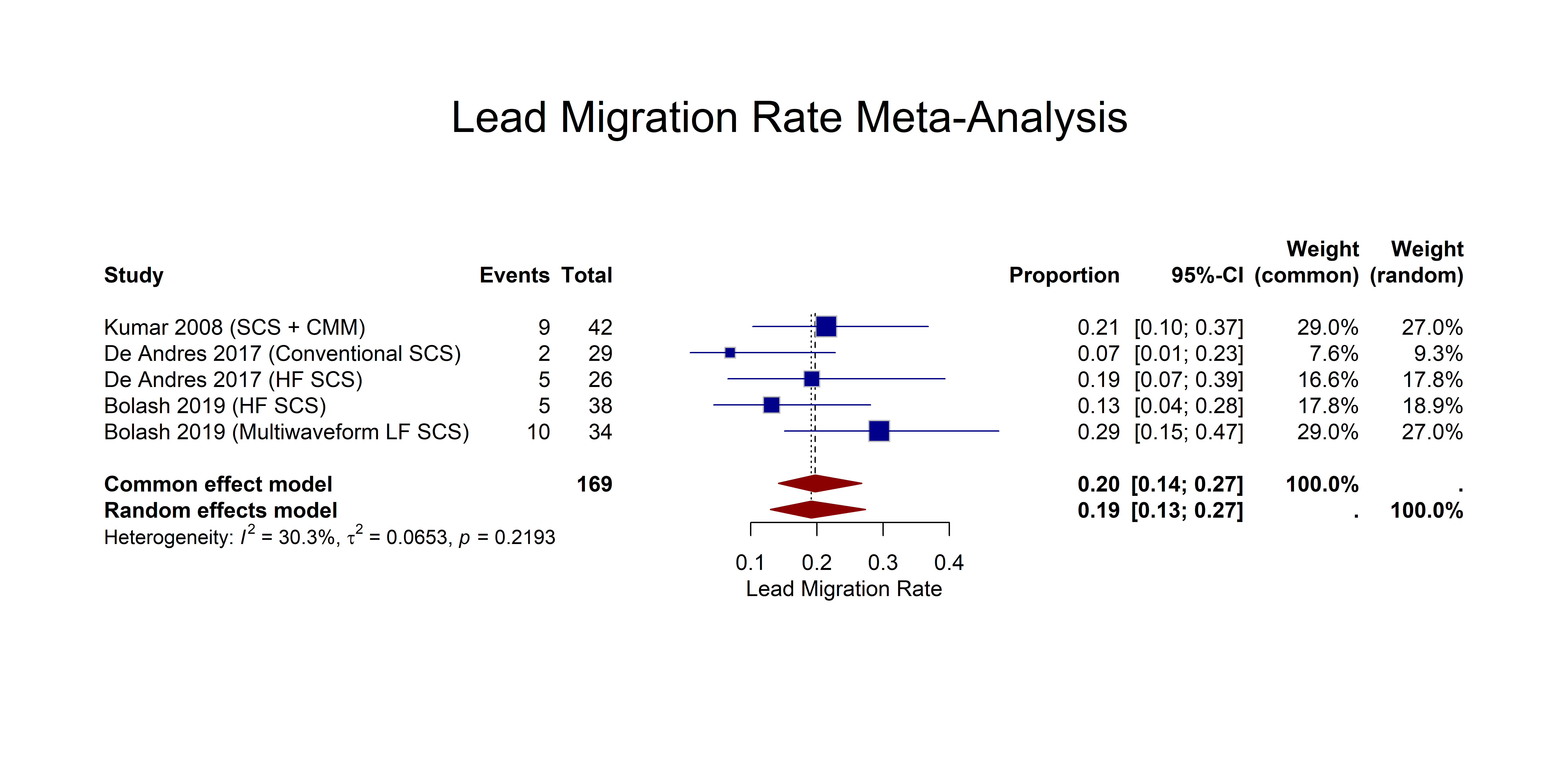

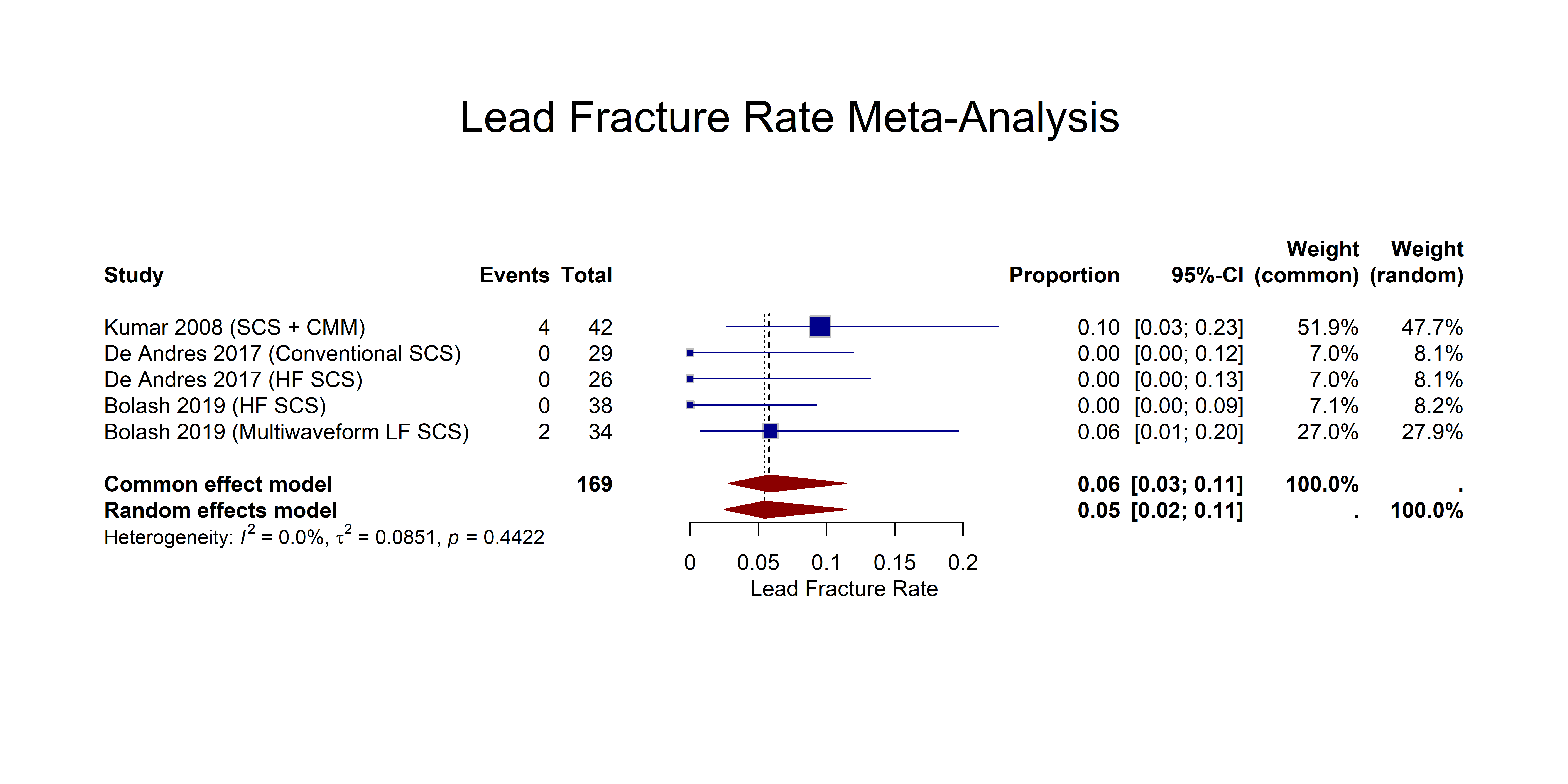

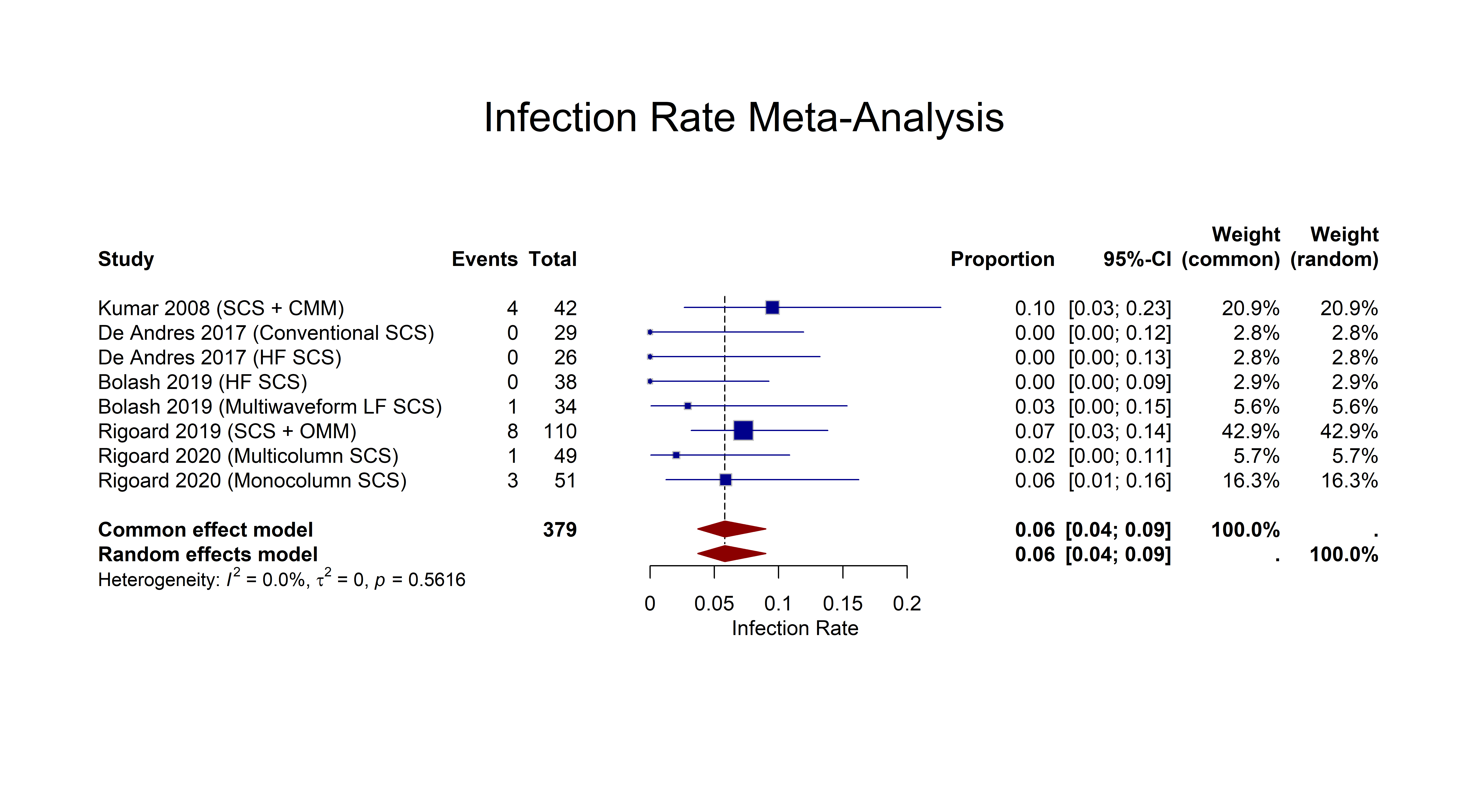

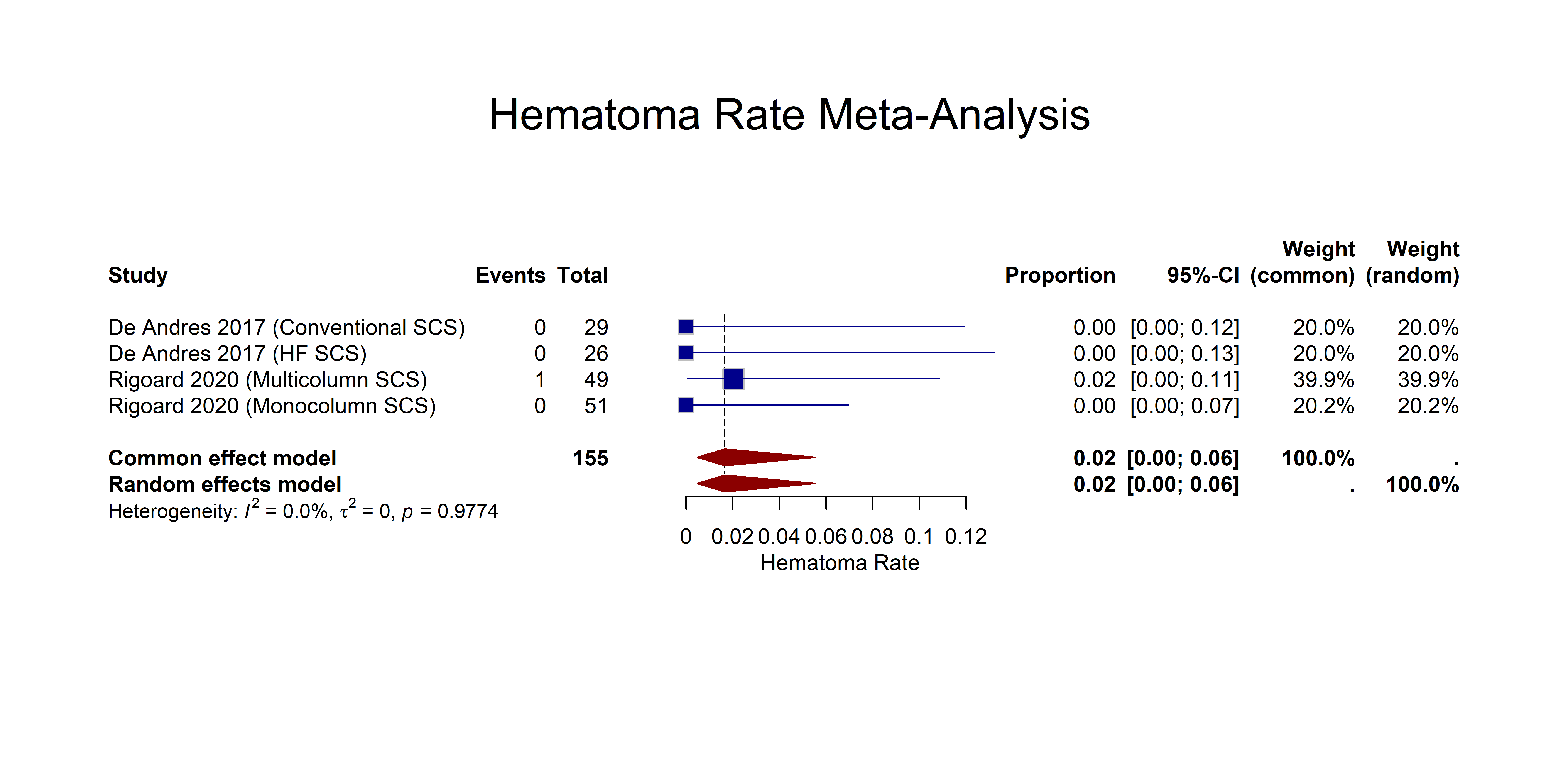

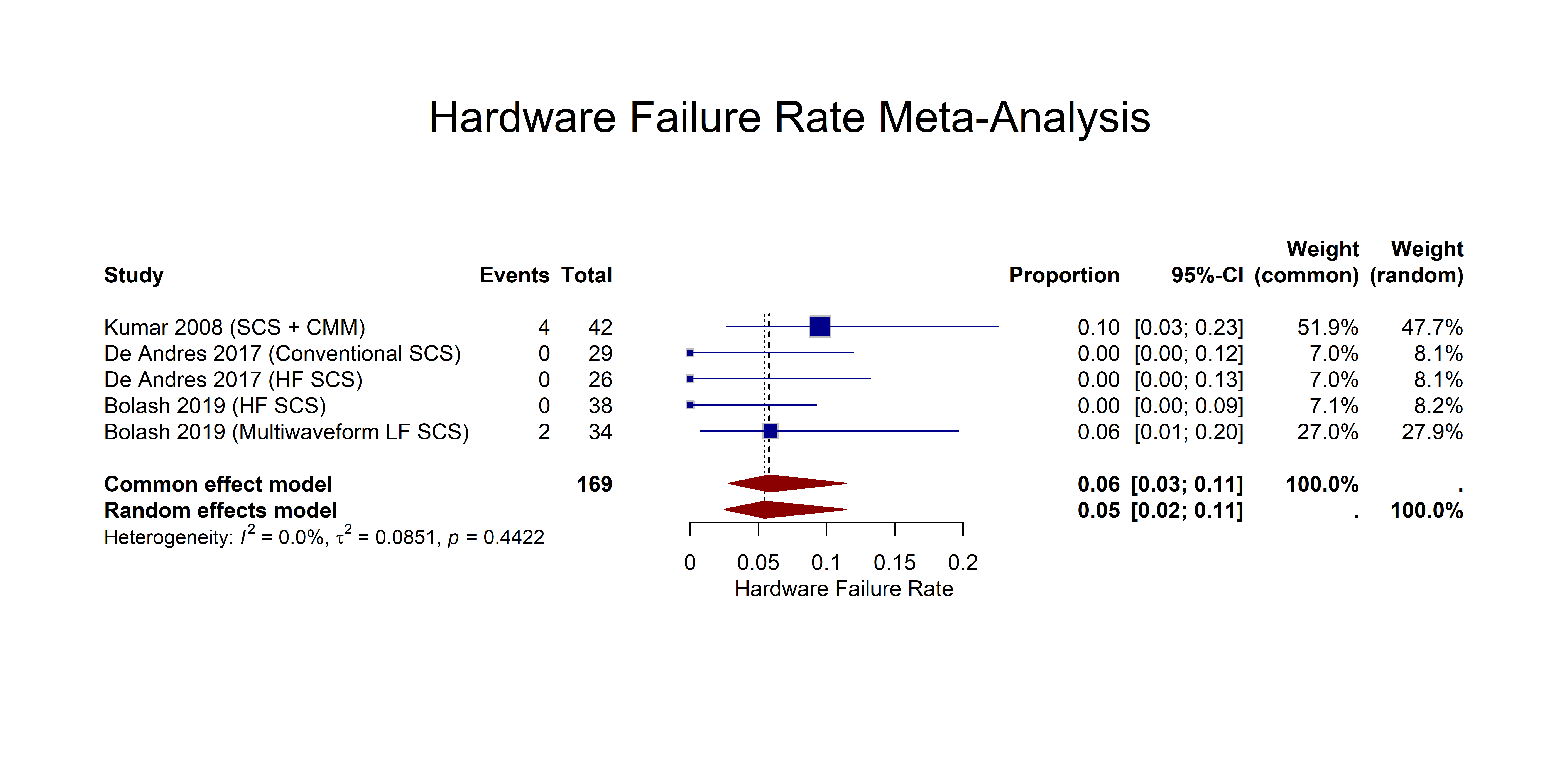

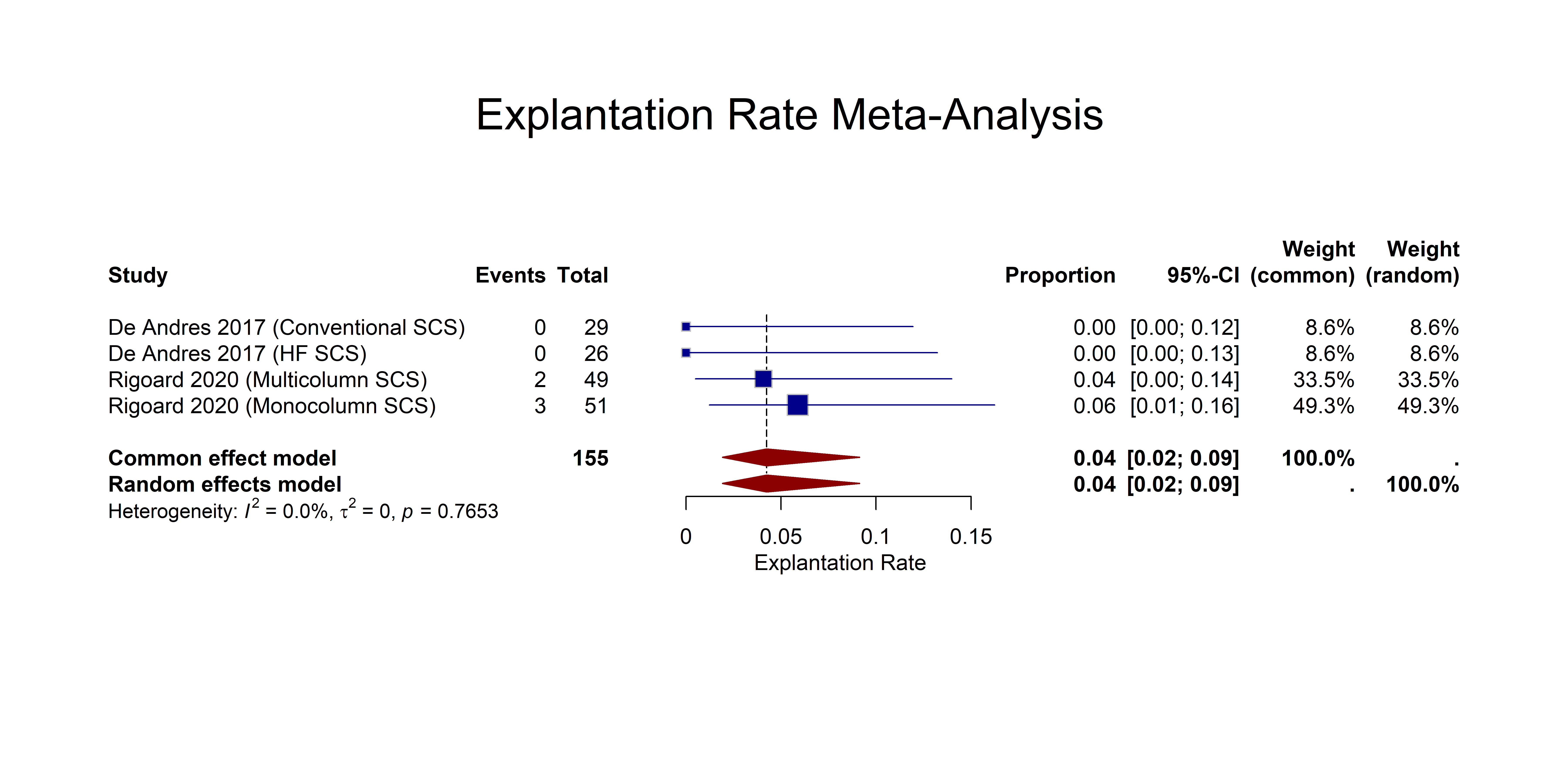

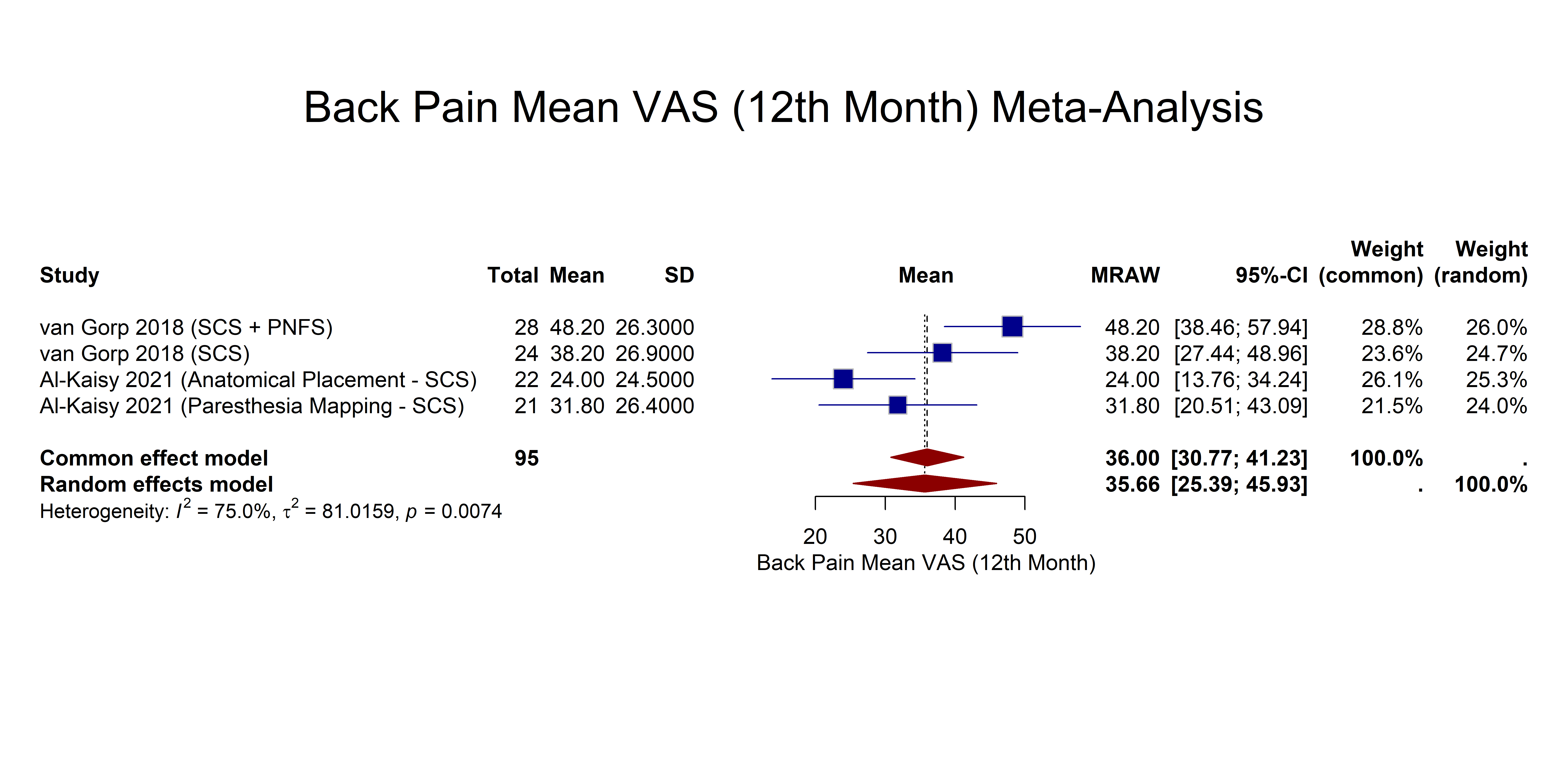

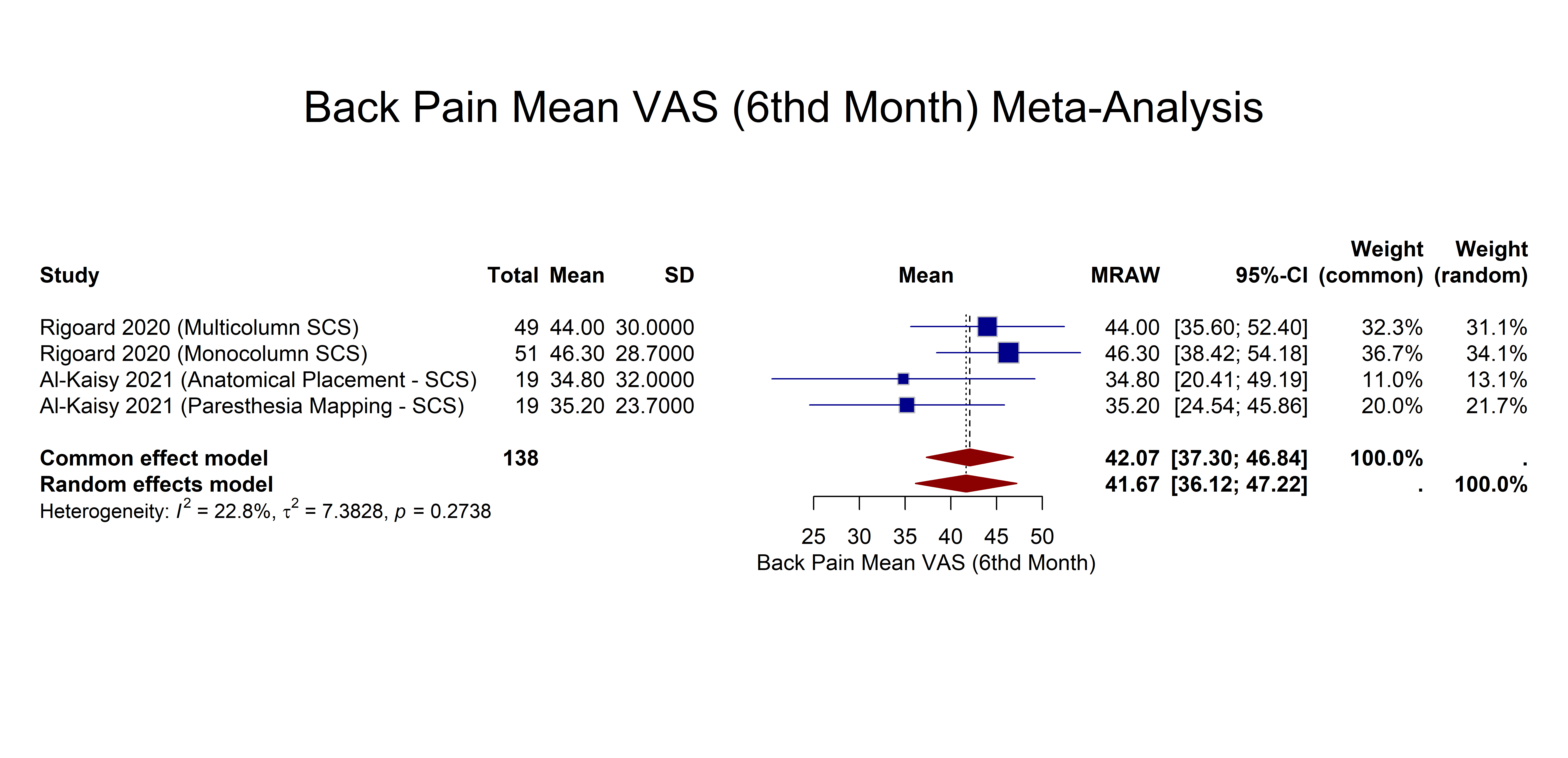

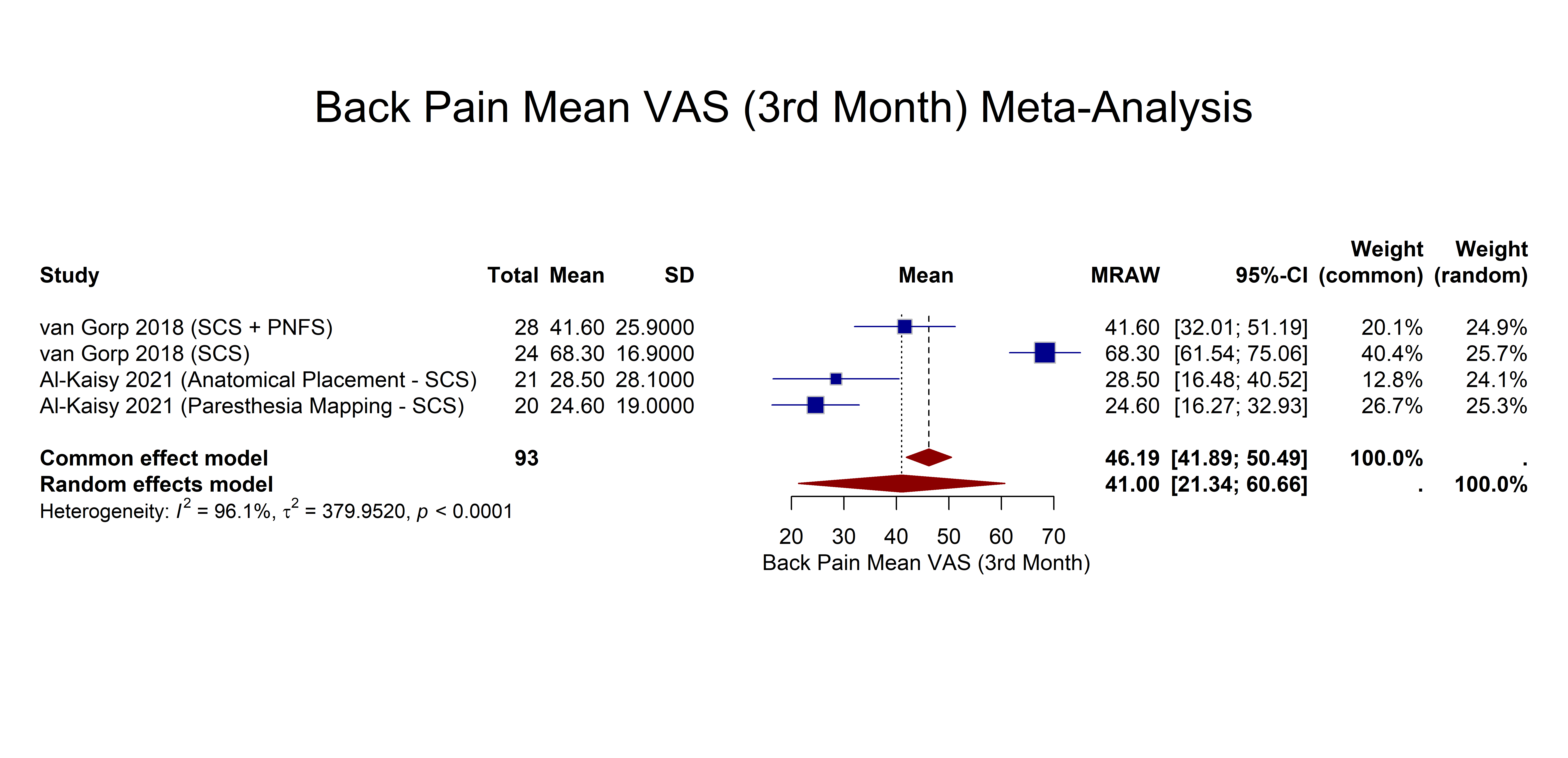

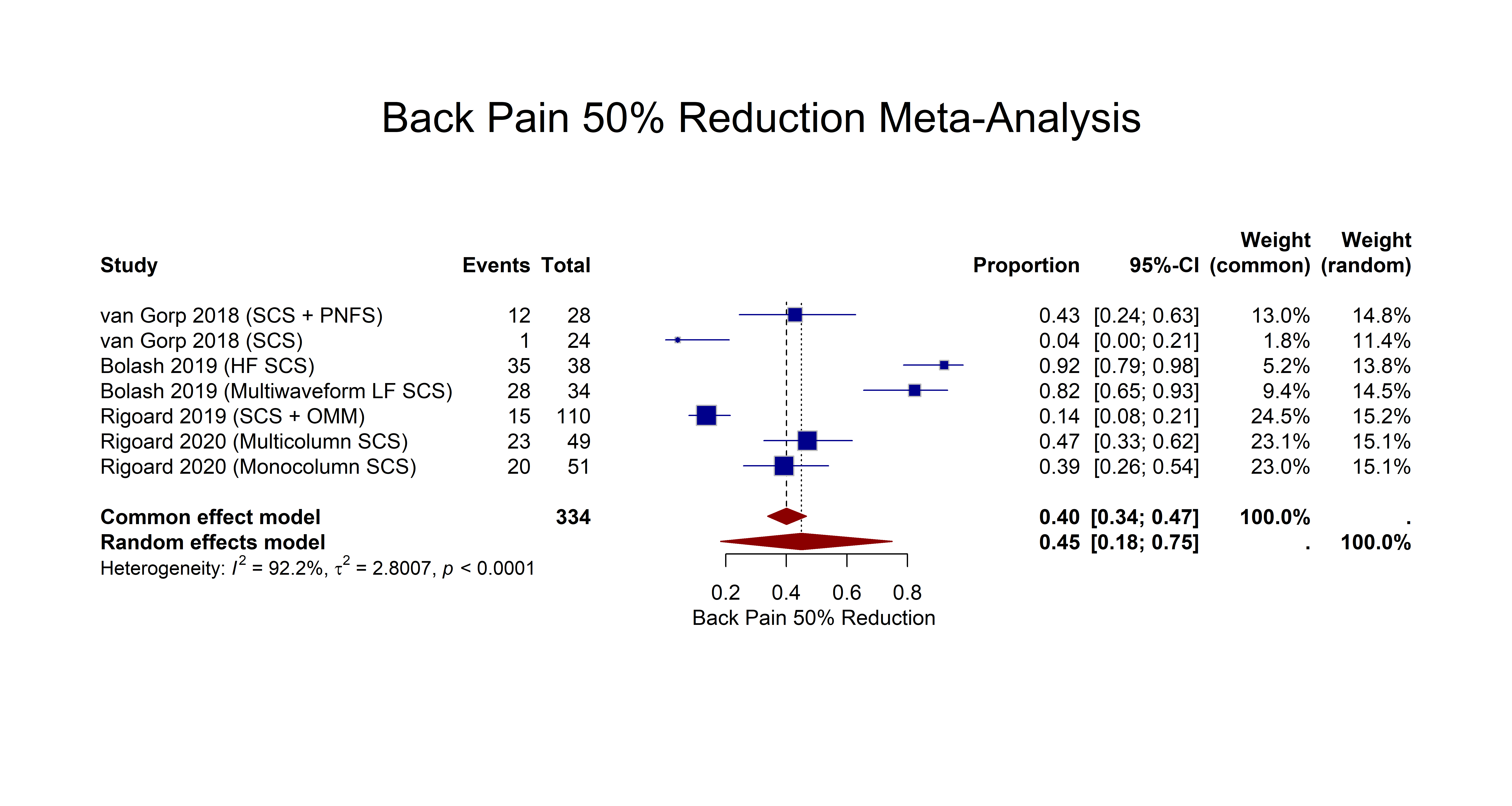

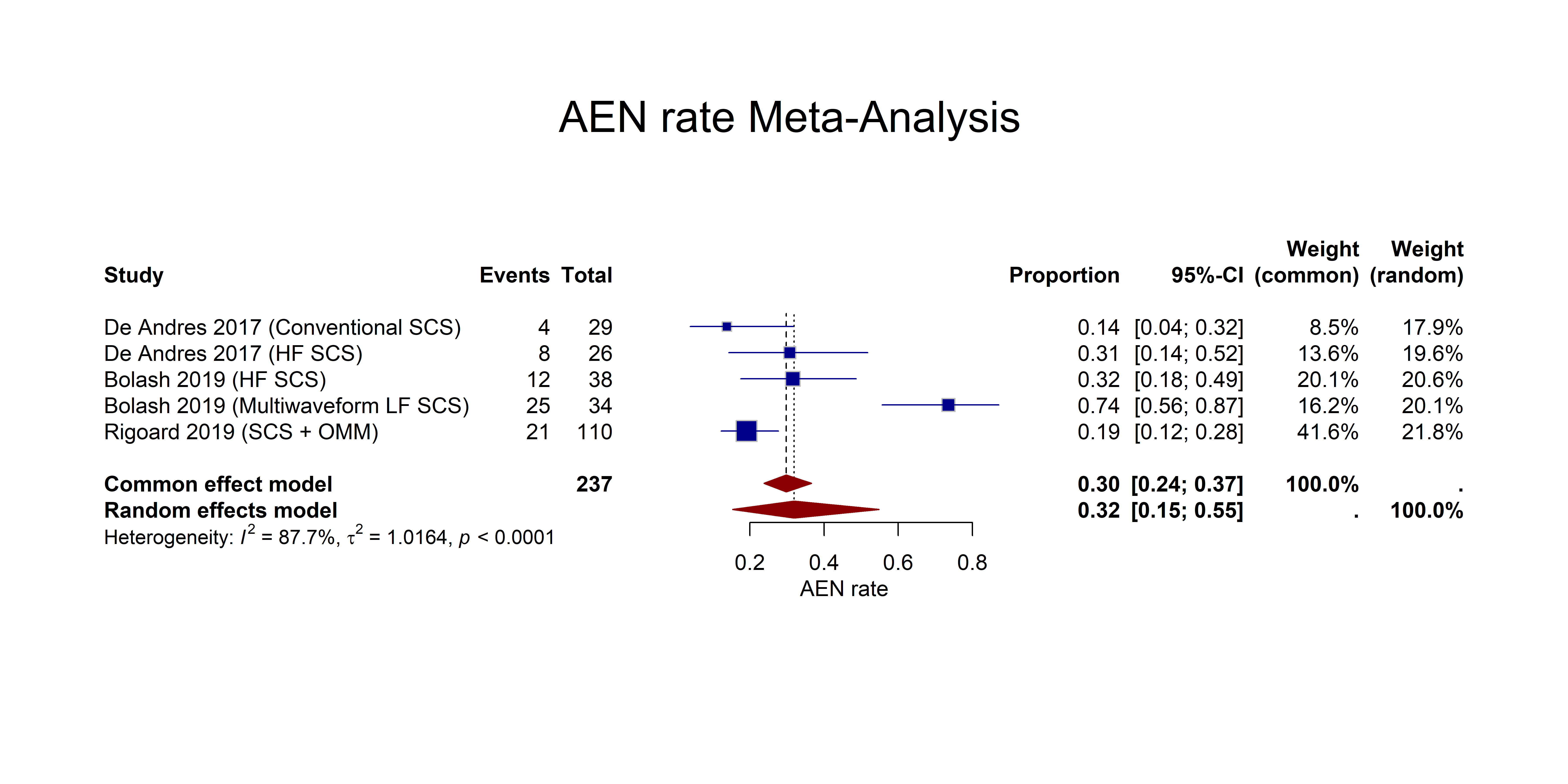

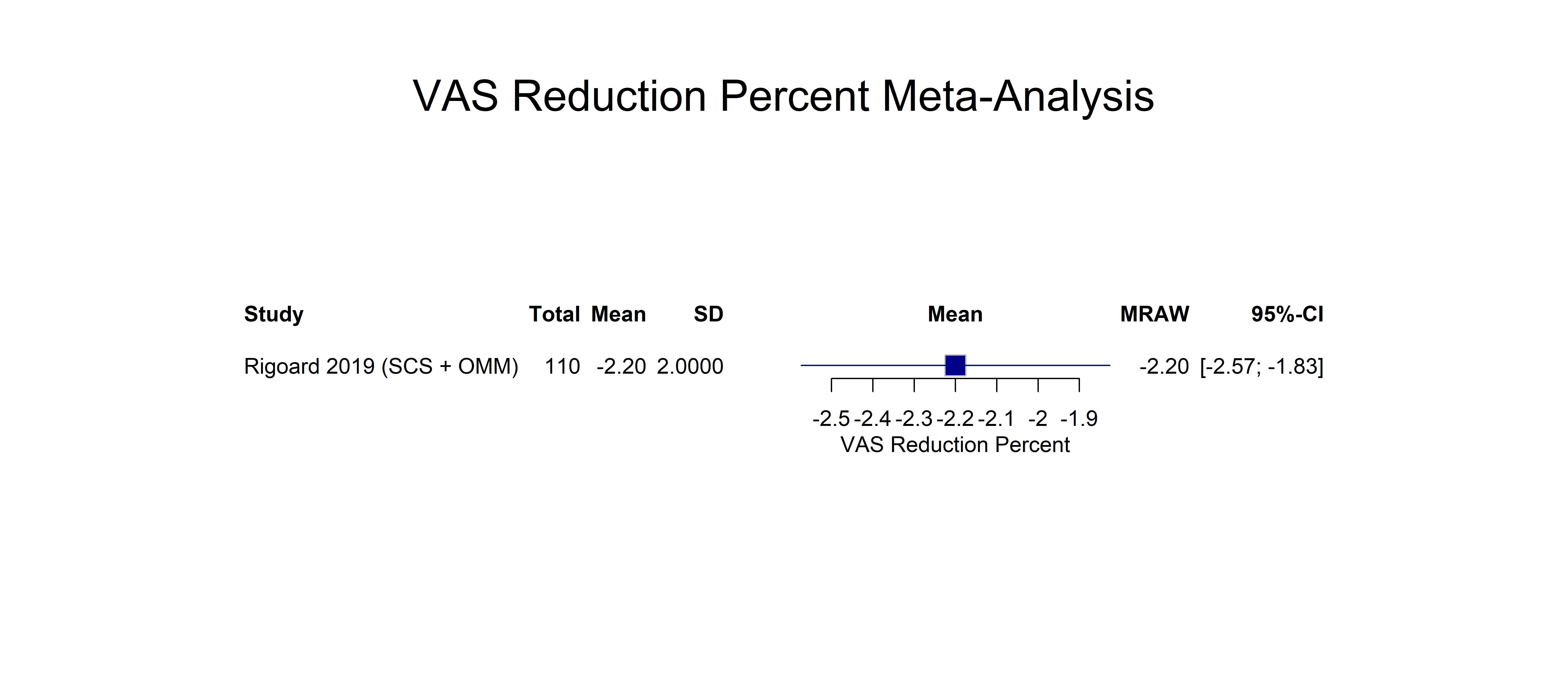

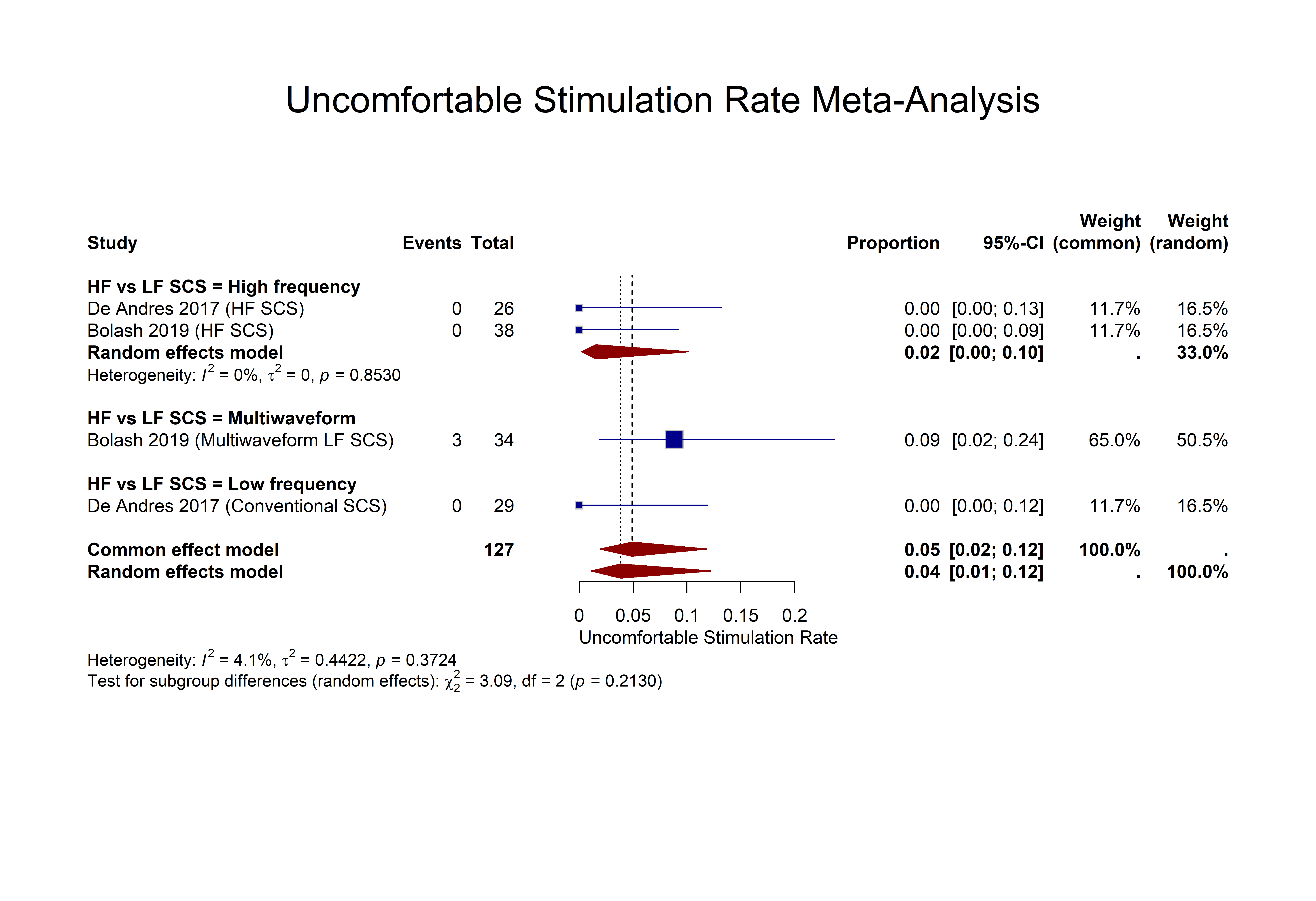

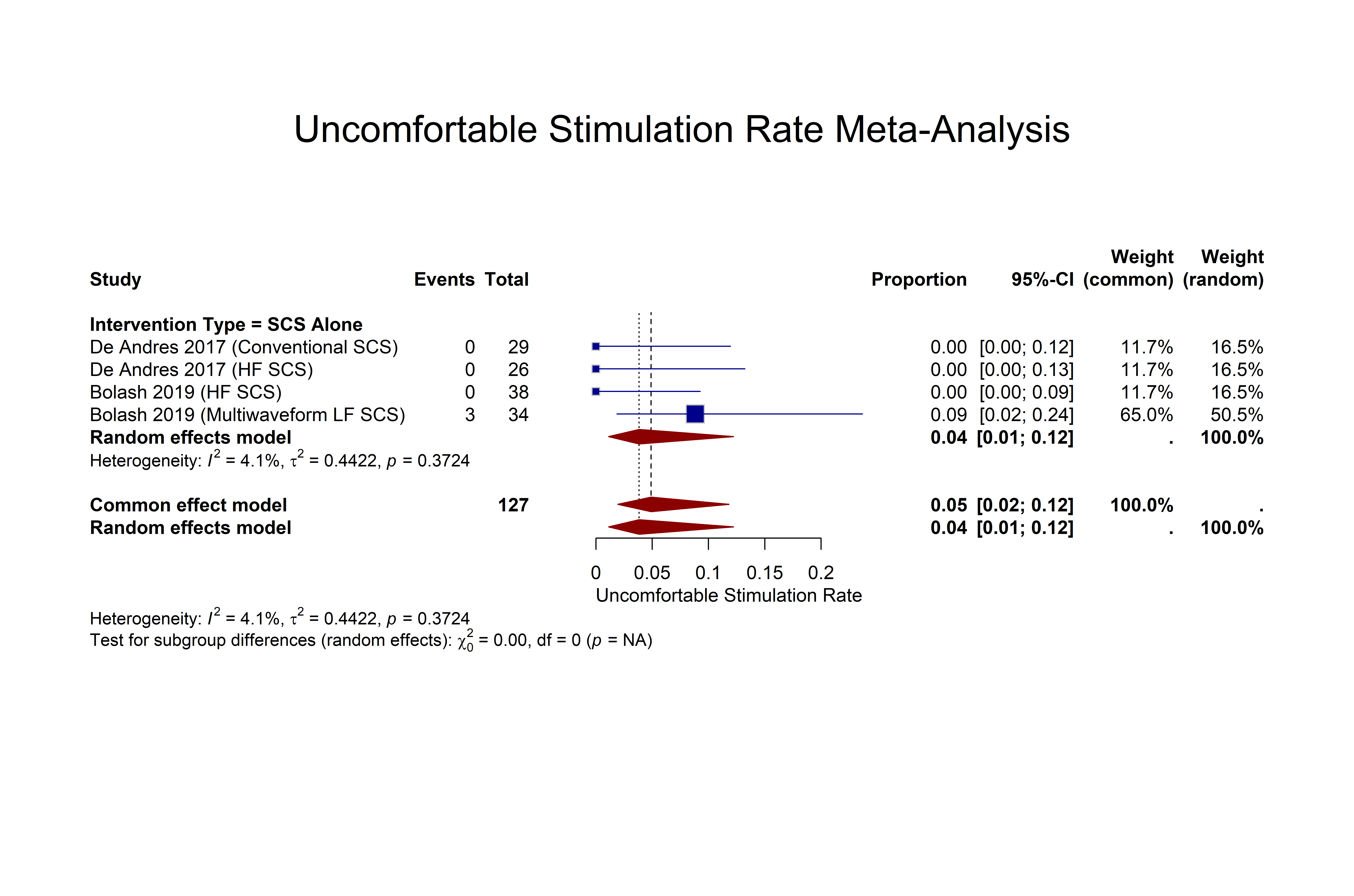

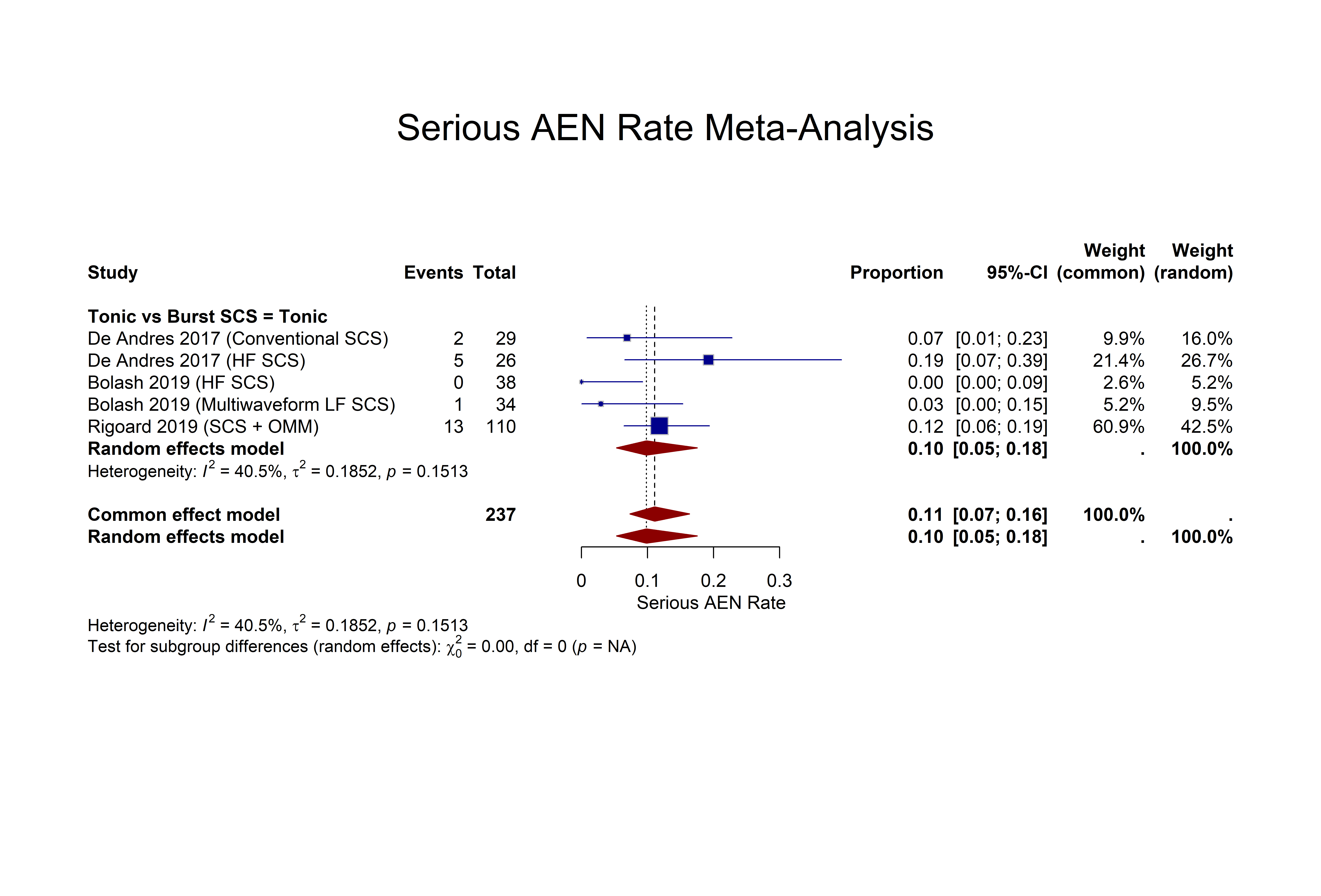

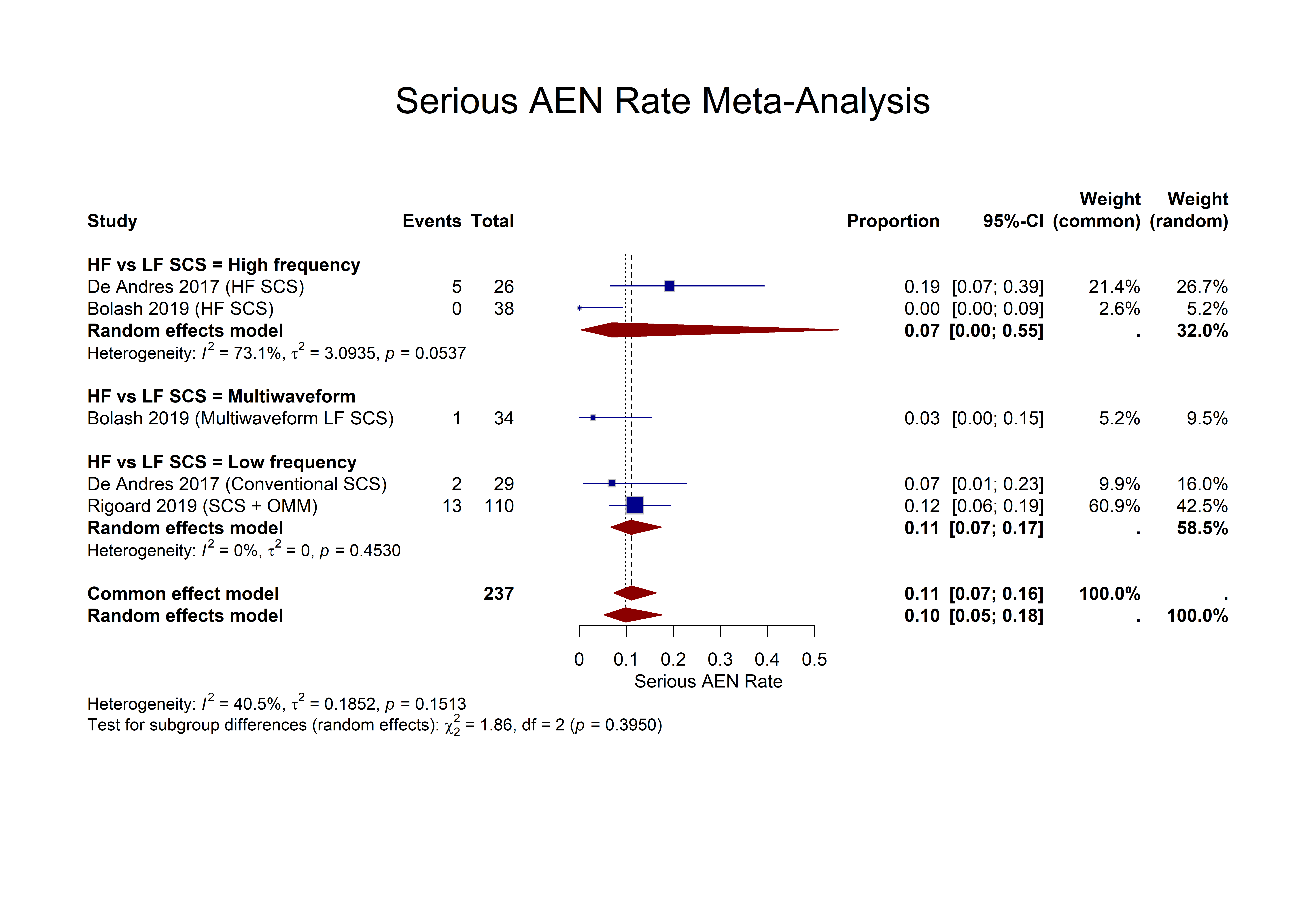

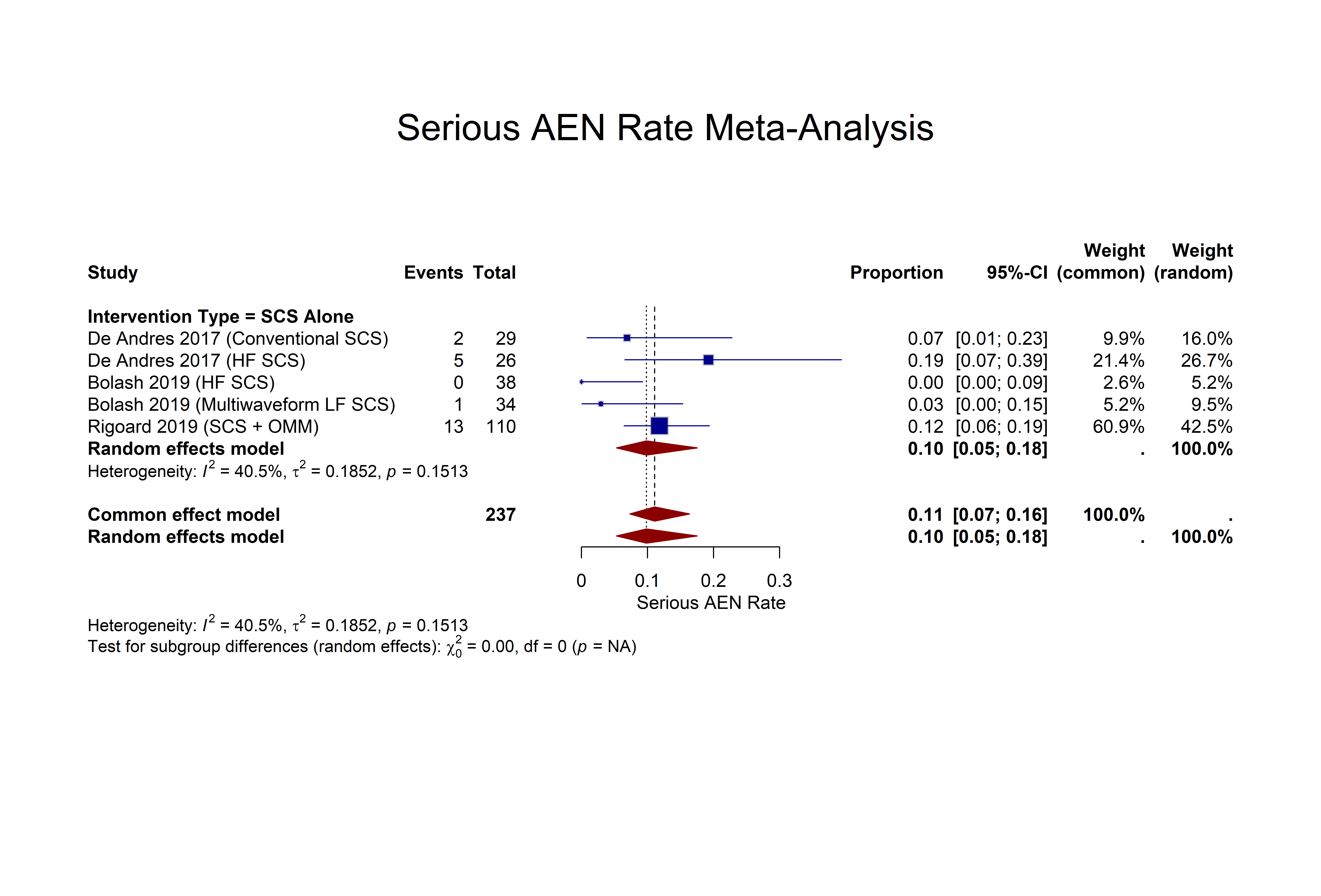

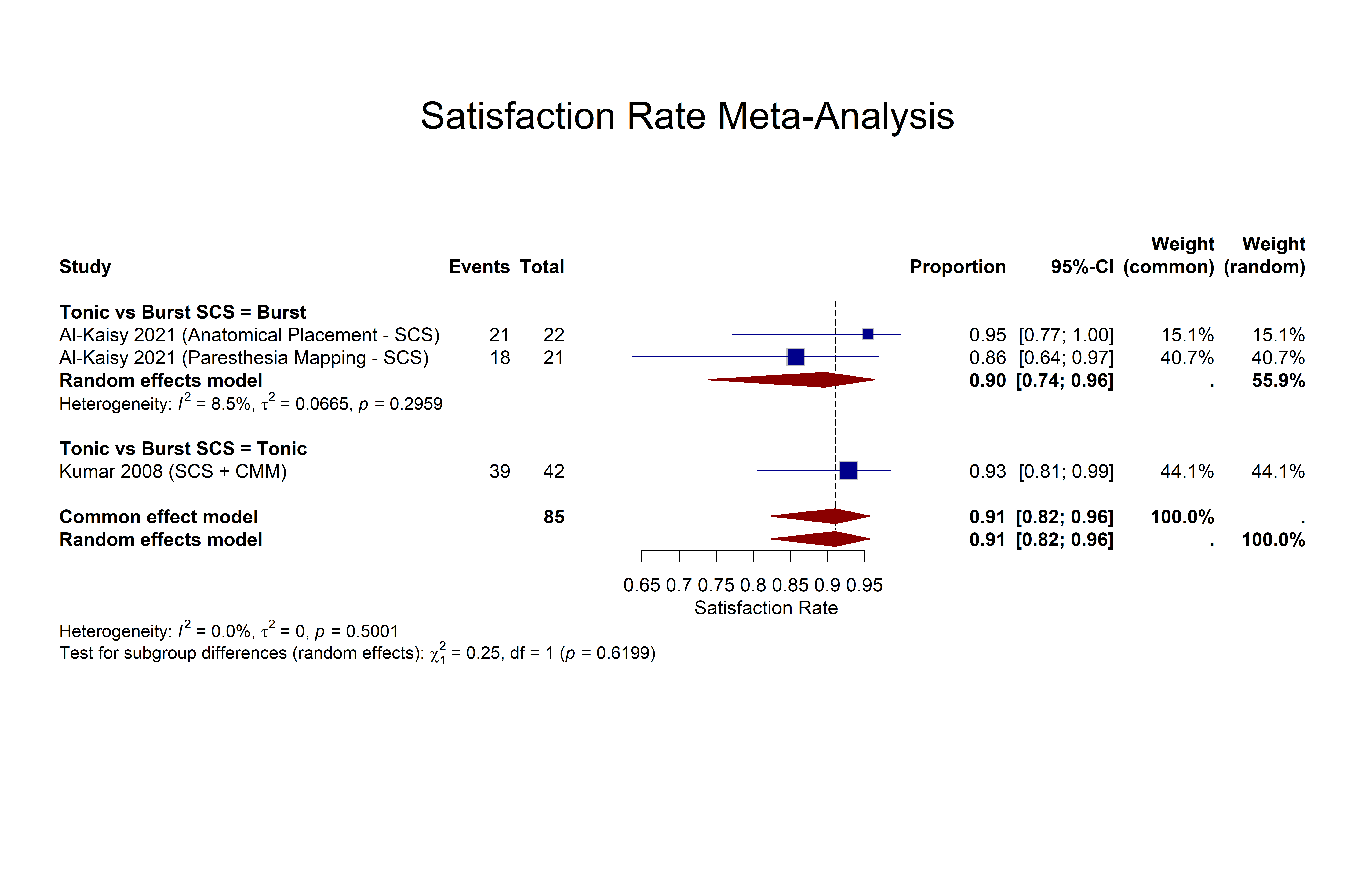

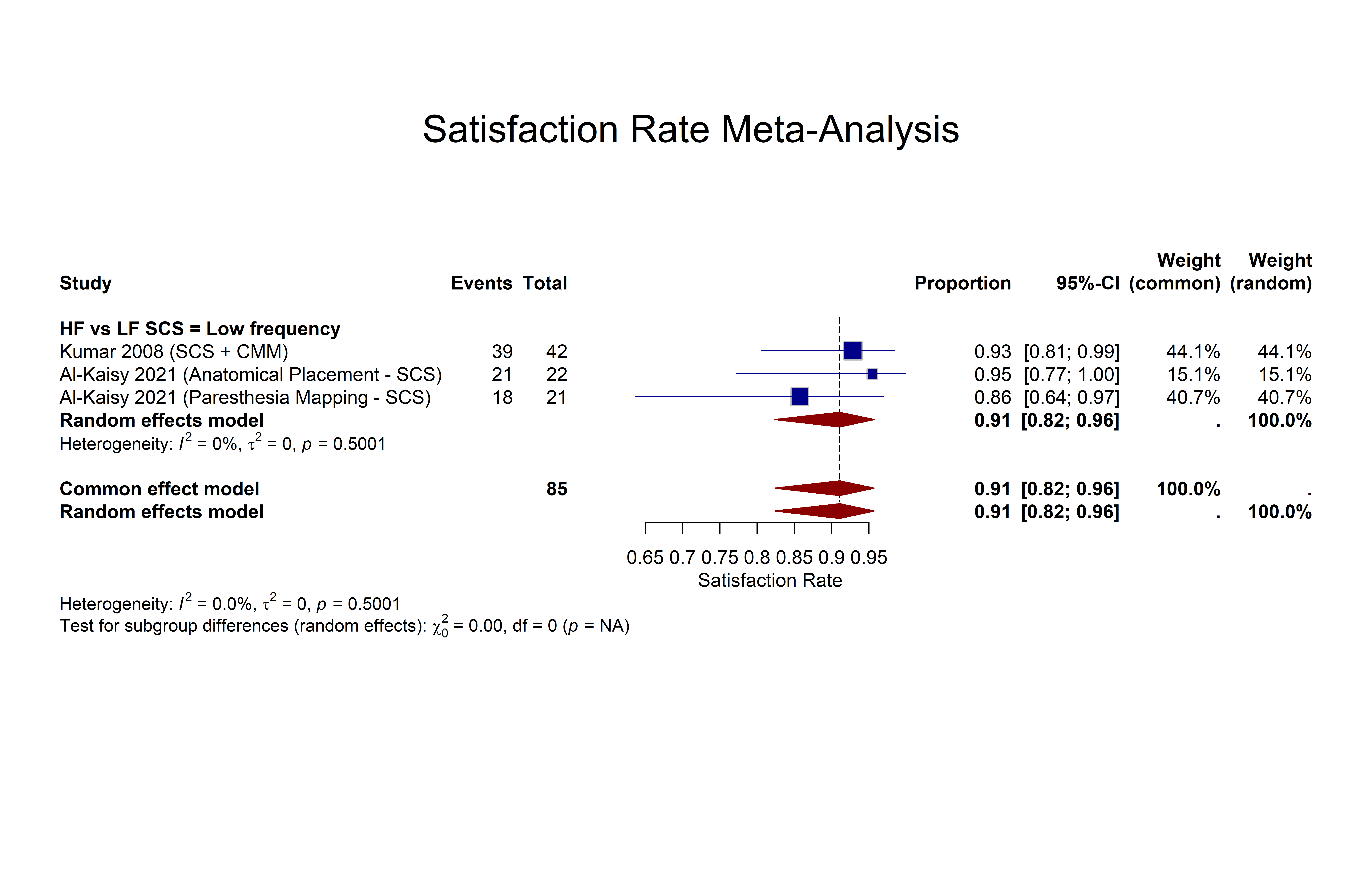

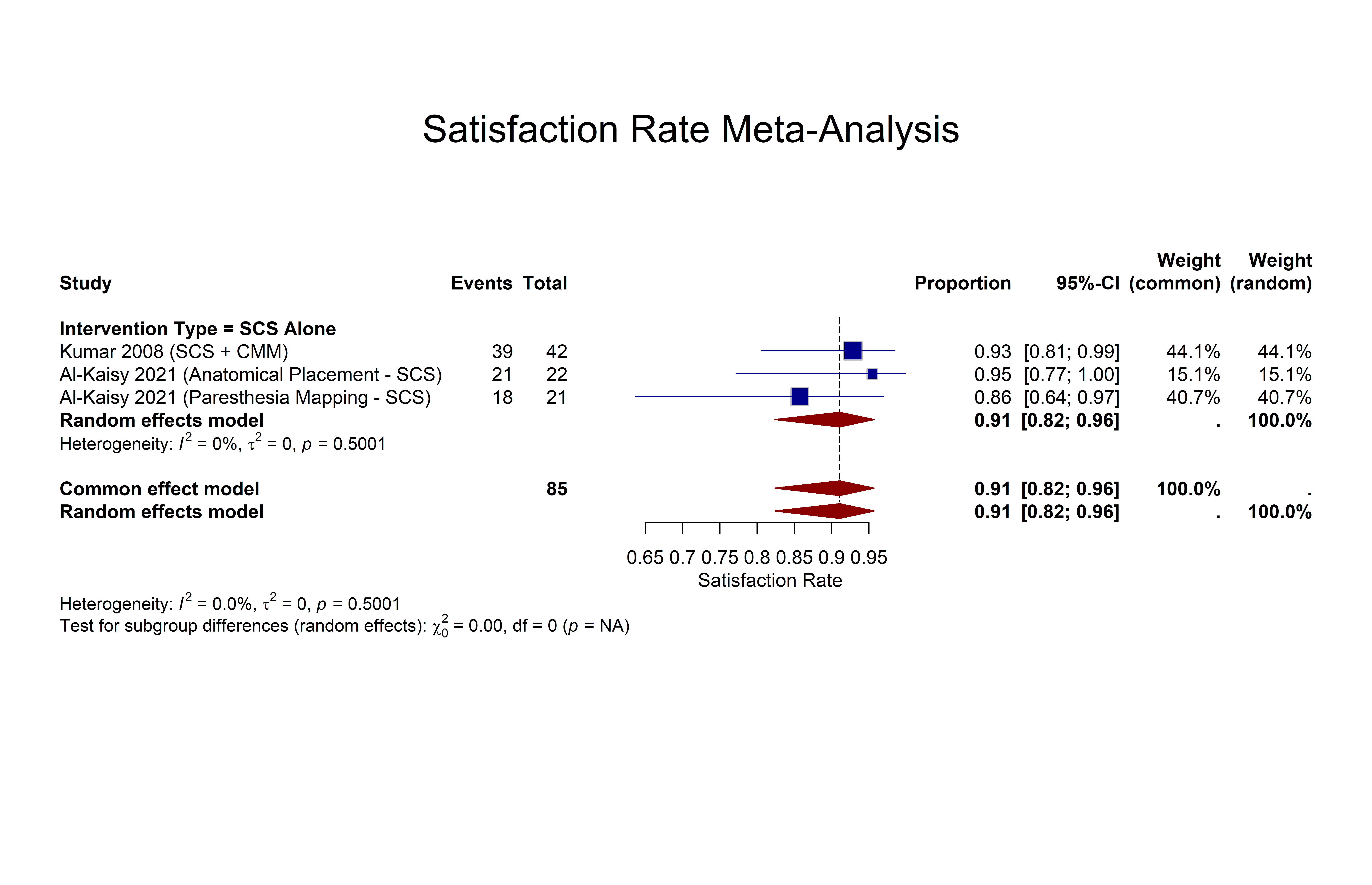

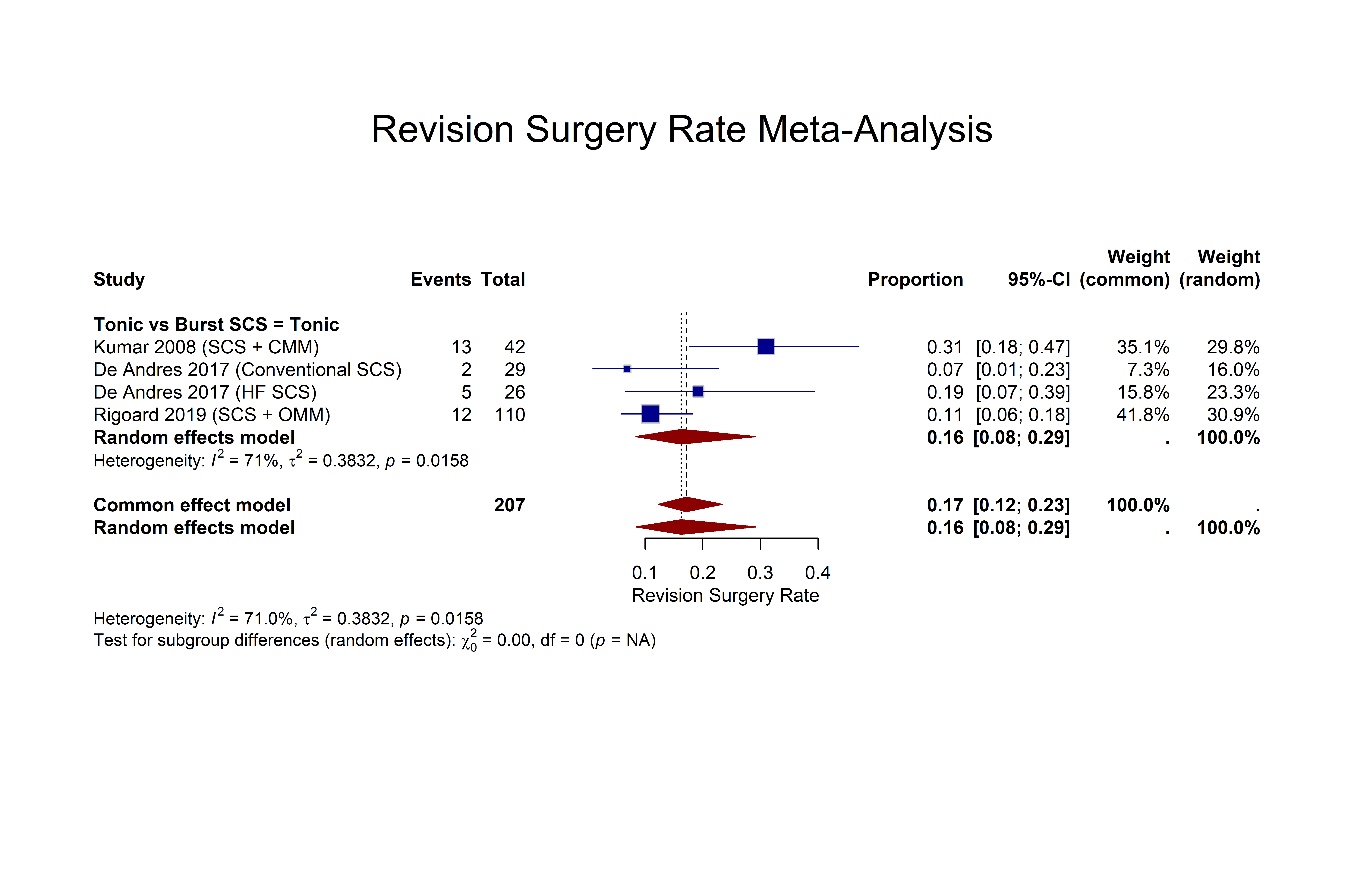

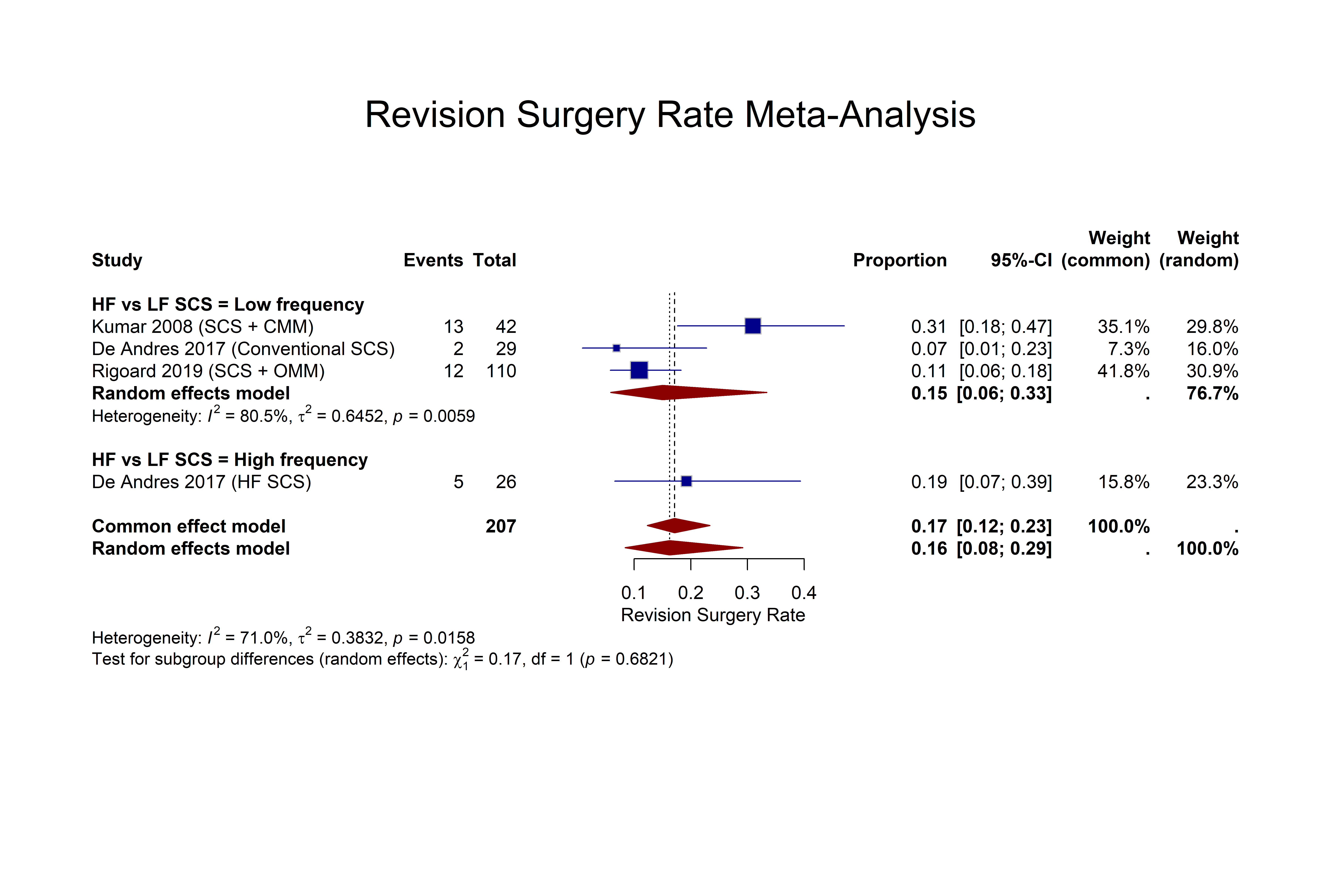

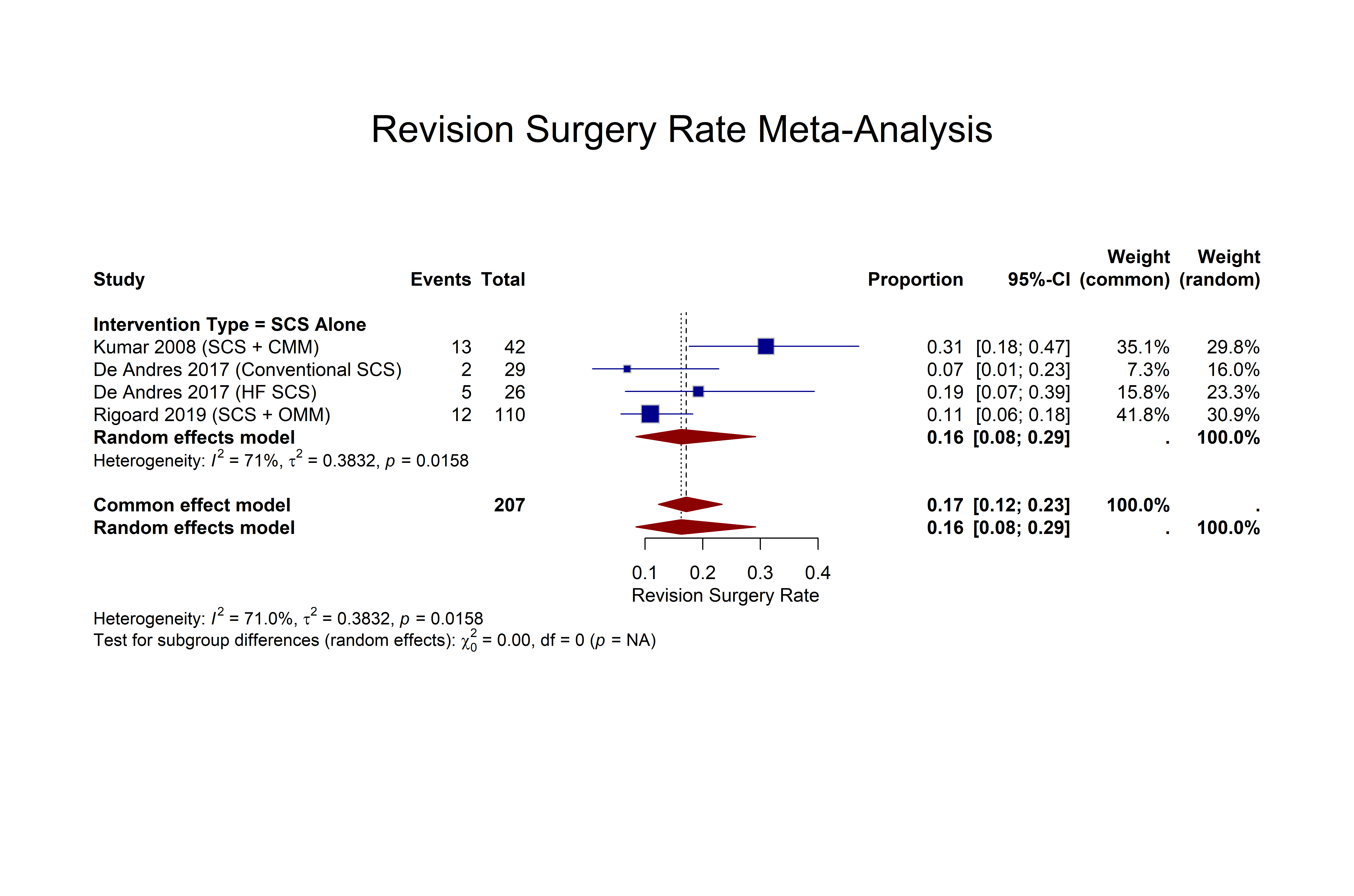

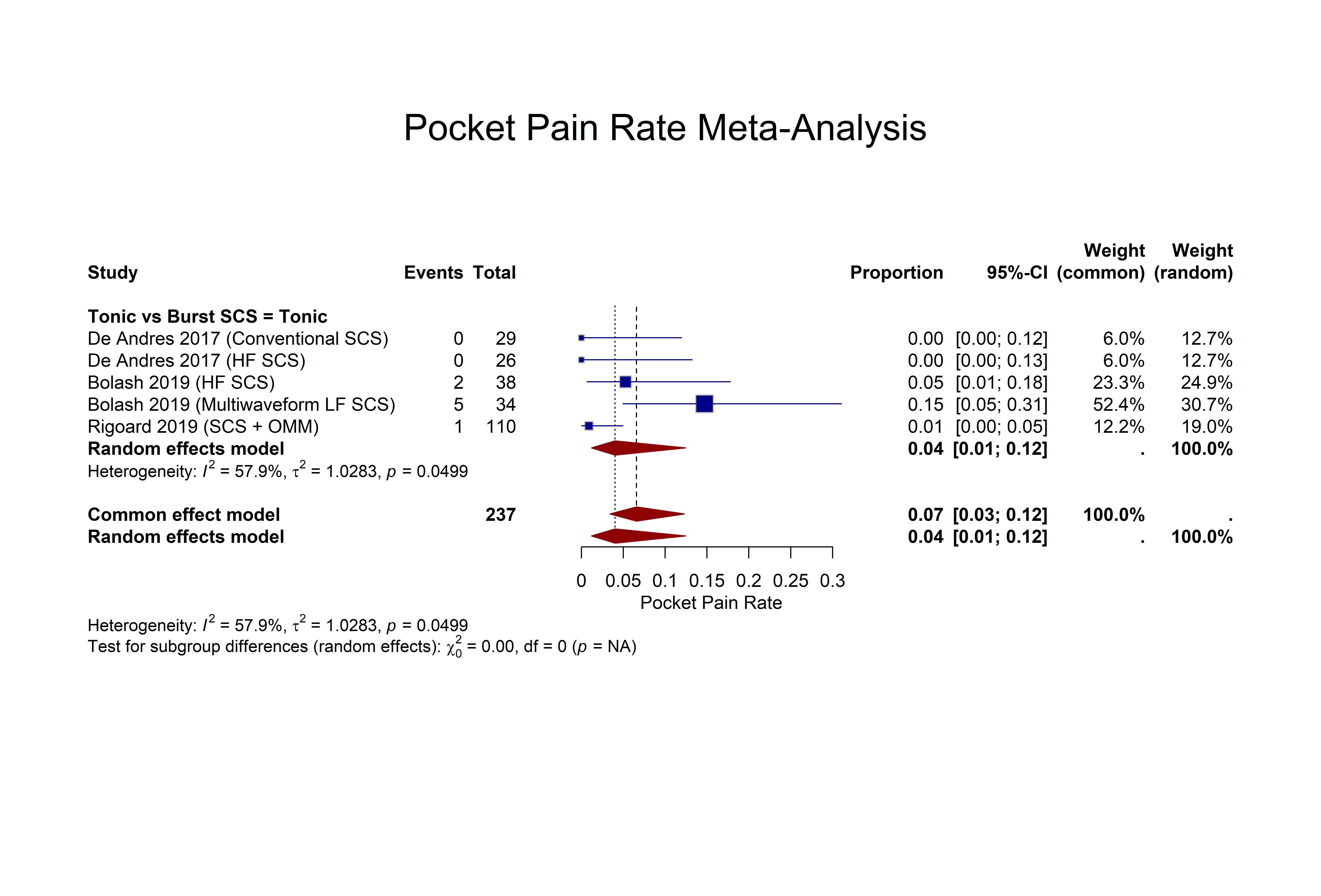

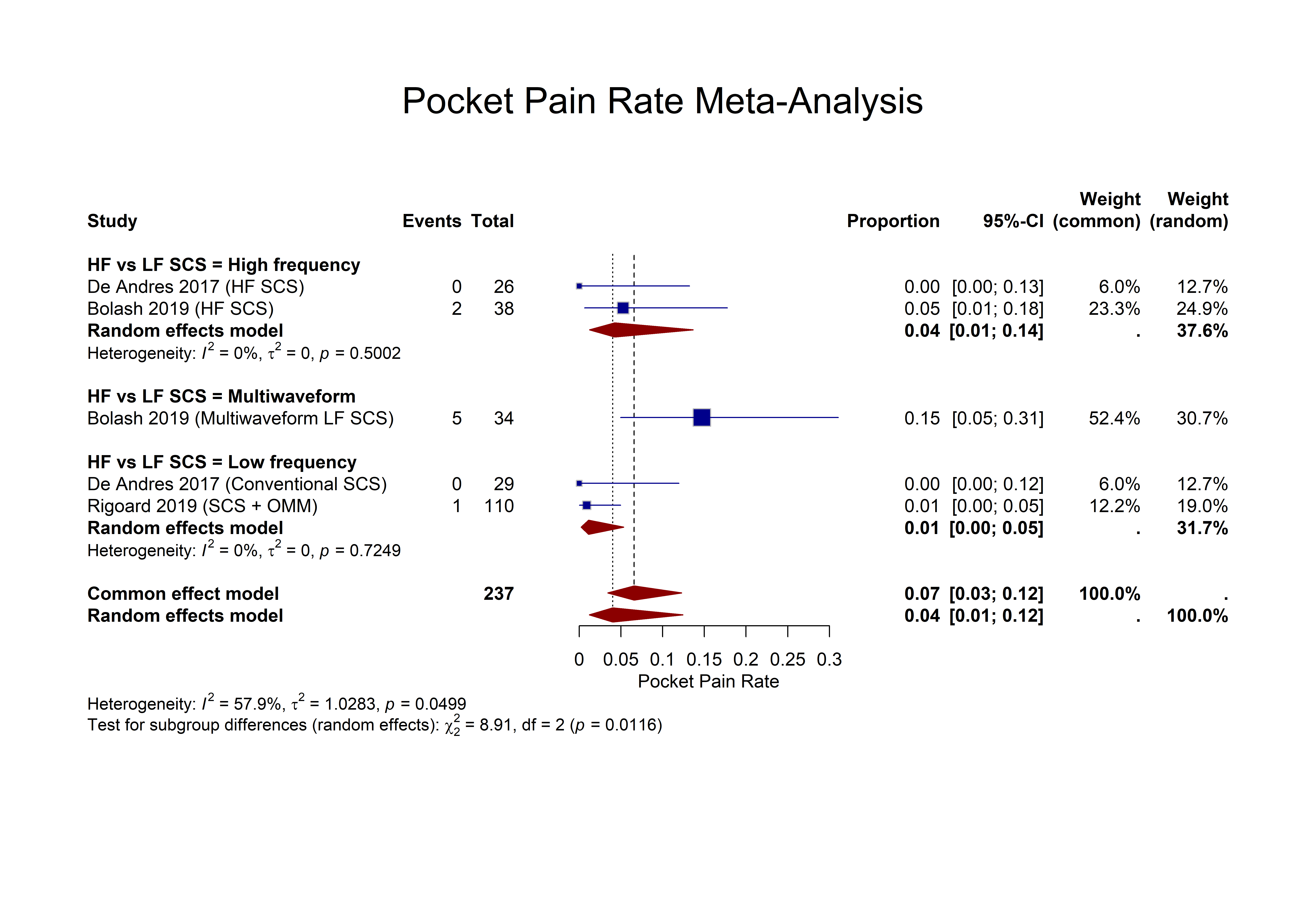

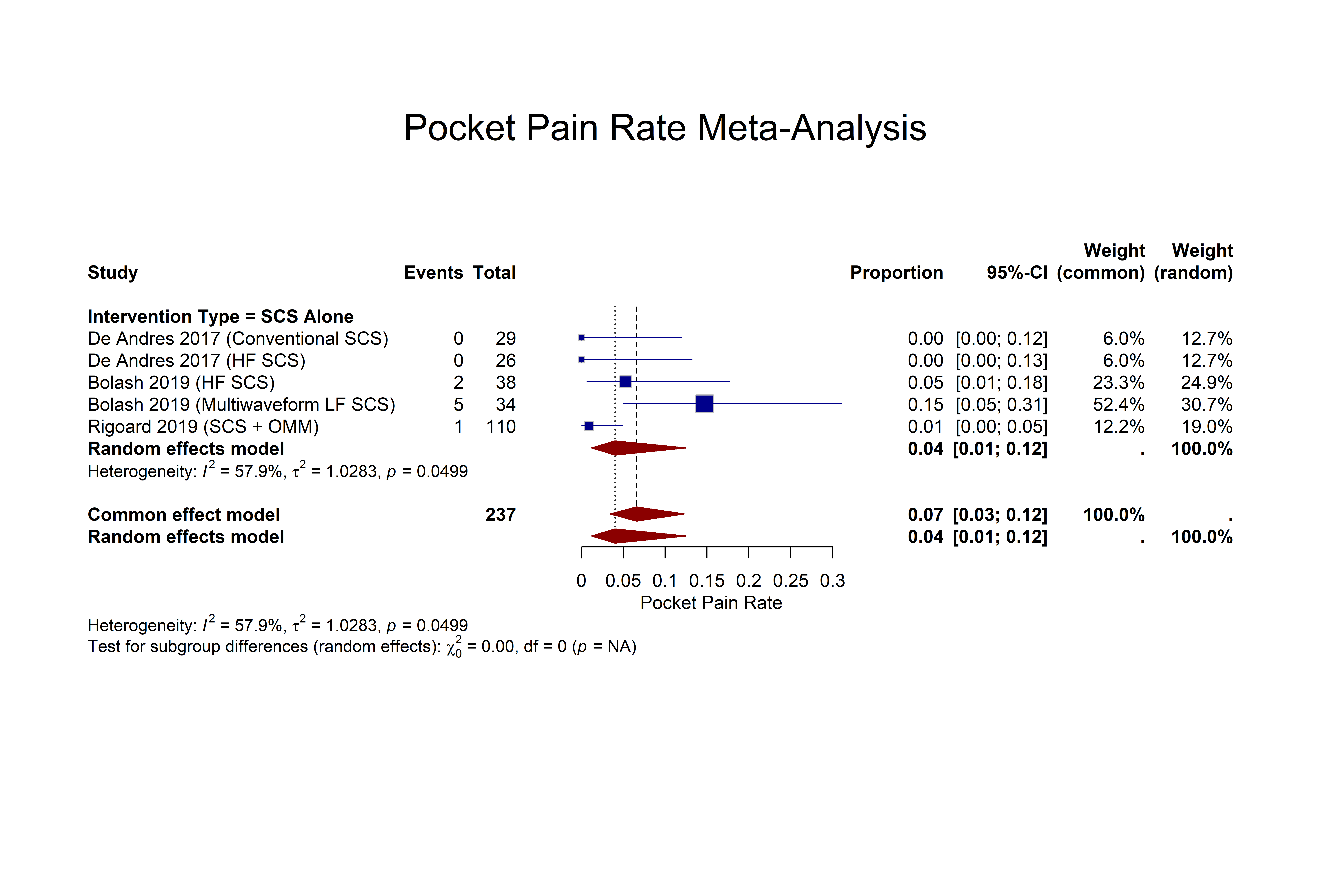

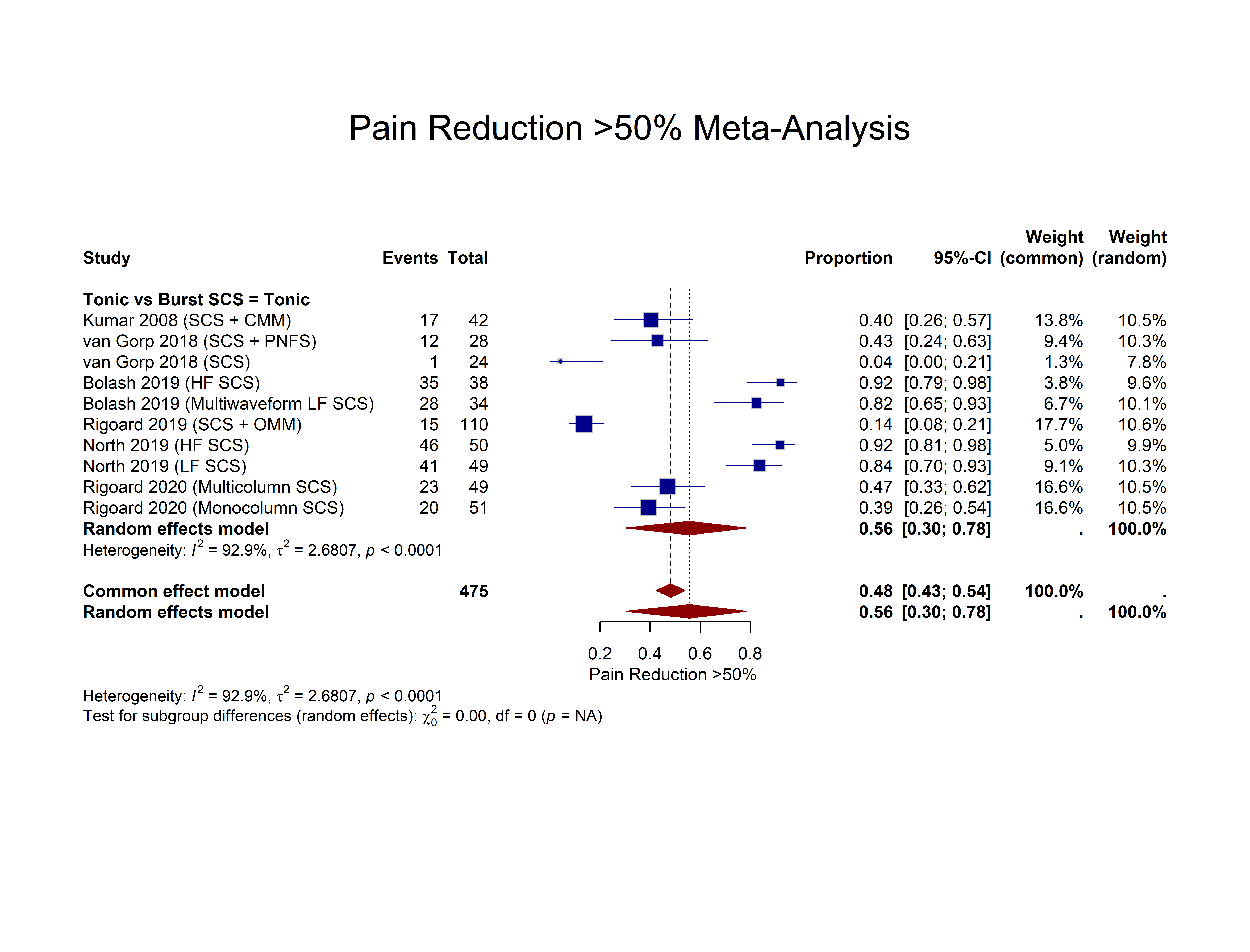

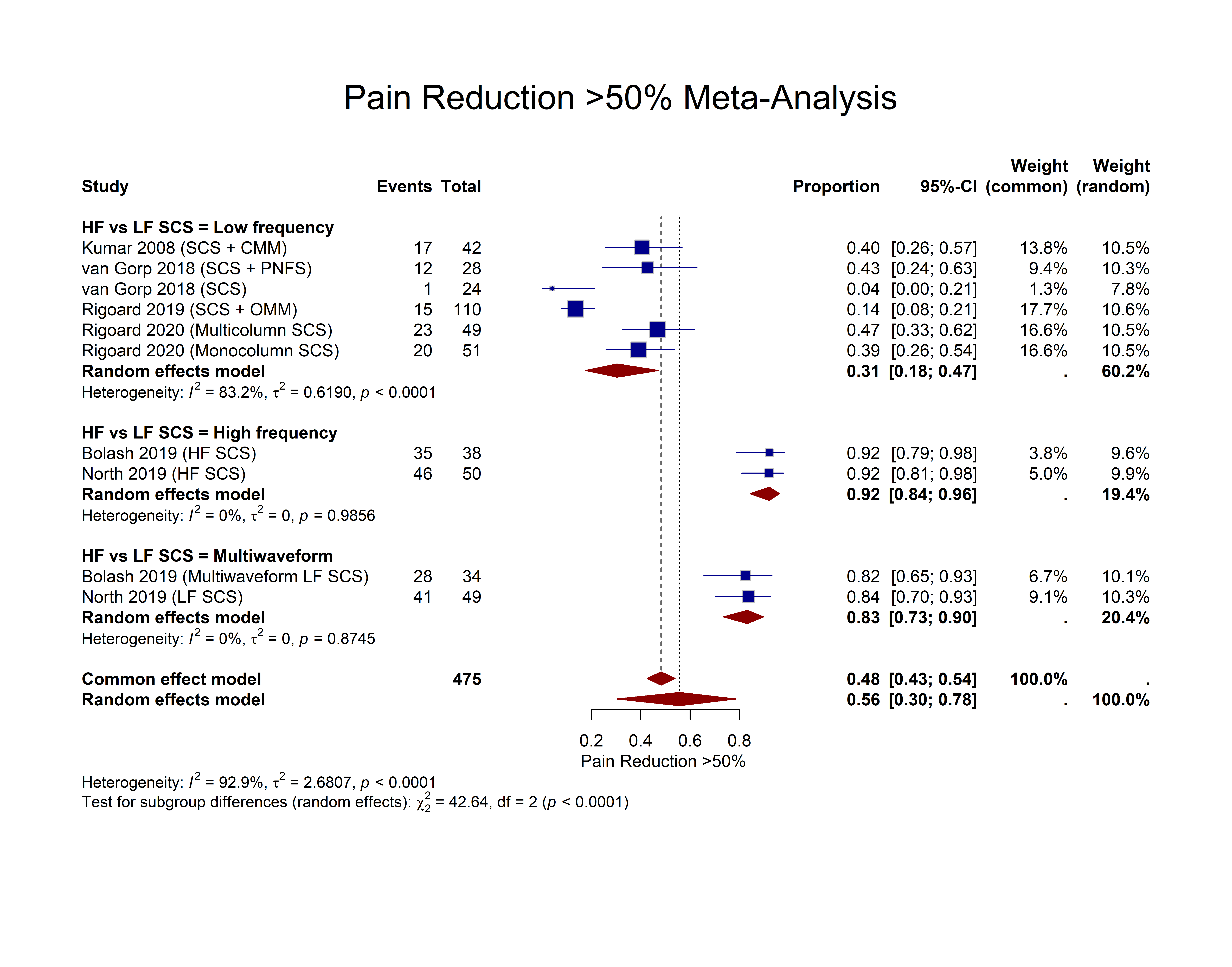

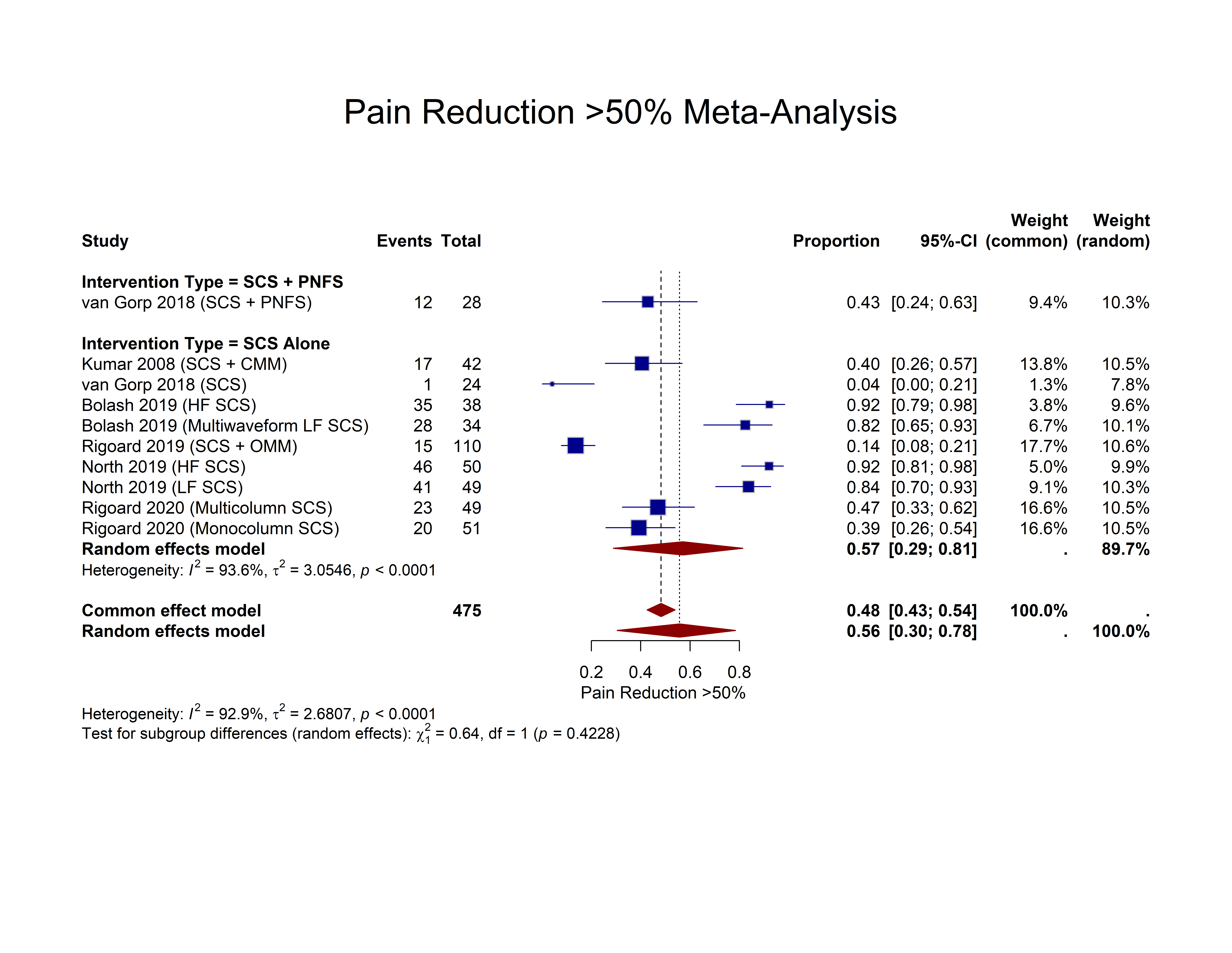

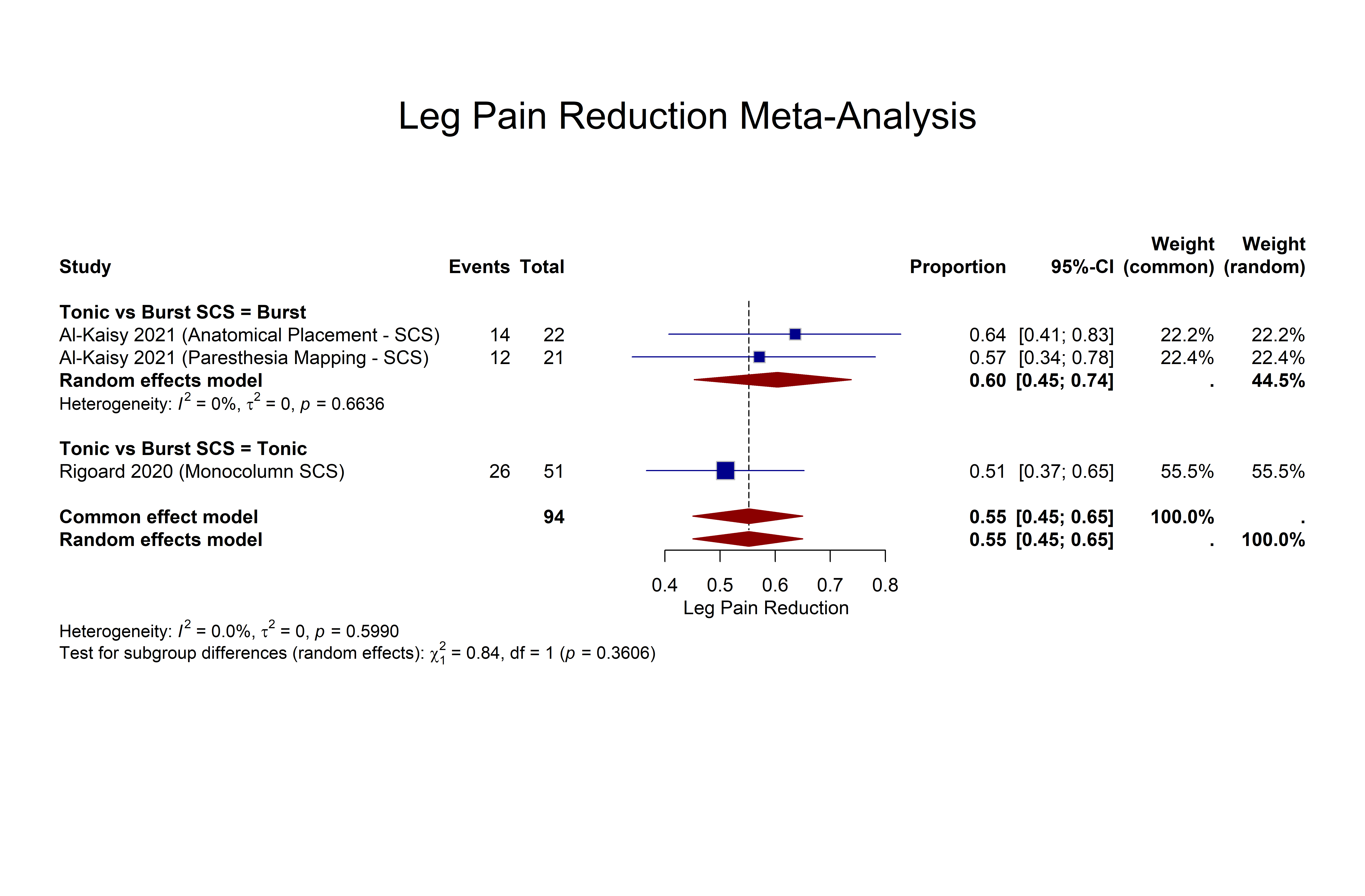

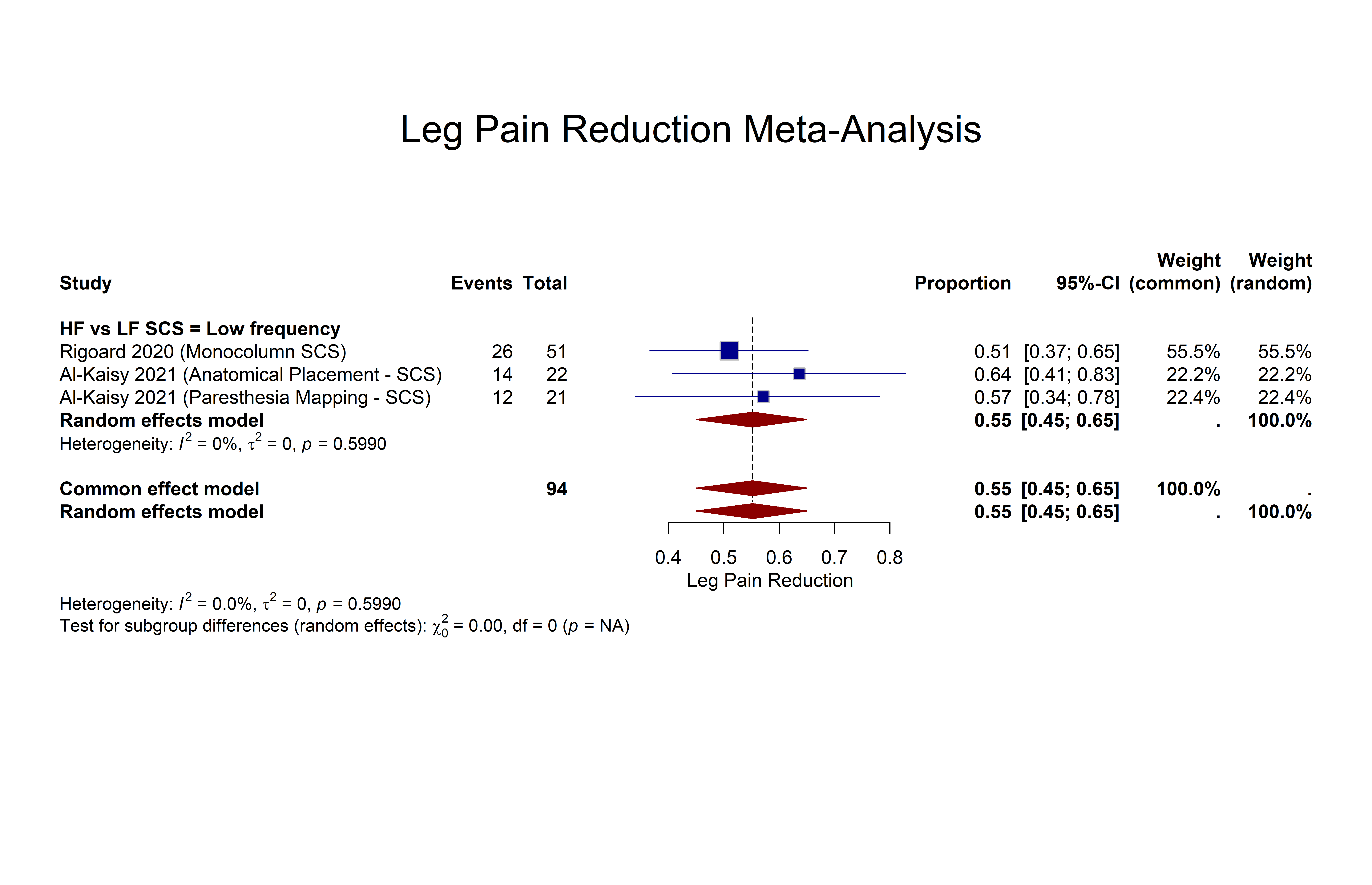

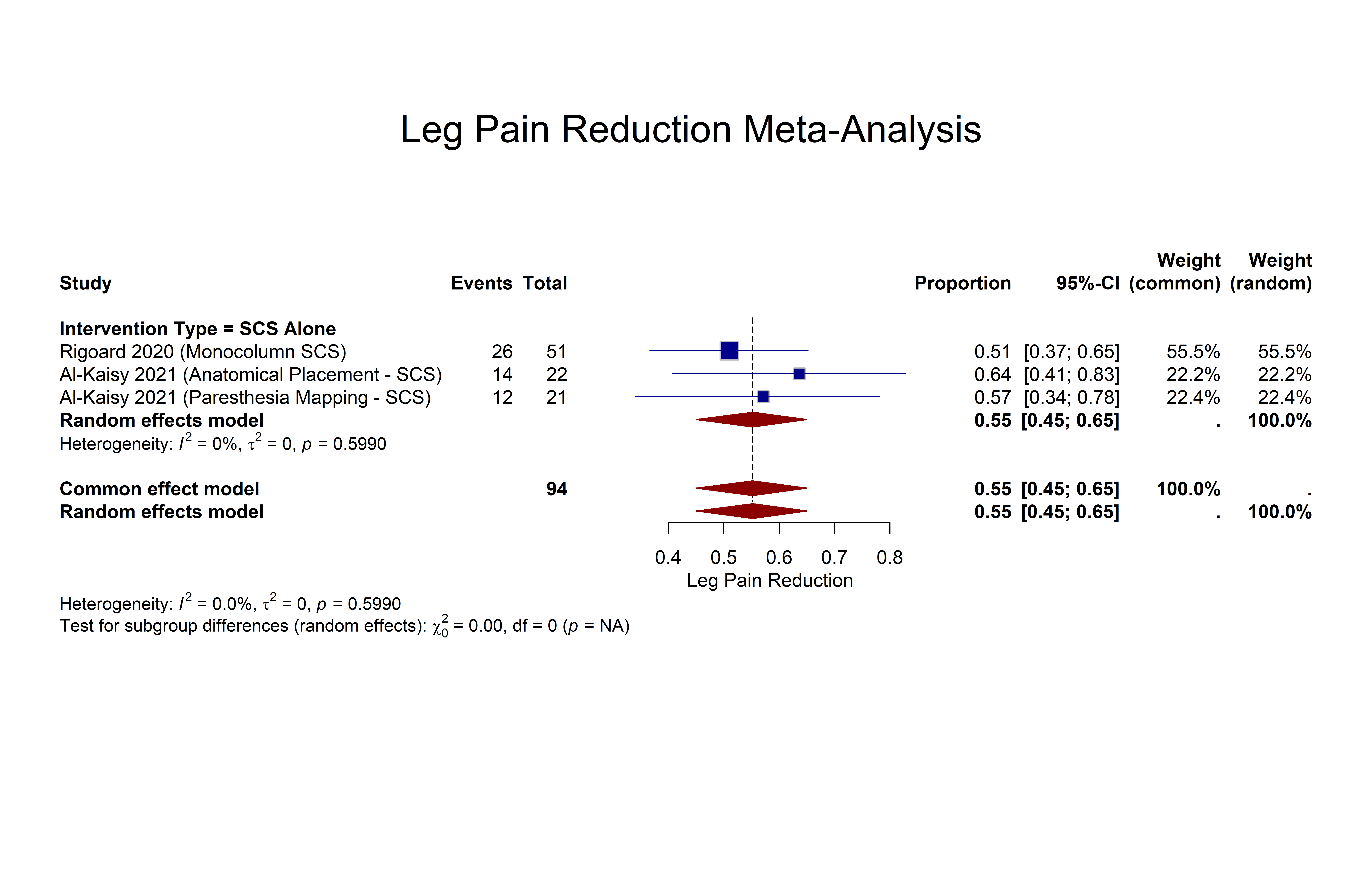

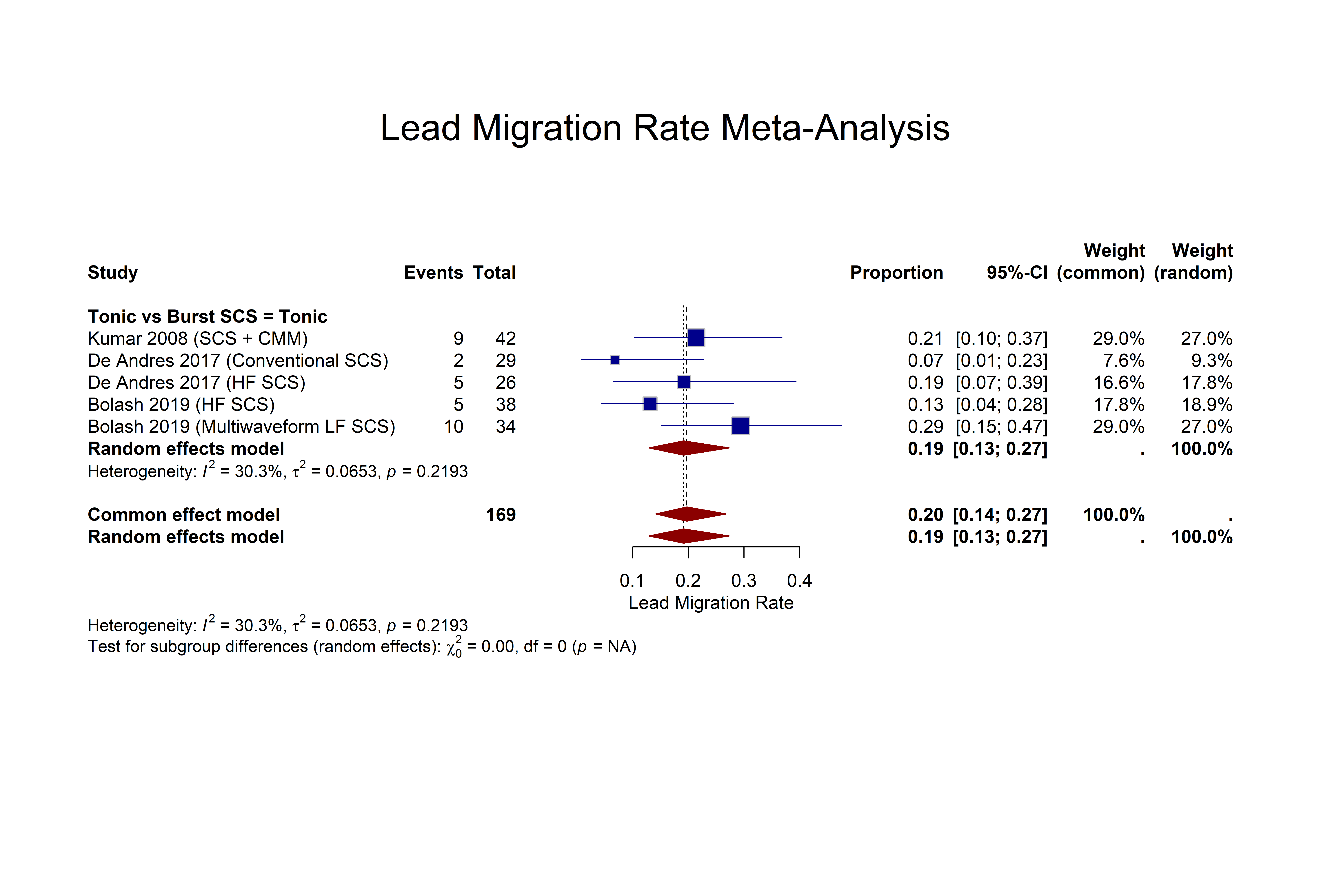

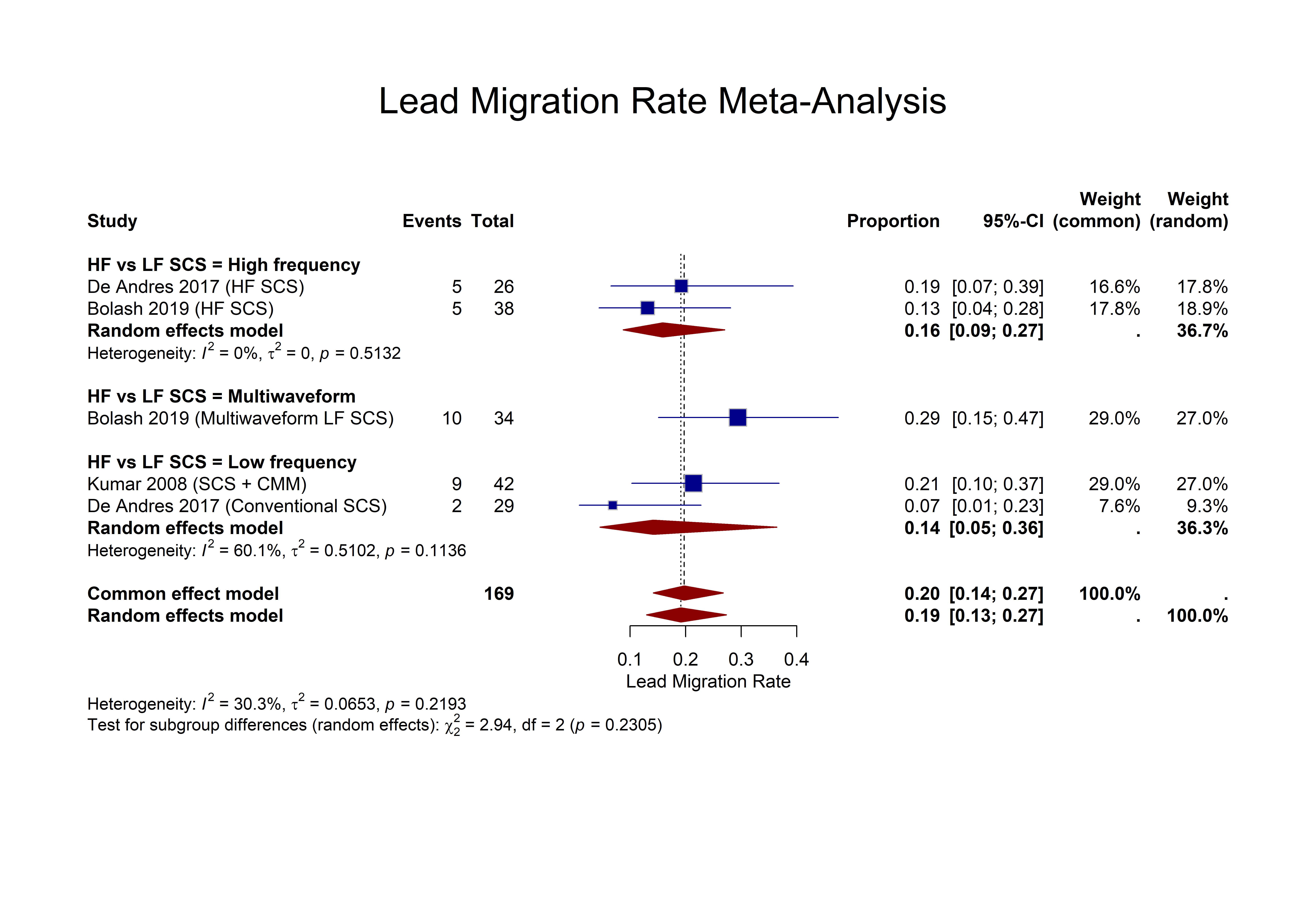

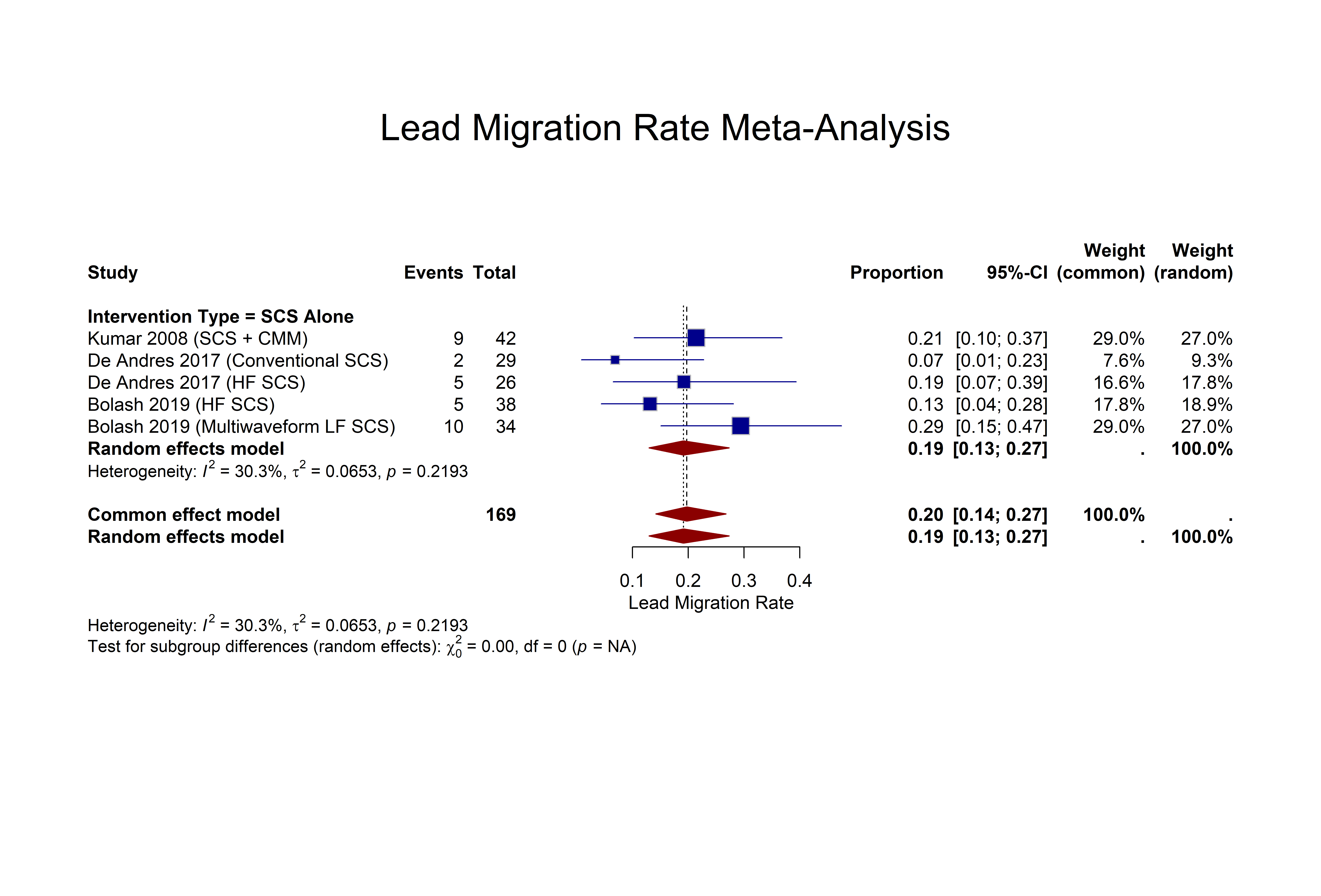

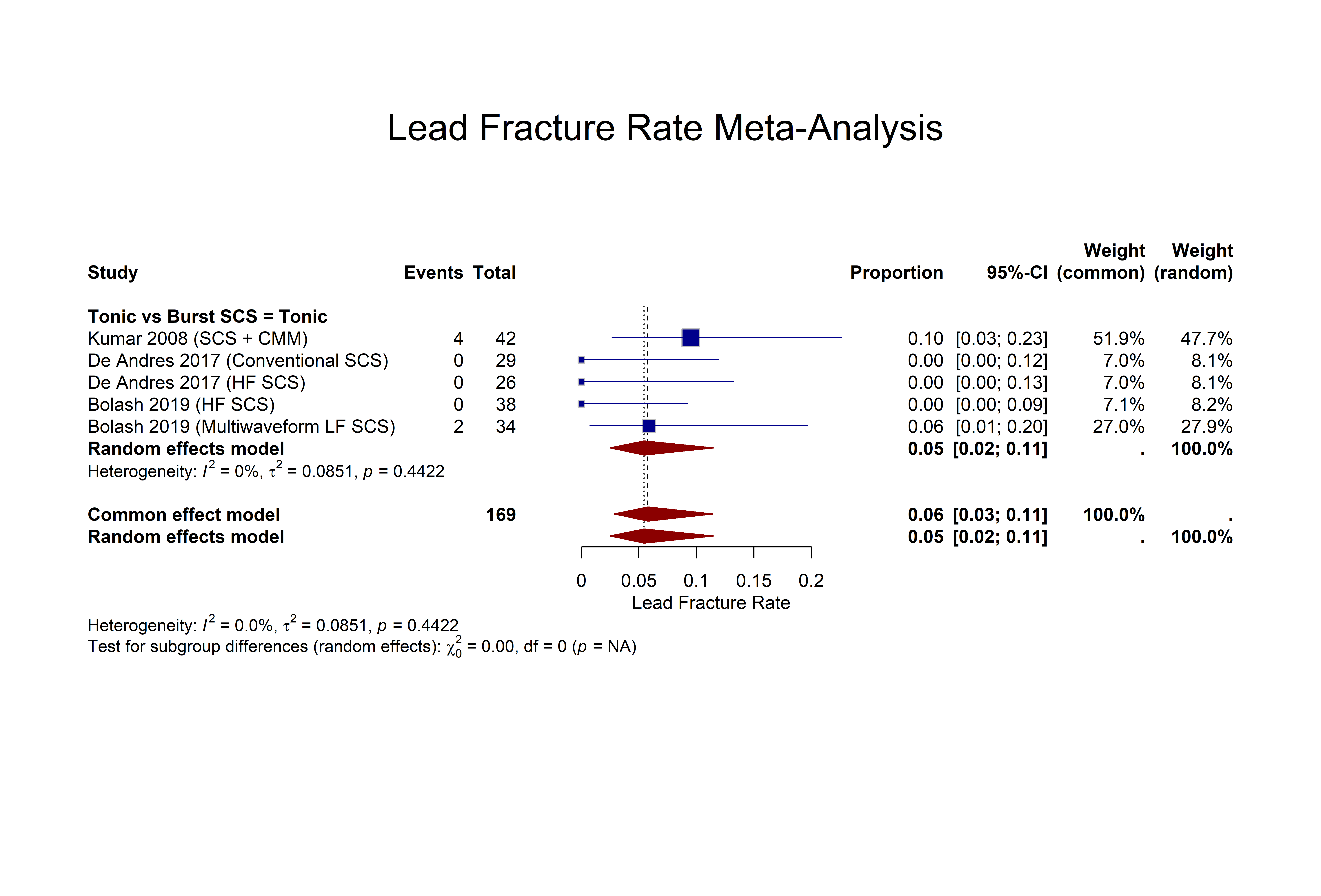

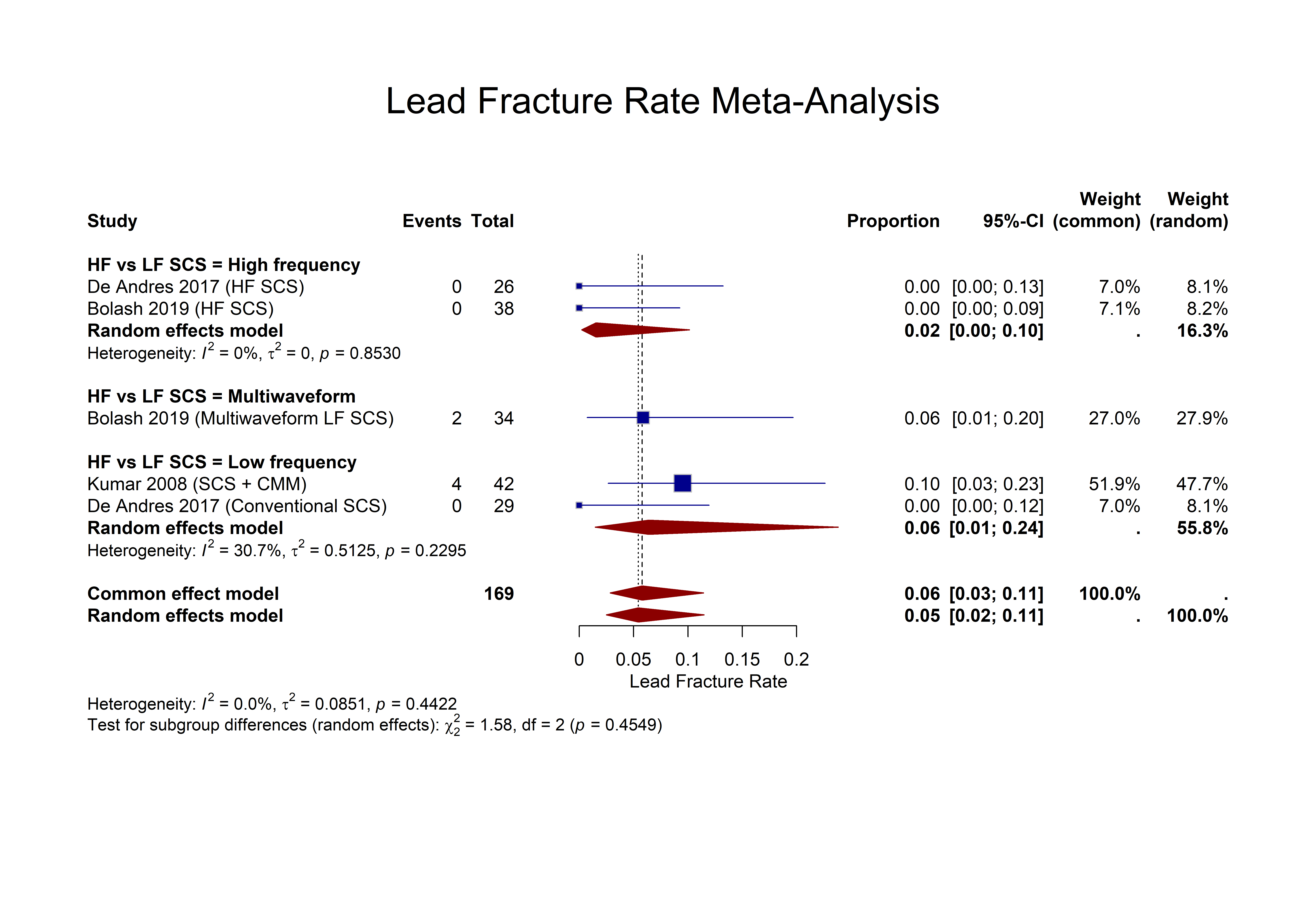

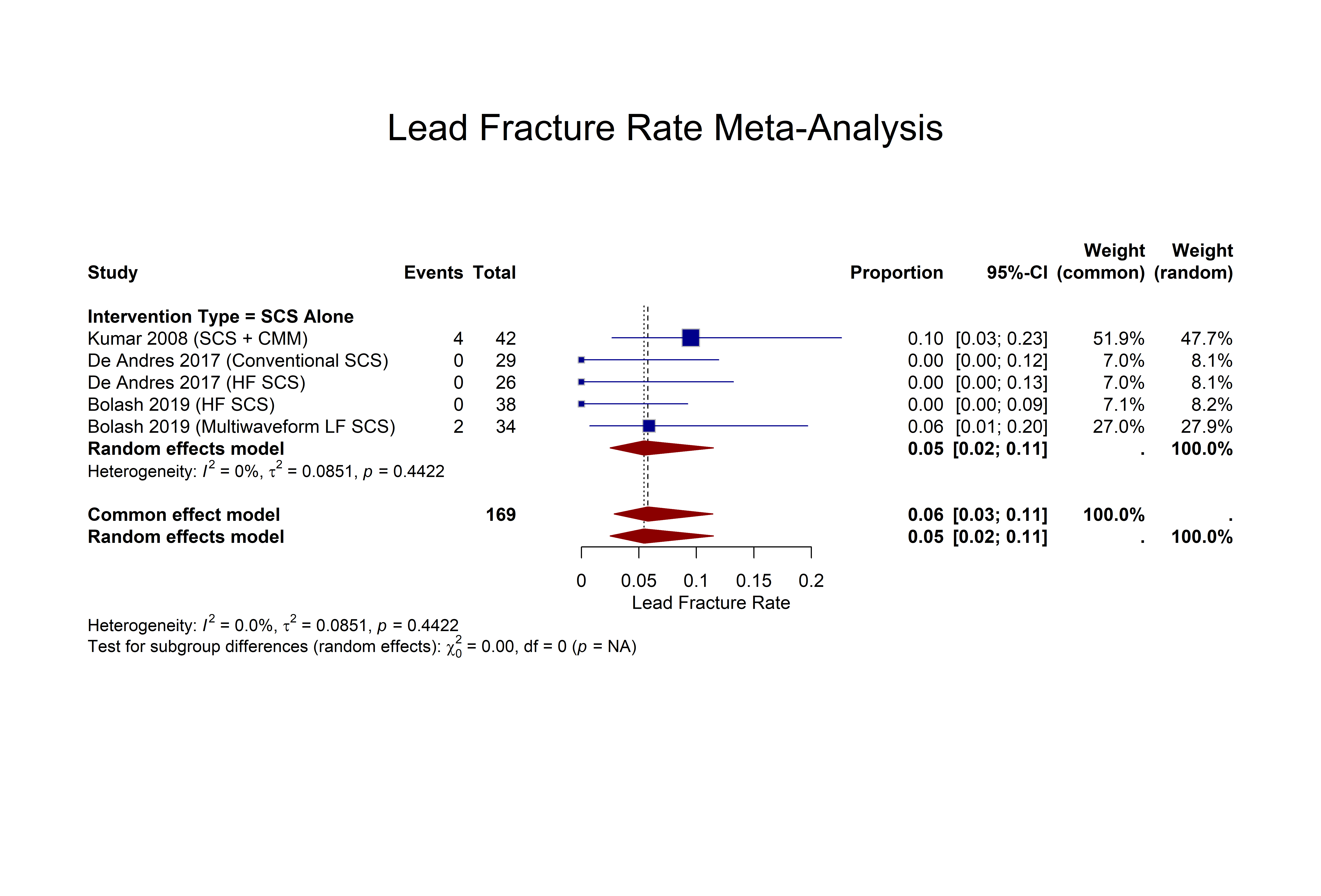

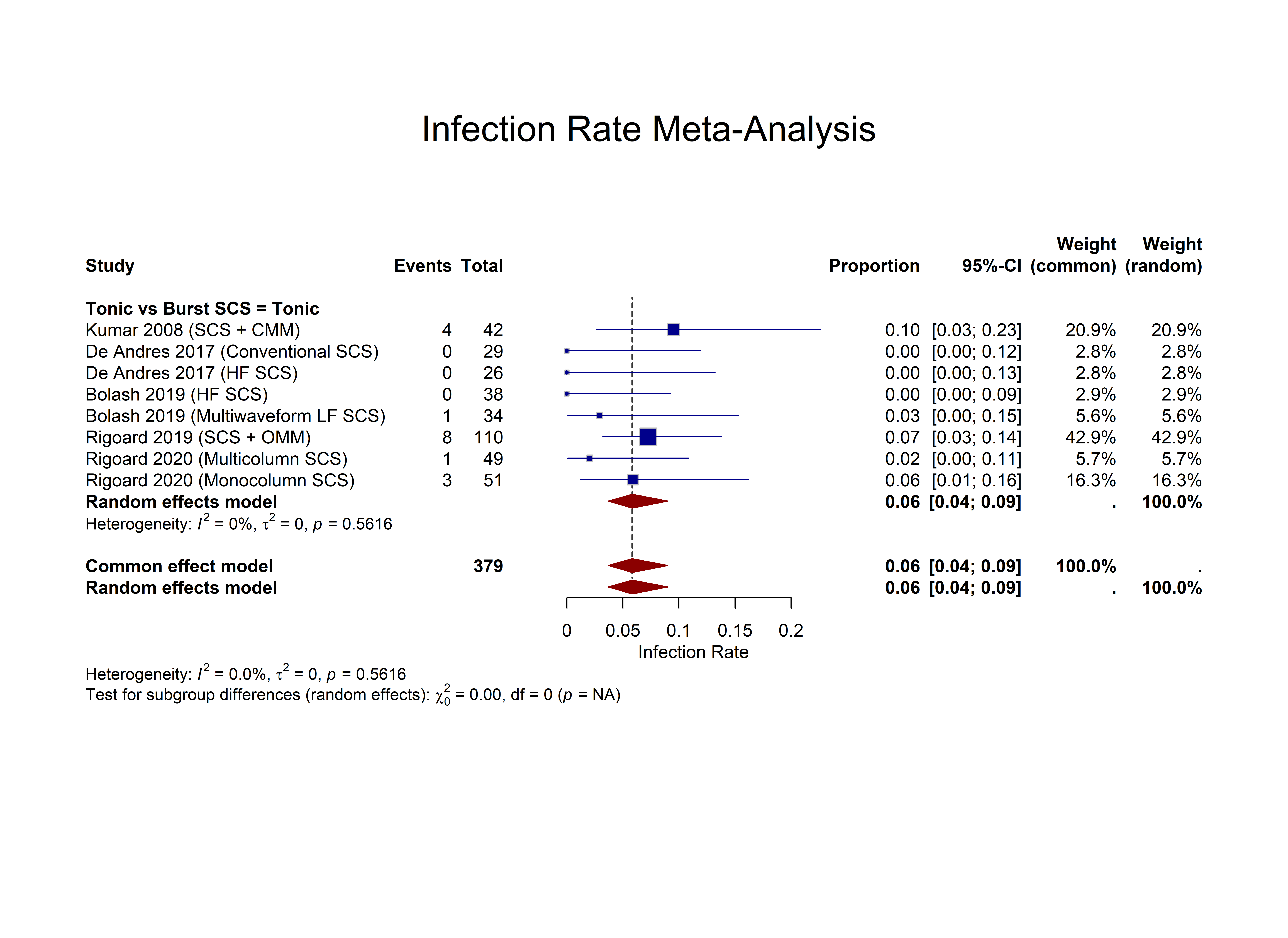

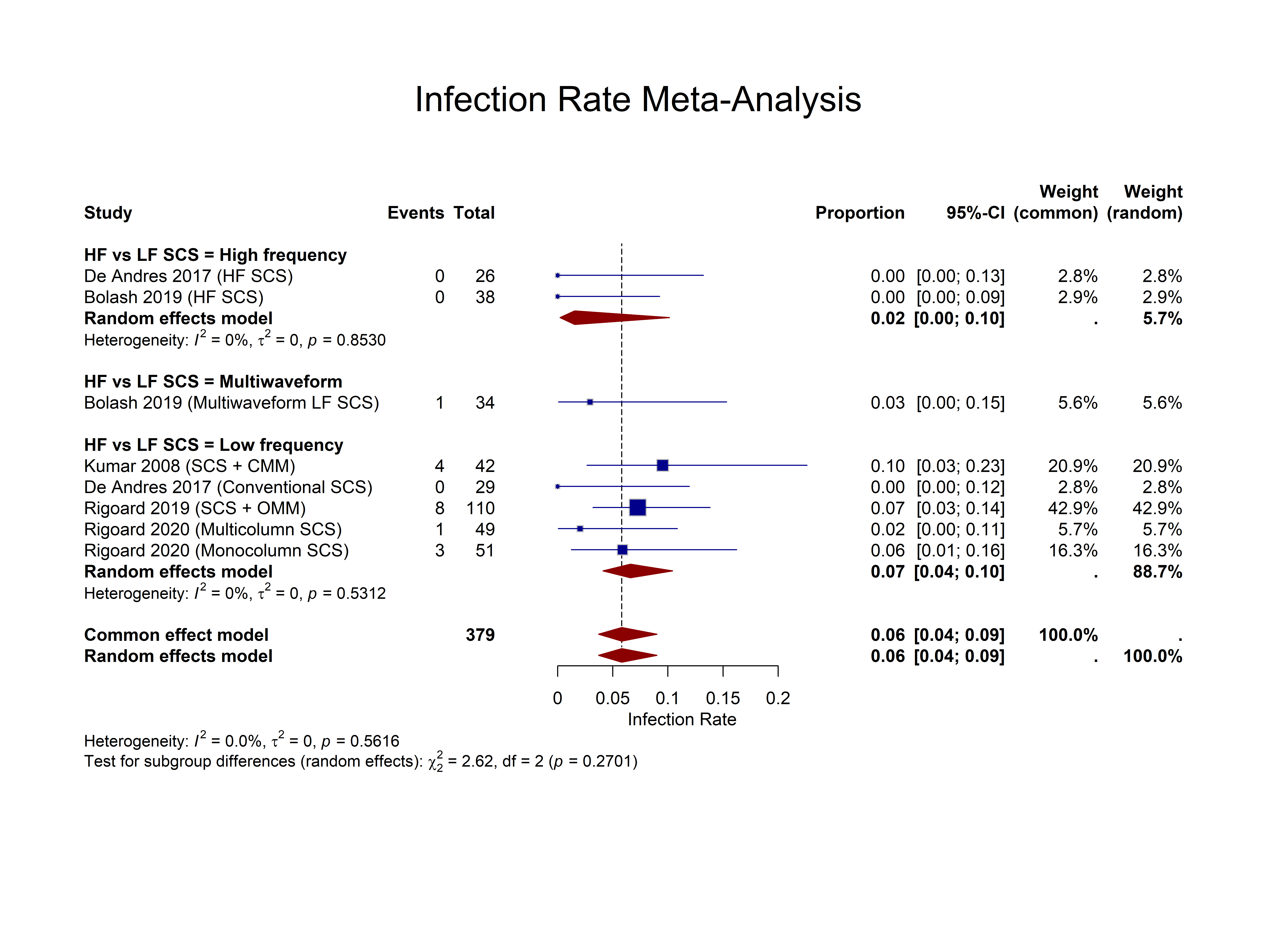

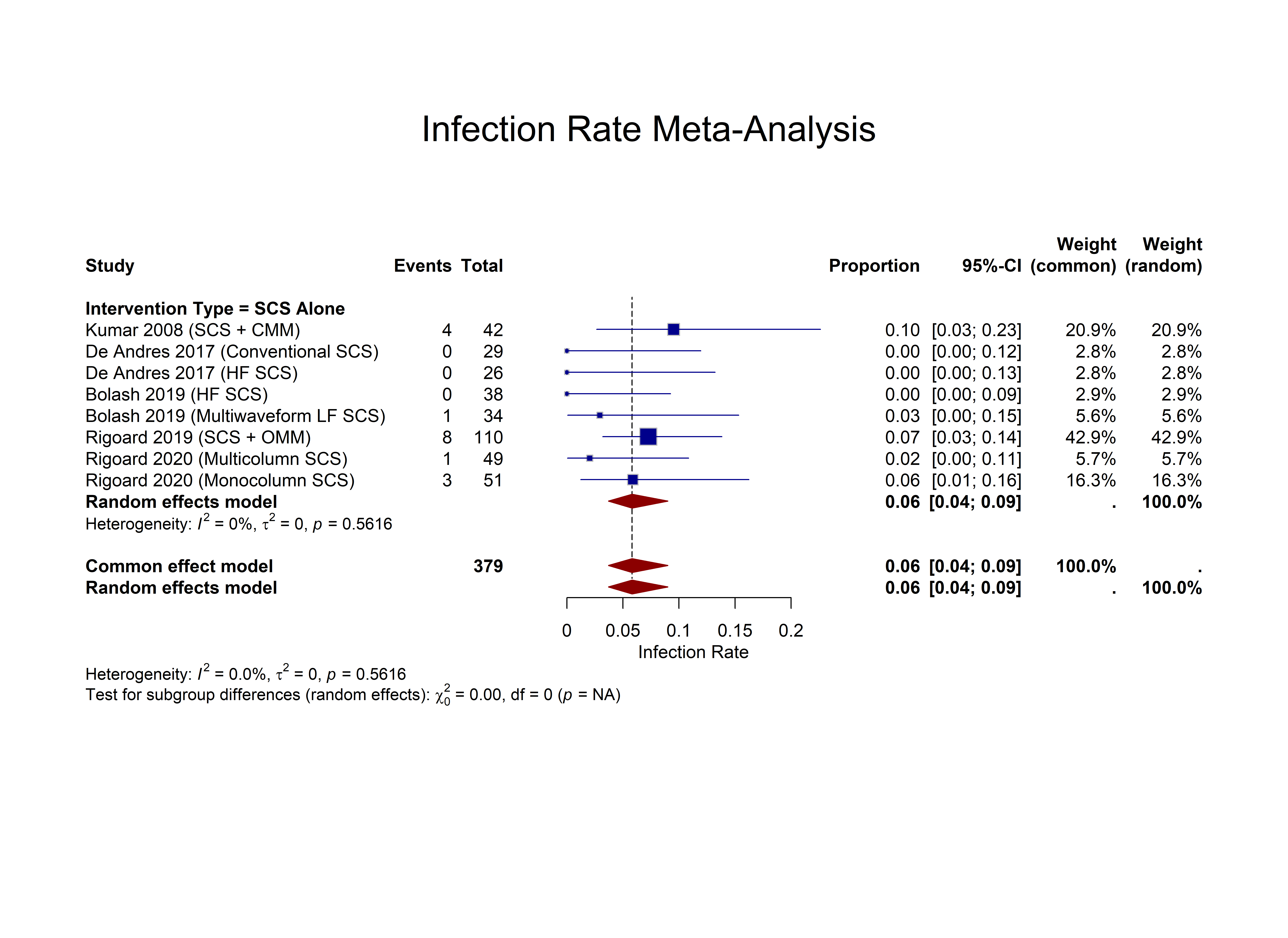

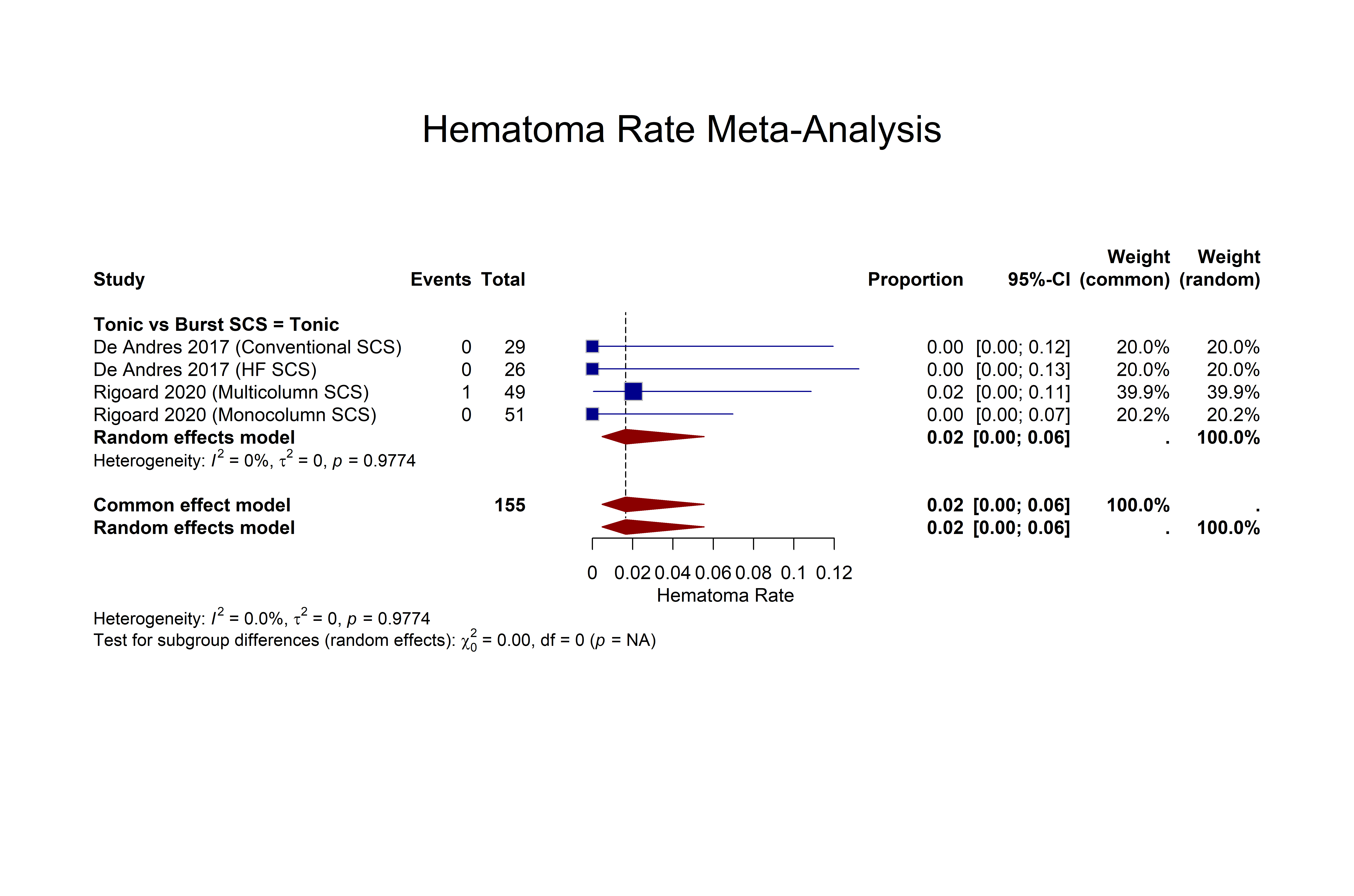

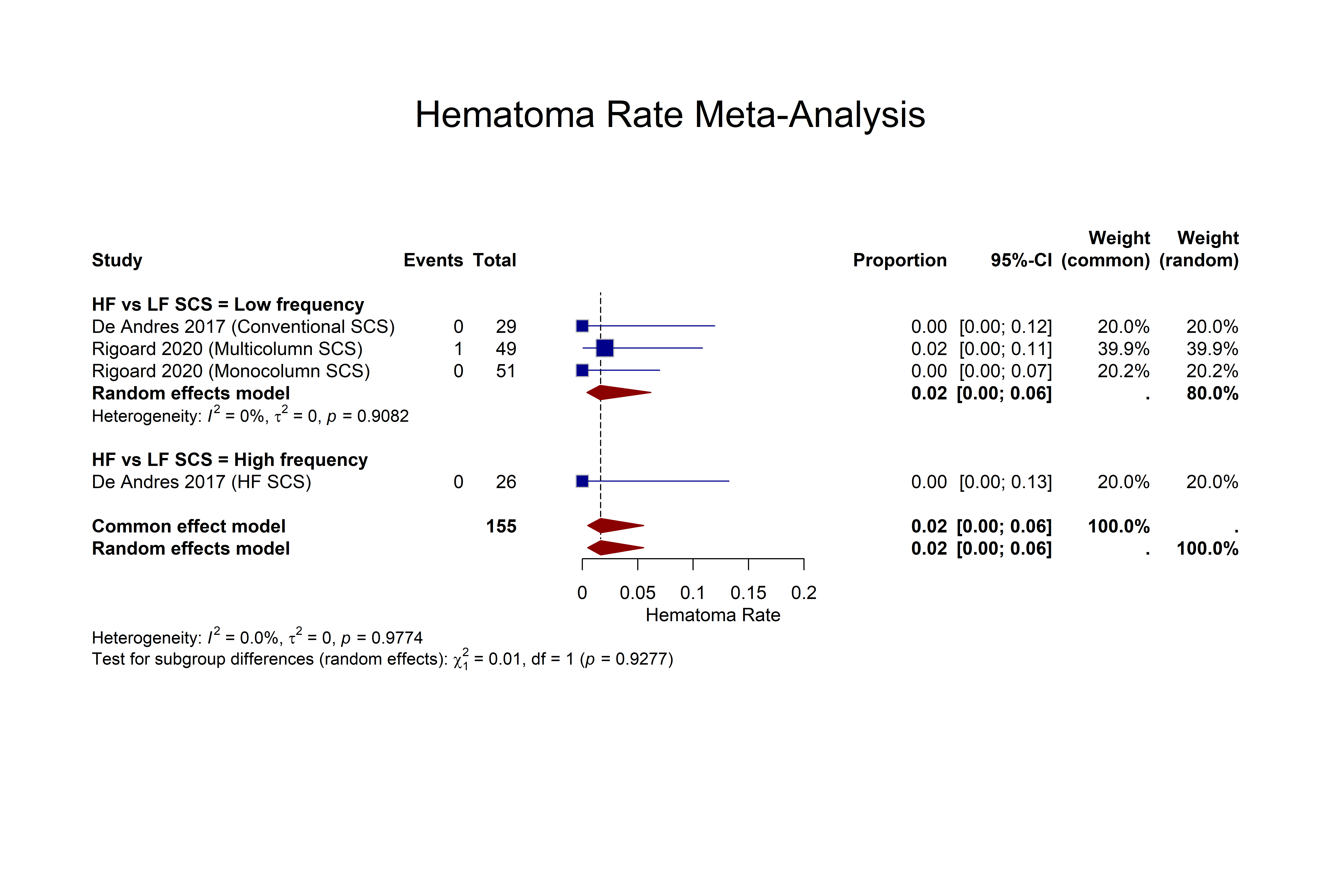

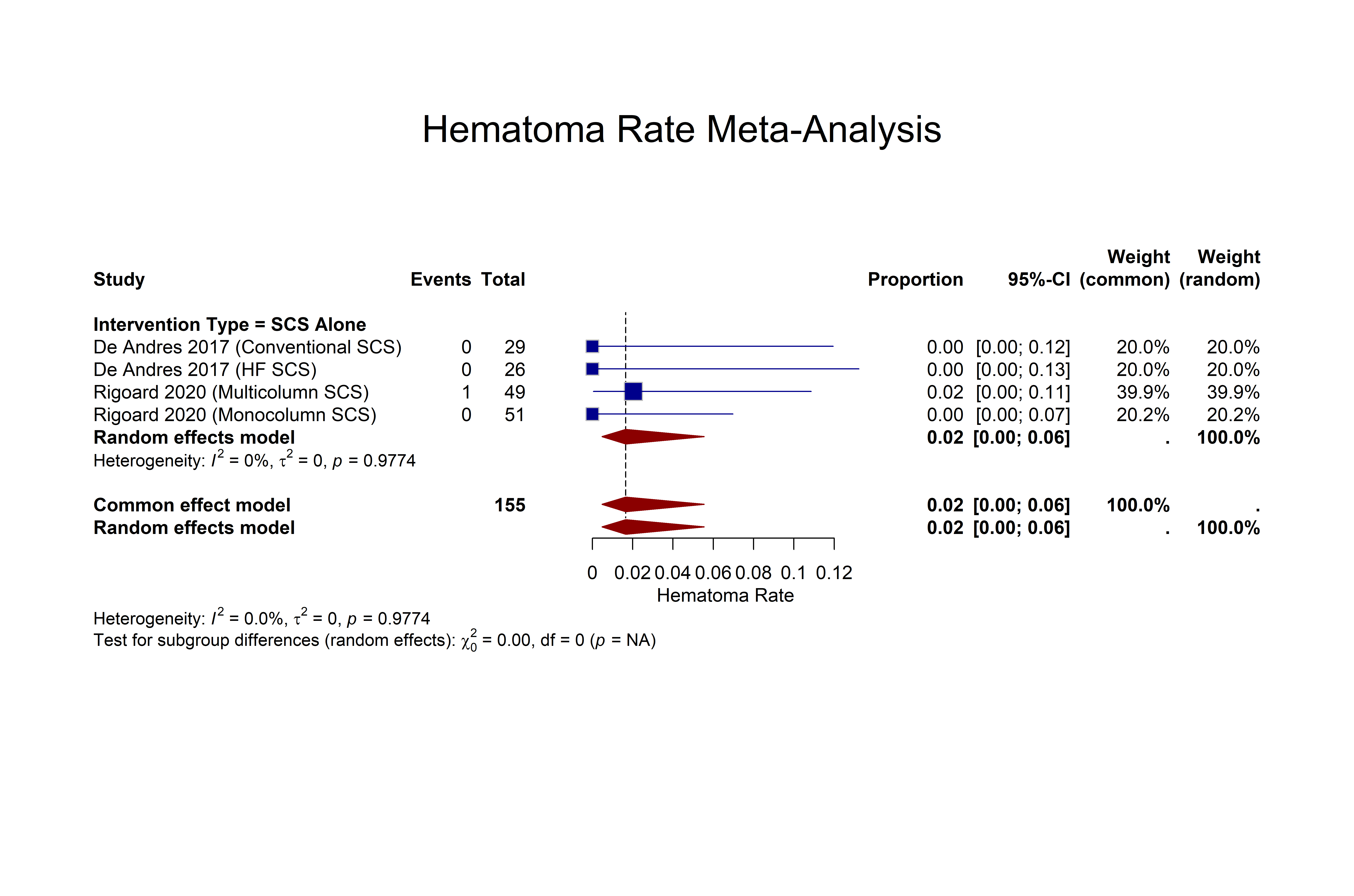

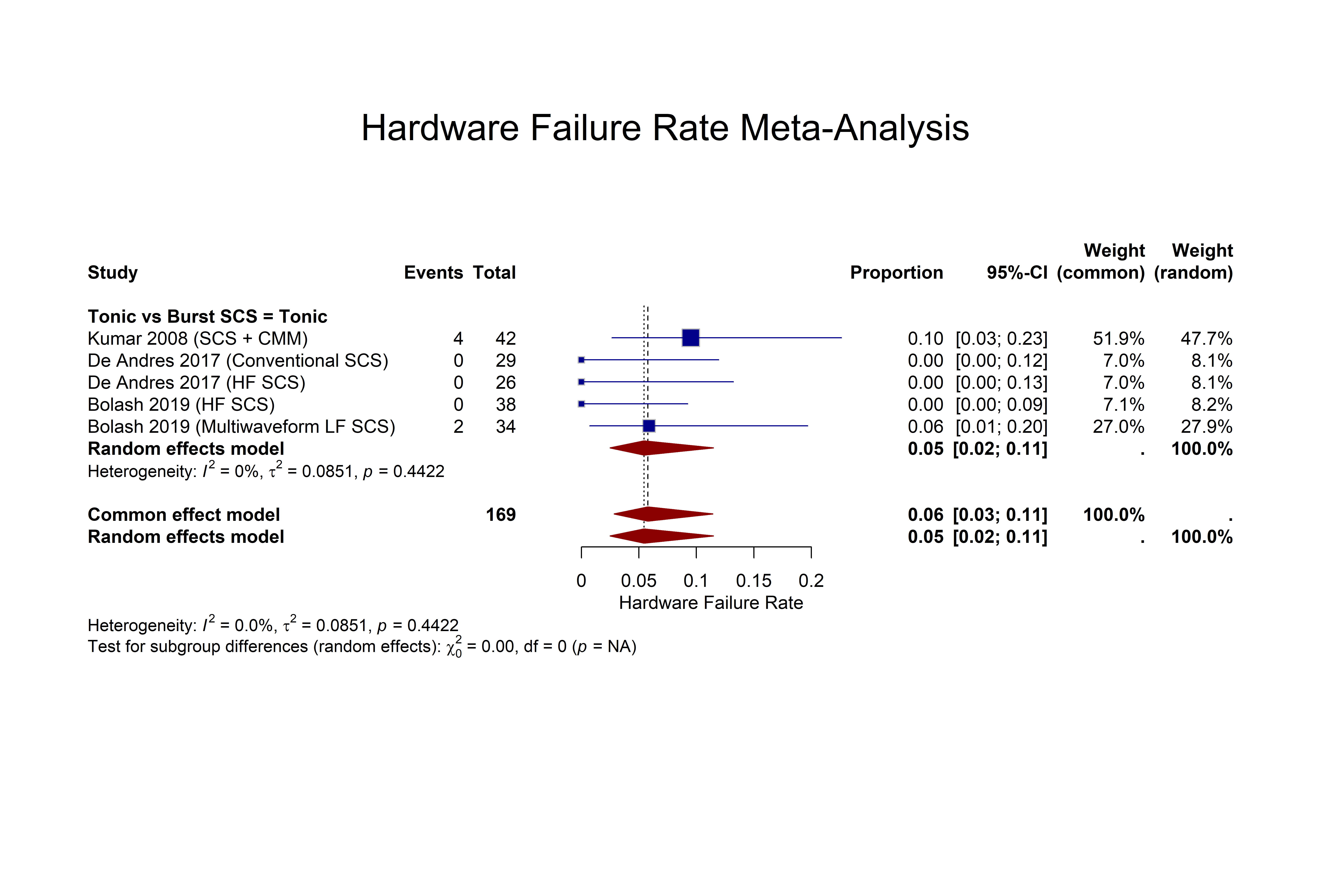

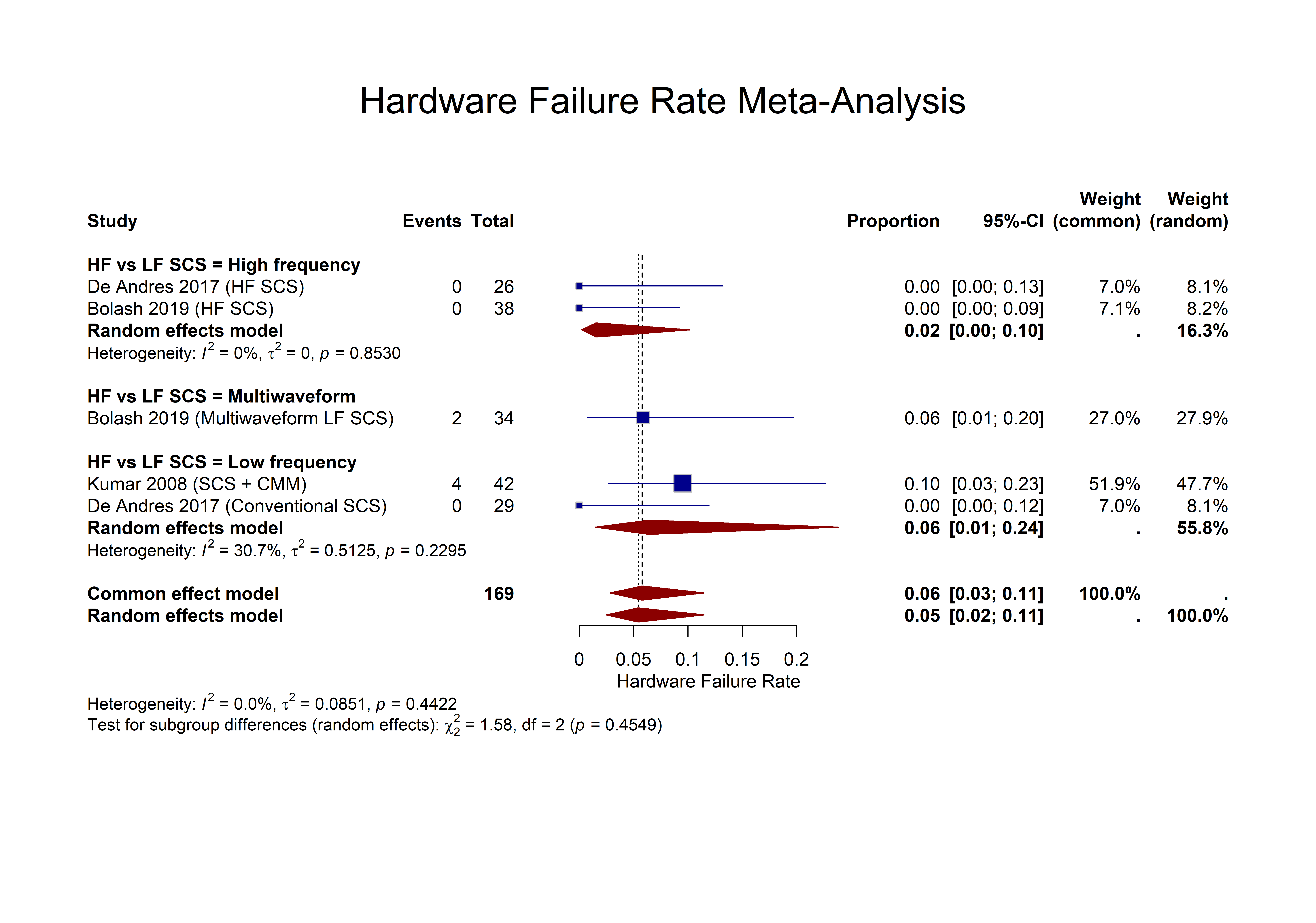

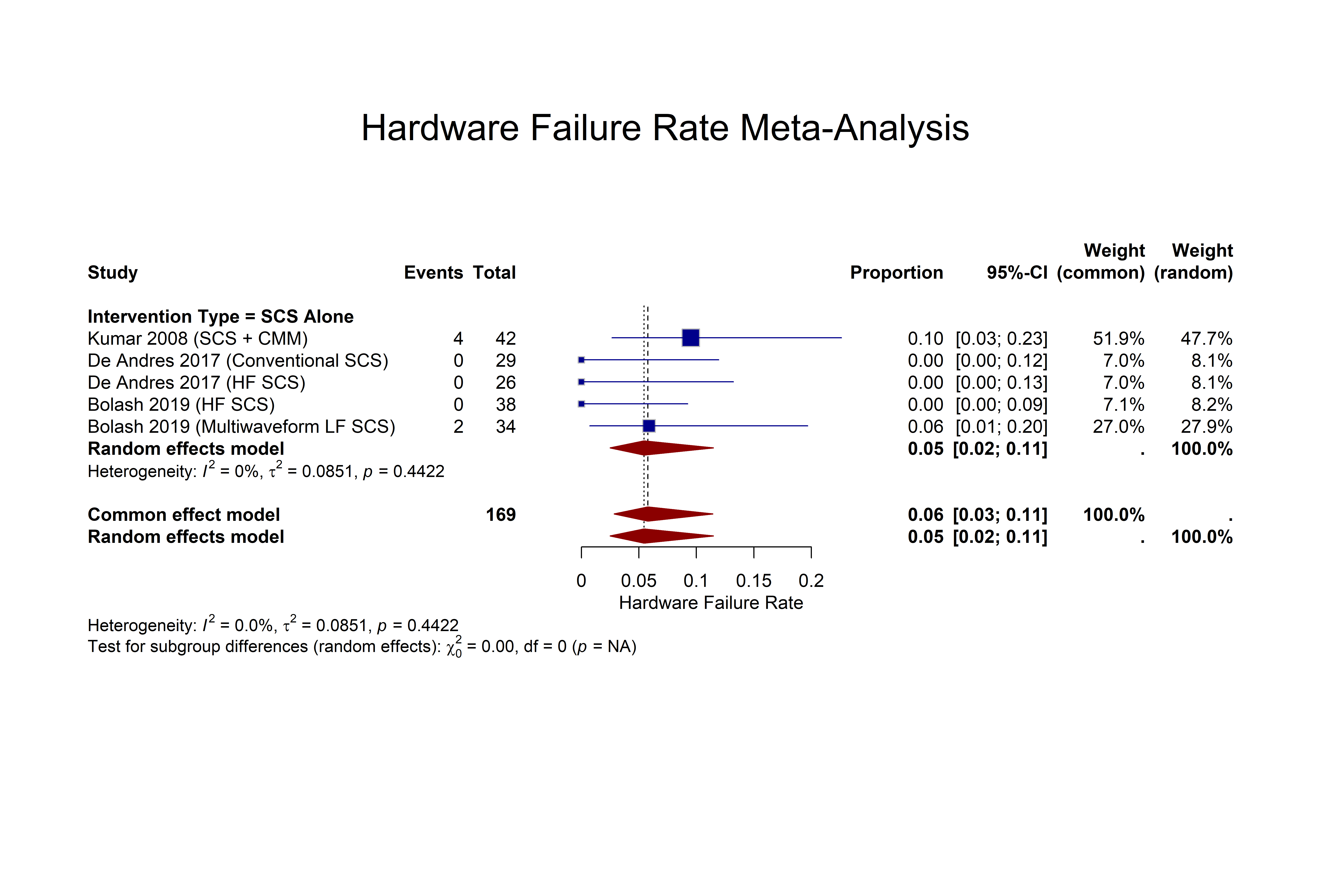

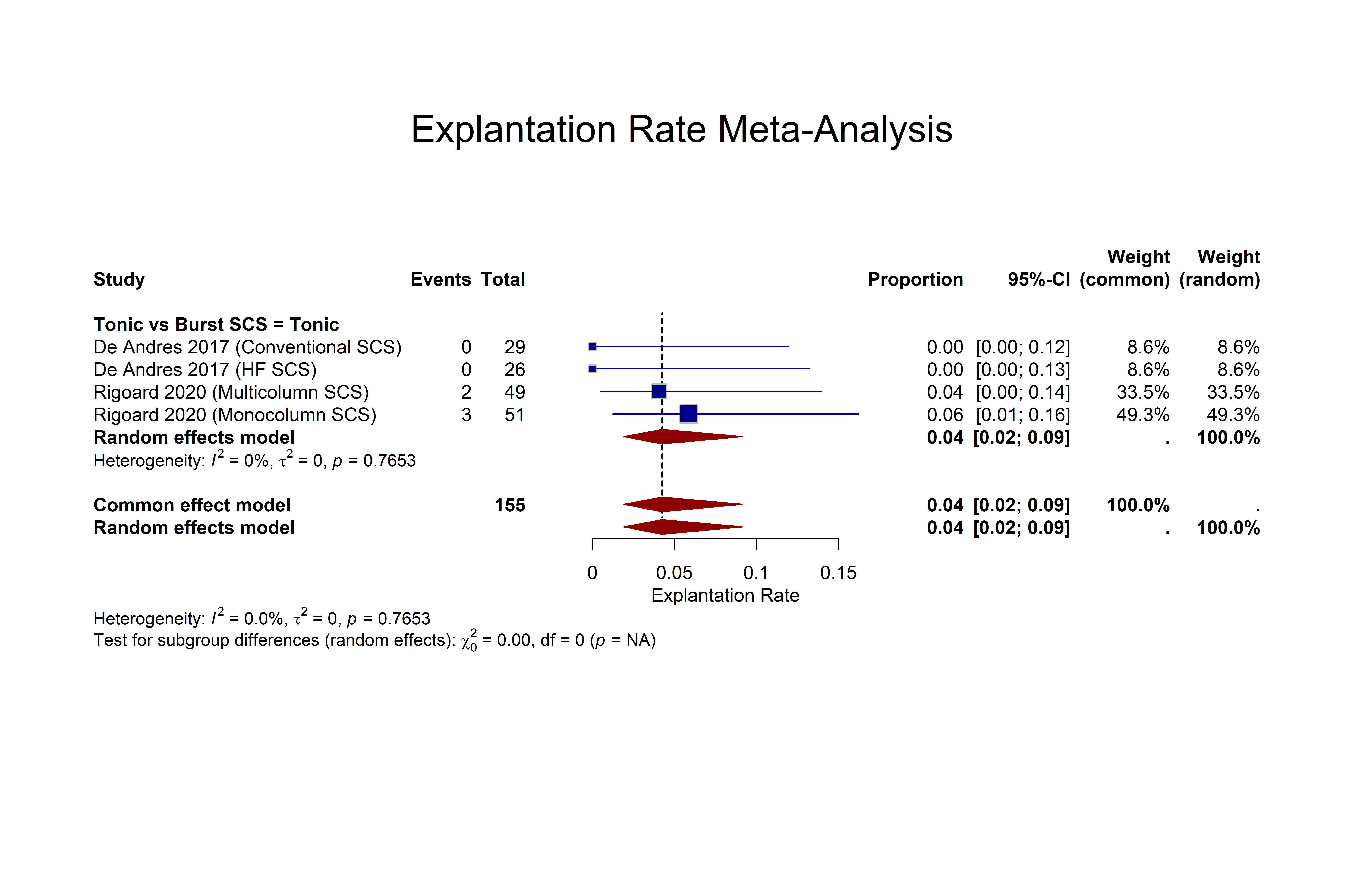

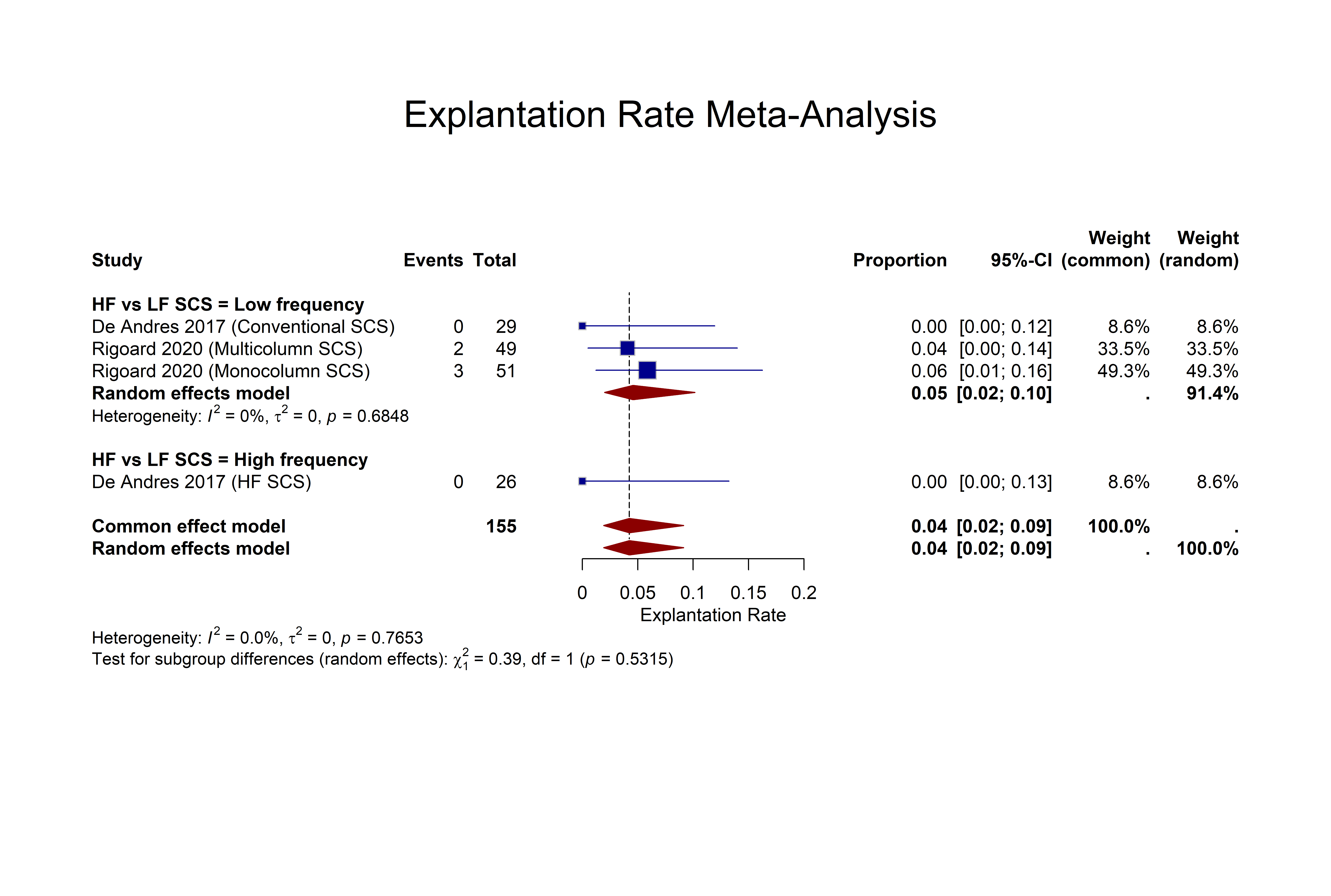

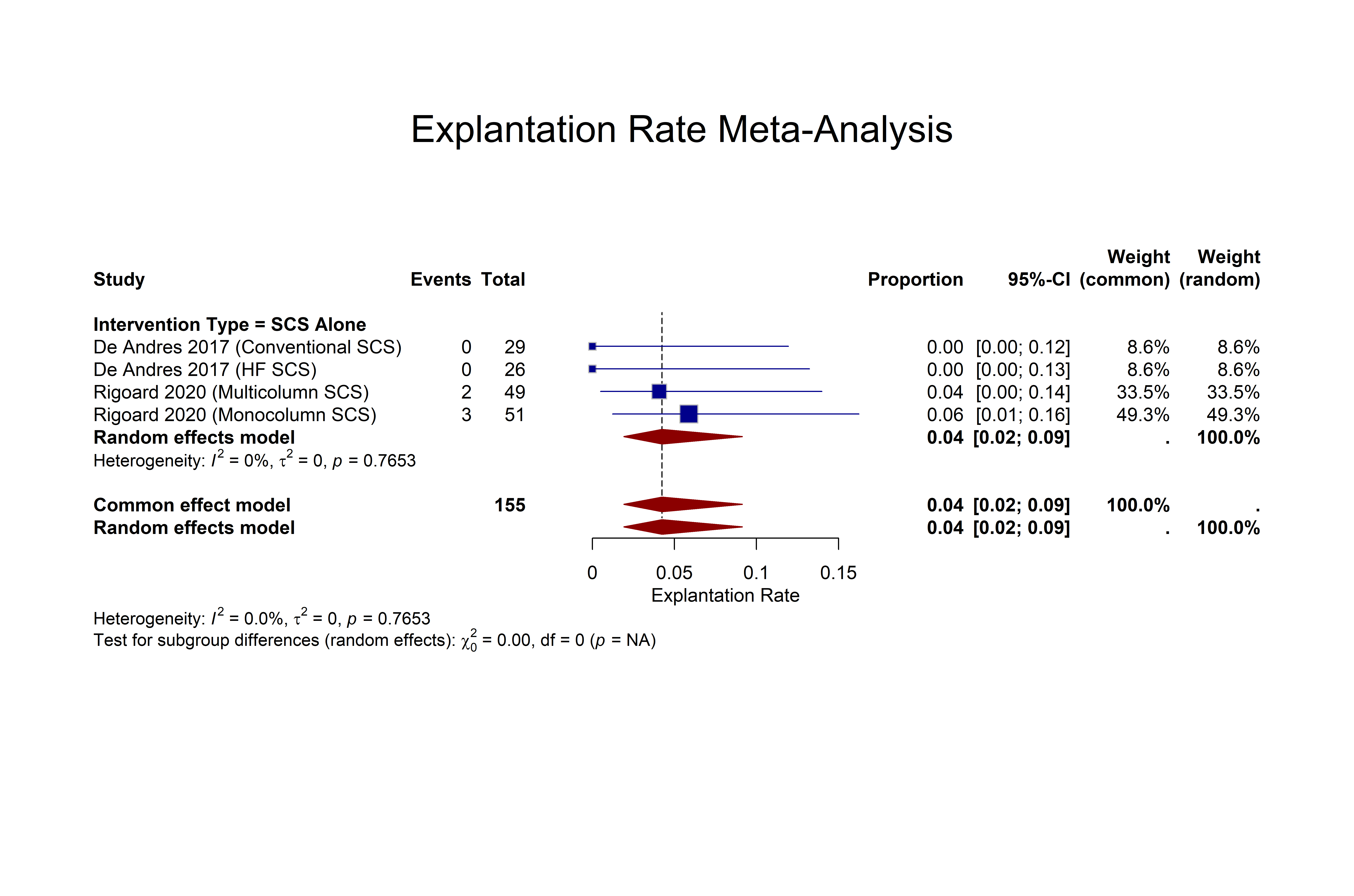

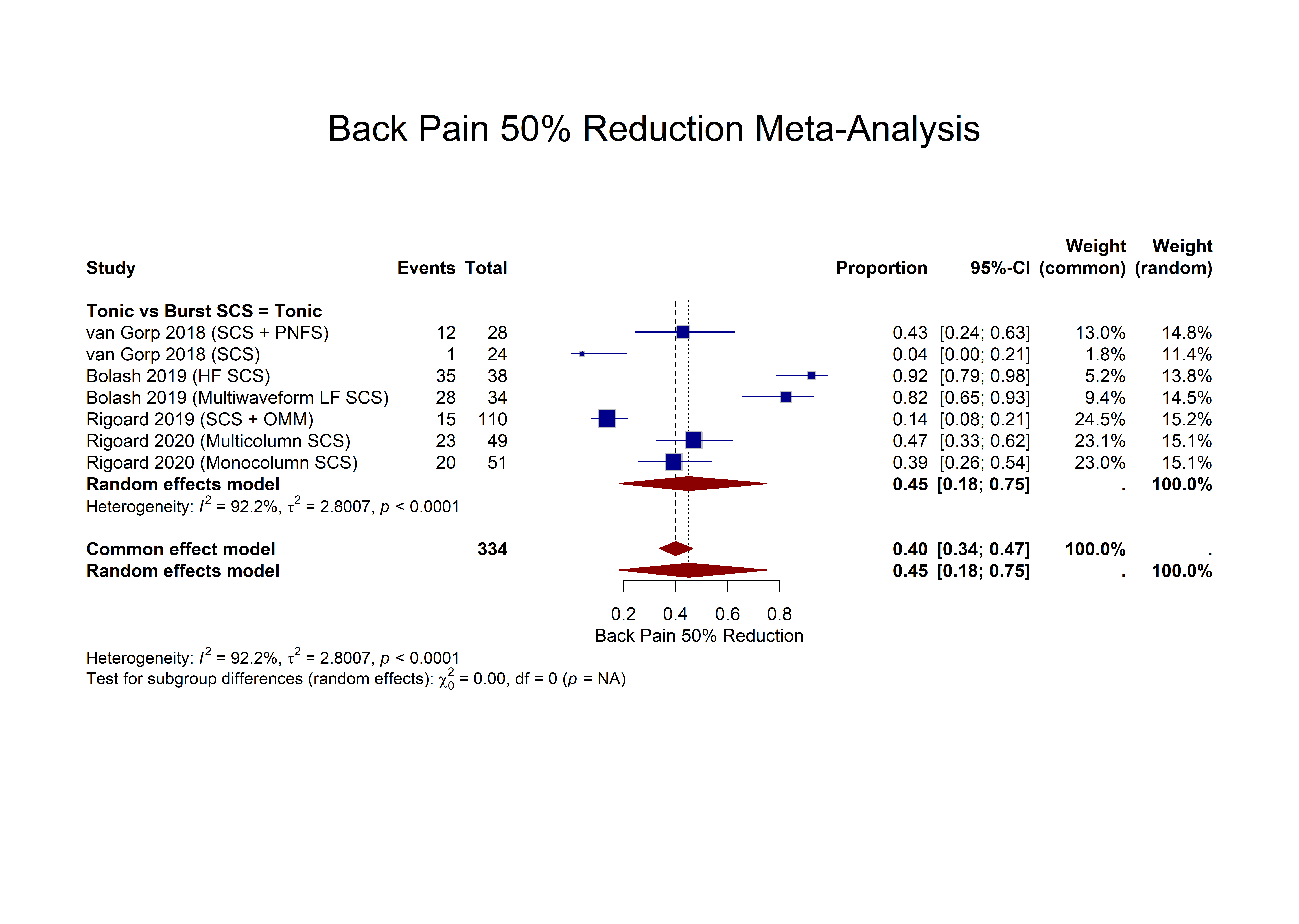

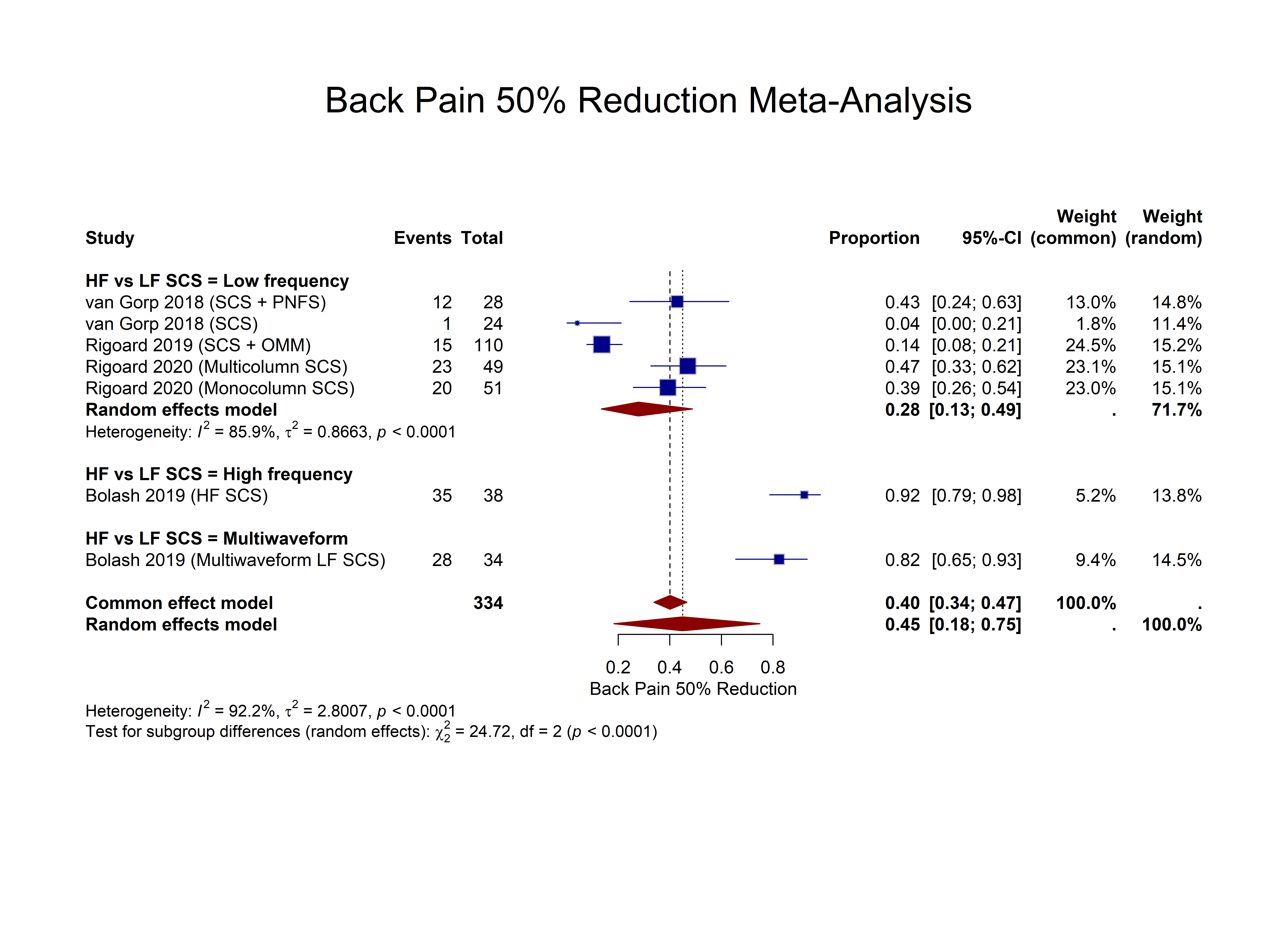

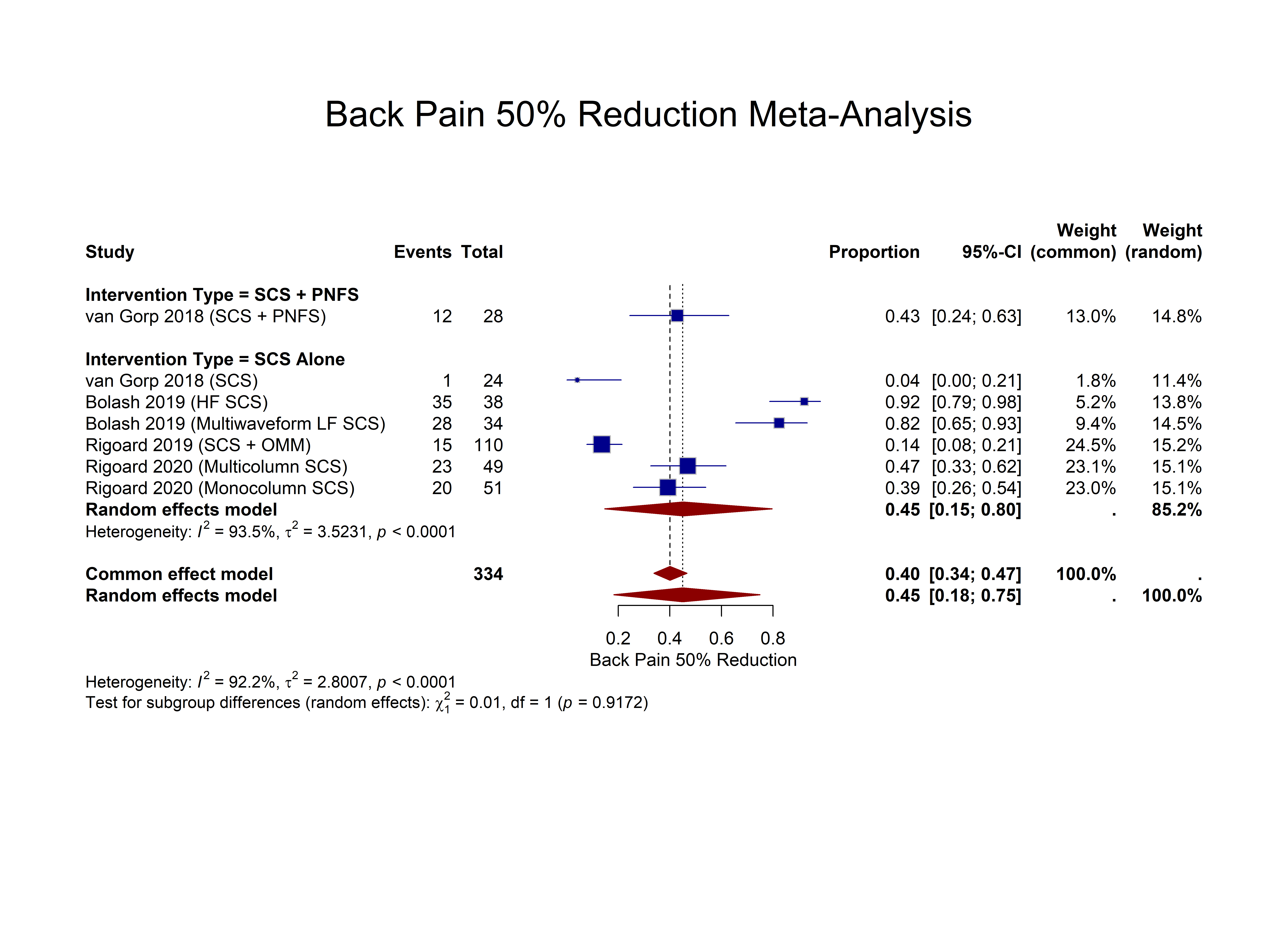

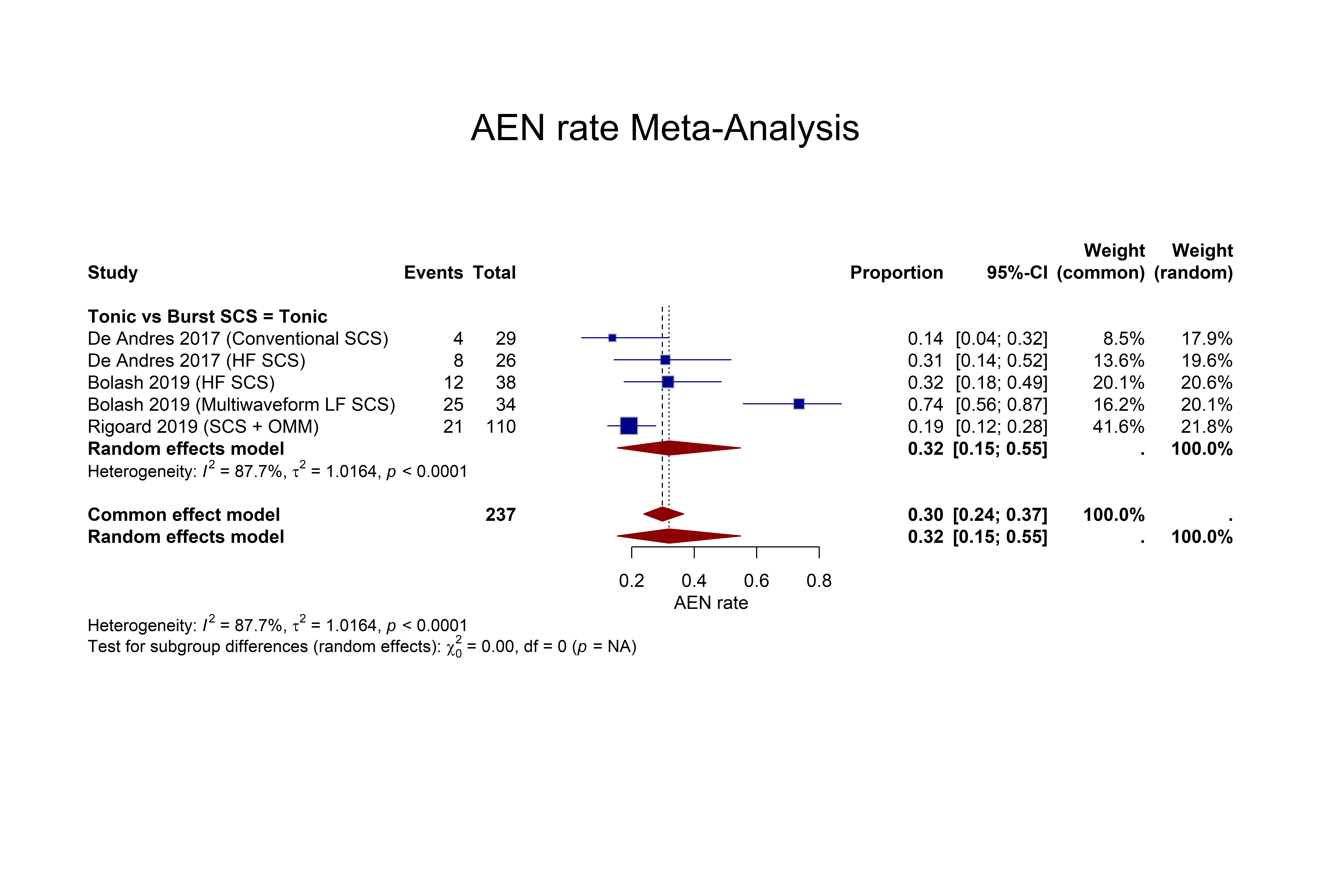

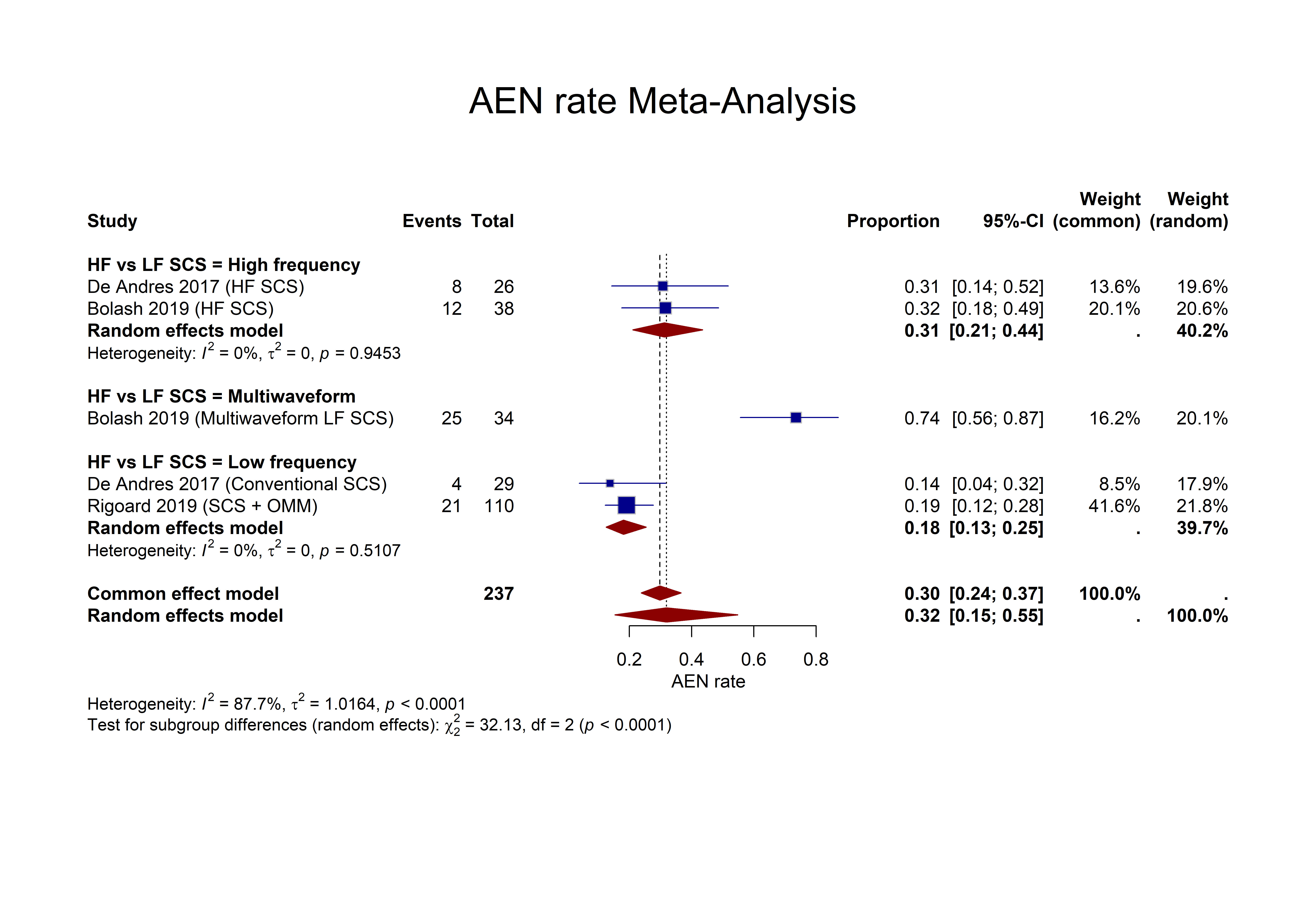

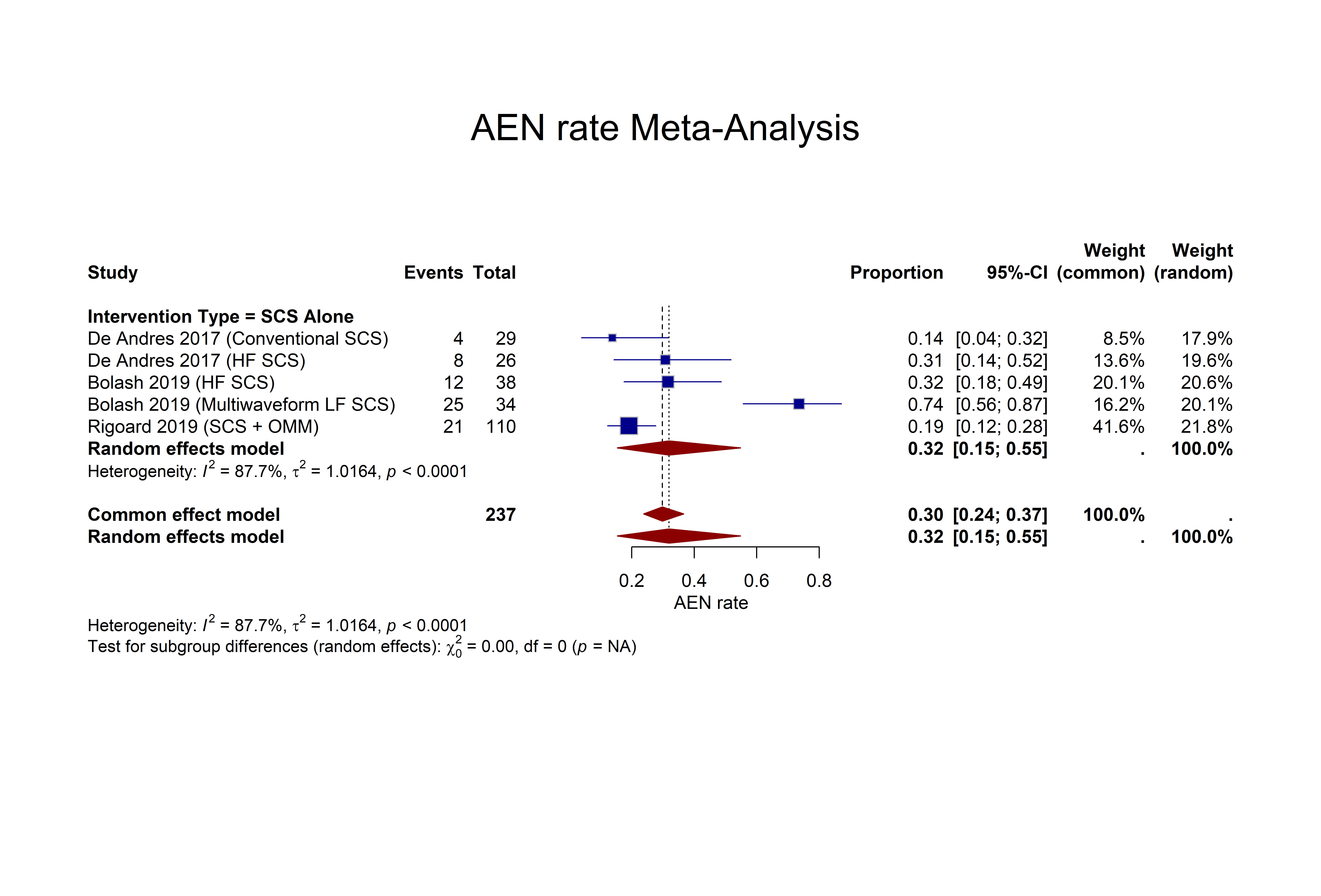

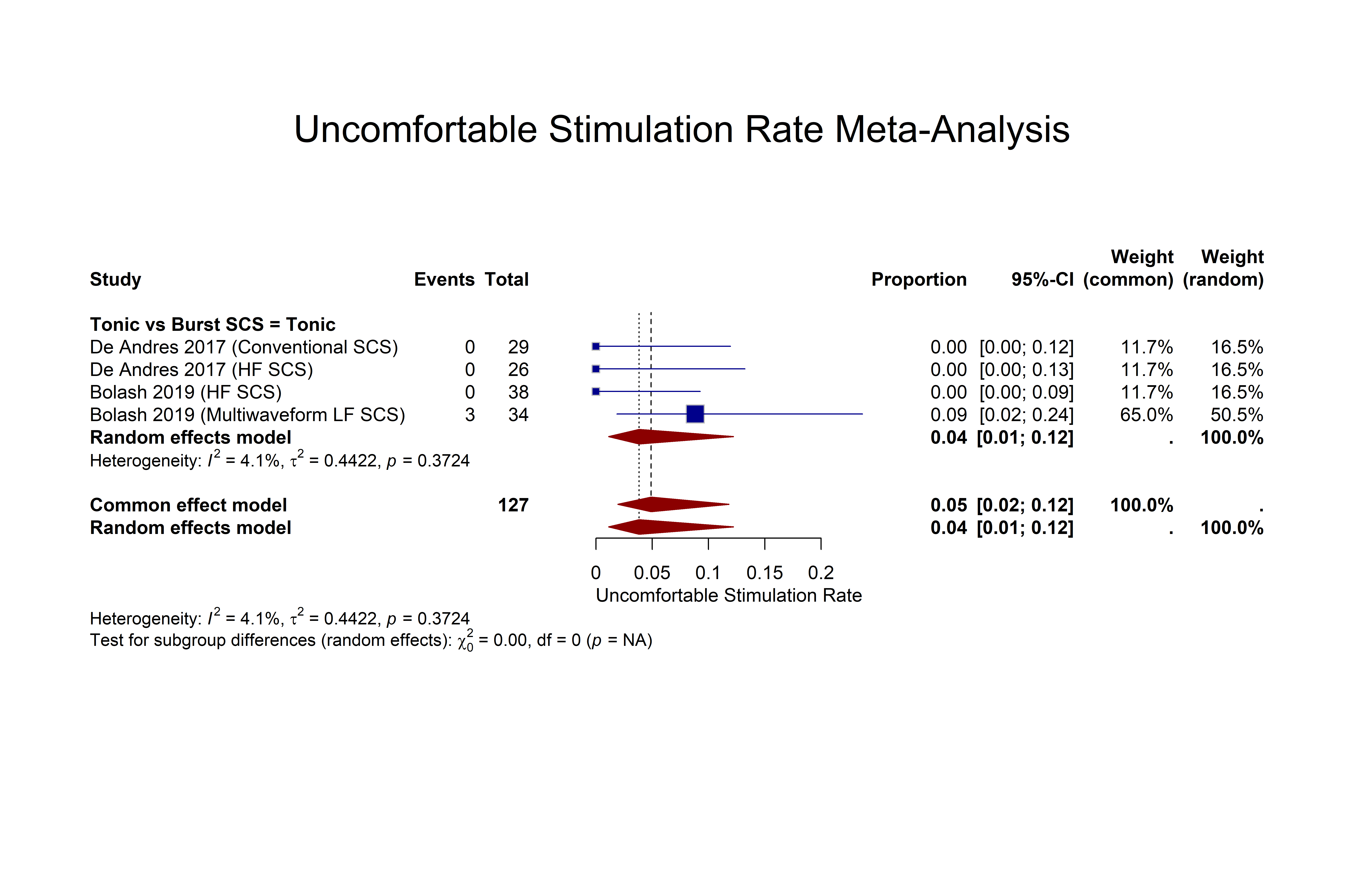
